# Brain-age in ultra-low-field MRI: how well does it work?

**DOI:** 10.1101/2025.10.19.25338298

**Authors:** Francesca Biondo, Carly Bennallick, Sophie A. Martin, Lemuel Puglisi, Thomas C. Booth, David A. Wood, Juan Eugenio Iglesias, František Váša, James H. Cole

## Abstract

**Introduction:** Brain-age is an estimate of the brain’s biological age derived from neuroimaging data, and has been proposed as a biomarker of brain health and disease risk. While brain-age estimation commonly uses high-field (HF) magnetic resonance imaging (MRI) (*>* 1.5 T) this is costly and inaccessible, limiting its applicability. Emerging ultra-low-field (ULF) MRI (*<* 0.1 T) technology is a cheaper and more accessible alternative, but its lower resolution raises questions about whether biomarkers like brain-age can be estimated reliably.

**Methods:** We assessed different brain-age pipelines in 23 adults scanned on one HF system (GE Signa Premier at 3 T) and two identical ULF systems (Hyperfine Swoop at 64 mT). 14 distinct acquisitions were used, defined by T1-or T2-weighting, resolution, and preprocessing: raw anisotropic orientations (axial, coronal, sagittal), isotropic scans, and super-resolution derivatives from multi-resolution registration (MRR) and SynthSR. These inputs (a total of *n* = 573 scans) were analysed with five brain-age software packages (BrainageR, SynthBA, MIDI, DeepBrainNet, Py-BrainAge). Performance evaluation entailed validity (brain-age vs. actual age), correspondence (ULF brain-age vs. HF brain-age), and test-retest reliability (ULF1 brain-age vs. ULF2 brain-age).

**Results:** Overall, performance was mixed across pipelines, though several ULF pipelines achieved performance comparable to HF. The four best-performing combinations were SynthBA on T2 scans without SynthSR, MIDI on T2 scans without SynthSR, PyBrainAge on T1 scans with SynthSR and using FreeSurfer `recon-all-clinical`, and BrainageR on T1 scans with SynthSR. These showed moderate-to-strong validity (*r* = 0.76–0.92, *R*^2^ = 0.54–0.64, MAE = 6.49–8.21 years), moderate-to-strong correspondence to HF (*r* = 0.84–0.93, ICC = 0.72–0.92), and excellent test-retest reliability (*r* = 0.97–0.99, ICC = 0.97–0.99). Moreover, some anisotropic acquisitions achieved comparable validity and reliability to MRR images when tested with the best-performing model, SynthBA (*R*^2^ = 0.57–0.62, ICC [CI] = 0.99 [0.97– 1.00], for coronal T2).

**Conclusion:** This first systematic evaluation of brain-age at ULF demonstrates that accurate and reliable estimates can be achieved across multiple pipelines, without necessarily requiring image enhancement. Performance depended on the combination of model, scan type, and preprocessing. ULF brain-age estimation could be a practical and scalable tool for clinical decision-making, population research, and long-term patient monitoring, thereby helping to make advanced neuroimaging biomarkers more accessible worldwide.

## 1. Introduction

Magnetic resonance imaging (MRI) biomarkers remain a cornerstone of brain health research. Over the past decade, their translational utility has expanded rapidly, providing valuable insights not only into brain structure, pathology and treatment monitoring across neurological and psychiatric conditions (Thompson et al., 2020; Young et al., 2024) but also into brain changes and differences in healthy individuals, for example in normative ageing studies (Bethlehem et al., 2022; Rutherford et al., 2022b,a; Dorfschmidt et al., 2025).

Despite these advances, MRI-derived markers are not without limitations. Conventional high-field (HF) MRI (1.5-3 T) systems remain largely inaccessible. This is largely driven by their high purchase and ongoing operational costs (e.g., cooling, trained staff) as well as complex installation requirements (e.g., shielding, structural reinforcement, upgraded power supply)(Arnold et al., 2023). Contraindications such as metal implants or claustrophobia further restrict their utility. These multiple barriers result in long wait lists, limited accessibility in resource-limited settings and a lack of representativeness in MRI cohorts for brain research (Martin et al., 2023; Abate et al., 2024).

Recently, a new generation of portable ultra-low-field (ULF) MRI systems (*<* 0.1 T) has emerged, offering a compelling alternative to conventional scanners. These inexpensive systems have negligible operational costs (e.g., low power, minimal training to run), can be operated from a standard power outlet, used at the bedside, and deployed in settings where HF MRI is impractical (Arnold et al., 2023; Kimberly et al., 2023; Sorby-Adams et al., 2024; Váša et al., 2025). Their open designs improve patient comfort and tolerance, and their reduced field strength minimises many common contraindications. These features make ULF MRI a promising avenue for increasing accessibility and broadening participation in neuroimaging research and clinical care. For example, their current deployment in low-to-middle-income countries for paediatric neurodevelopment research underscores their feasibility for large-scale global health studies (Abate et al., 2024; Pretzsch et al., 2025). In turn, increased accessibility and reduced cost make repeated scans far more feasible, facilitating longitudinal tracking of brain changes for both clinical and research purposes. However, ULF MRI comes at a cost, with lower signal-to-noise ratio (SNR) and reduced resolution compared with HF MRI (Arnold et al., 2023; Váša et al., 2025). Nonetheless, early findings suggest that ULF MRI can provide clinically meaningful information (Arnold et al., 2022; Deoni et al., 2022a; Sorby-Adams et al., 2024), and imaging denoising or enhancement techniques can help overcome its inherent resolution limitations (Deoni et al., 2022b; Iglesias et al., 2023; Briski et al., 2024; Baljer et al., 2025b,a). For example, multi-resolution registration (MRR) improves effective resolution and test-retest reliability (Deoni et al., 2022b; Váša et al., 2025), while machine learning-based super-resolution techniques such as SynthSR (Iglesias et al., 2023) enable the synthesis of isotropic, high-resolution images from low-quality inputs, improving segmentation accuracy and agreement with HF scans. Notably, Sorby-Adams et al.(2024) demonstrated that hippocampal and white matter hyperintensity measures derived from ULF scans processed with SynthSR are highly comparable to those from HF MRI. SynthSR is integrated into the widely used neuroimaging preprocessing software FreeSurfer (Fischl, 2012; Iglesias et al., 2023). In this study, we extend these advances to brain-age estimation, systematically evaluating this measure at ULF. Brain-age is an index of the brain’s biological age, typically obtained by training a machine learning model on MRI features from a healthy reference cohort to predict chronological age (Cole and Franke, 2017; Gaser et al., 2024). The trained model is then used to estimate age, or brain-age, in unseen scans. The difference between predicted and chronological age, commonly termed the brain-age delta or brain predicted age difference (brainPAD), serves as the key variable in downstream analyses. BrainPAD has emerged as a simple yet sensitive marker of brain health, providing clinically relevant information about both current status and future outcomes in patient and healthy cohorts, with older-than-expected brain-ages largely linked to negative outcomes such as cognitive decline, dementia risk and schizophrenia (Cole and Franke, 2017; Kaufmann et al., 2019; Wang et al., 2019; Biondo et al., 2022; Lee et al., 2022; Wrigglesworth et al., 2021; Wood et al., 2022; Mishra et al., 2023; Cumplido-Mayoral et al., 2024; Gaser et al., 2024).

Here, we test whether brain-age estimates from ULF MRI are valid and reliable. Leveraging a unique dataset (Váša et al., 2025) in which the same participants were scanned on a HF MRI system and two identical ULF scanners, we directly assess correspondence to HF benchmarks and test-retest reliability across ULF devices. To ensure a comprehensive evaluation, we apply five diverse brain-age models and explore multiple preprocessing pipelines, including MRR and SynthSR. The resulting set of combinations provides a rich framework for systematically characterising the potential of ULF MRI for brain-age estimation.

## 2. Materials and Methods

### 2.1 Data

Data were from the cohort described by Váša et al. (2025). Participants (n=23, aged 20–69 years) were recruited at King’s College London and St Thomas’s Hospital, and all provided informed consent. All reported being generally healthy and with no history of major neurological or psychiatric conditions. The sampling was designed to achieve approximate balance across sex and age decade (2–3 males and 2–3 females per decade). Each participant underwent scanning on a standard HF 3 T system (GE SIGNA Premier) and on two separate but identical ULF Hyperfine Swoop 64 mT systems (scanner software version was rc8.6.0). During the first session at King’s College London, T1- and T2-weighted images were acquired at 1 mm^3^ isotropic resolution on the 3 T system, followed immediately by ULF scans using both the scanner’s standard anisotropic sequences (axial (AXI), coronal (COR), sagittal (SAG); 1.5×1.5×5 mm^3^ for T2w, 1.6×1.6×5 mm^3^ for T1w) and an additional custom isotropic (ISO; 2.2 mm^3^ for T1w, 2.3 mm^3^ for T2w) sequence. A second ULF session was conducted on a different, identical scanner at St Thomas’s Hospital within a maximum of 36 days. This paired design permitted test-retest comparisons between ULF systems, as well as correspondence testing against the HF reference.

The scans from these three sources are referred to as HF (high-field at King’s College London), ULF1 (ultra-low-field at King’s College London) and ULF2 (ultralow-field at St Thomas’ Hospital).

### 2.2 Multi-resolution registration (MRR)

All anisotropic ULF images were processed using MRR (Deoni et al., 2022b), a form of super-resolution, which co-registers the three orthogonal acquisitions into a single higher-resolution volume (effective 1.5 mm^3^ for T2w, 1.6 mm^3^ for T1w). MRR allows integration of anisotropic acquisitions into isotropic space. MRR was implemented as previously described by Váša et al. (2025).

### 2.3 SynthSR v2 (SSR)

SynthSR is a neural-network-based super-resolution tool that converts clinical scans of arbitrary orientation, contrast and resolution into synthetic 1 mm^3^ isotropic T1-weighted MPRAGE-like images (Iglesias et al., 2023). In this study, version 2 of SynthSR was applied to the ULF scans that had been pre-processed with MRR. The implementation used FreeSurfer v8.0 with the –lowfield flag and default parameters. All scans, pre- and post-SynthSR processing, passed visual quality control (QC) using MRIcro^1^. See section SI 2.1 for details.

### 2.4. Scan types

The acquired MRI data, together with application of MRR and SynthSR, resulted in 14 distinct scan types. These are illustrated in Figure 1 (see also Table S1) and listed here: HF, 2 types: T1 and T2; ULF with MRR, 2 types: T1 and T2; ULF with MRR and SynthSR, 2 types: T1 SSR, T2 SSR; ULF single-acquisition, 8 types: T1 and T2 in AXI, COR, SAG, and ISO resolutions. For each scan type at each scanner site (HF, ULF1, ULF2), *n* = 23 participants were available, except for ULF1 T1 and its derivatives (*n* = 22) and for T2 ISO scans which were not acquired at ULF2.

**Figure 1.**
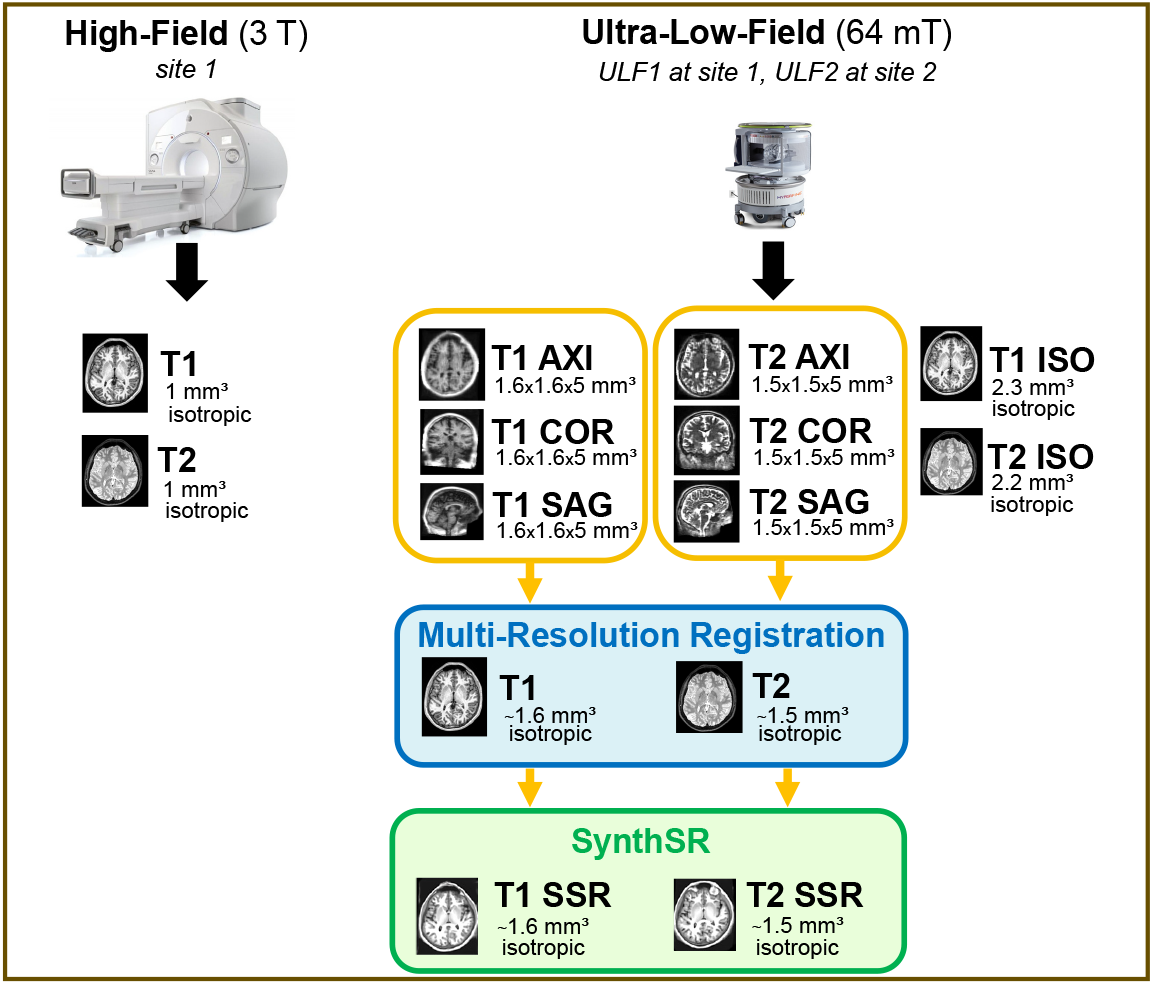
A total of 14 scan types were available. Two were acquired on the HF scanner: T1 and T2. Seven to eight single-acquisition scans were acquired on each ULF scanner: T1 and T2 in AXI, COR, SAG, and ISO orientations (with T2 ISO missing for ULF2). The three orthogonal ULF acquisitions for each scanner and contrast were then combined into an MRR volume of higher effective resolution, generating two additional scan types: T1 and T2. These MRR scans were further processed with SynthSR, resulting in two more scan types: T1 SSR and T2 SSR. For each scan type at each scanner site (HF, ULF1, ULF2), *n* = 23 participants were available, except for ULF1 T1 and its derivatives (*n* = 22). The total number of scans across scan types was *n* = 573. All five brain-age models were applied to six scan types: two HF (T1 and T2) and four ULF-derived images (T1, T2, T1 SSR, T2 SSR at both sites). The remaining eight ULF scan types were evaluated only with the best-performing brain-age model. Abbreviations: HF, highfield; ULF1/ULF2, ultra-low-field site 1/site 2; MRR, multi-resolution registration; SSR, SynthSR; AXI, axial; SAG, sagittal; COR, coronal; ISO, isotropic.

Given the large amount of scans (a total of *n* = 573), the main analyses focused on HF, ULF+MRR and ULF+MRR+SSR scans (six scan types, *n* = 228 in total). The remaining eight ULF single acquisition and ISO scan types were used only once with the best-performing brain-age model.

### 2.5. Brain-age models

Five different brain-age models were used to generate age predictions for each scan. Each model has its own preprocessing pipeline, typically involving image registration and/or tissue segmentation. After preprocessing, the scan quality was assessed using a QC procedure specific to each model. The deep-learning models (SynthBA, DeepBrainNet, MIDI) had the common requirement for input scans to be skull-stripped and linearly registered to a standard template. To streamline this, SynthBA’s preprocessing step was used for all three deep-learning models. The preprocessing and QC procedure for each model is described below and Table 1 summarises which model was applied to which scan.

**Table 1:** Brain-age models (rows) applied to which scan types (columns)

| Model | HF<br>T1 | HF<br>T2 | ULF<br>T1 | ULF<br>T2 | ULF<br>T1_SSR | ULF<br>T2_SSR | HF<br>T1+T2 | ULF<br>T1+T2 |
| --- | --- | --- | --- | --- | --- | --- | --- | --- |
| BrainageR | Yes | n/a | Yes | n/a | Yes | Yes<sup>†</sup> | n/a | n/a |
| SynthBA | Yes | Yes | Yes | Yes | Yes | Yes | n/a | n/a |
| MIDI-T1 | Yes | n/a | Yes | n/a | Yes | Yes | n/a | n/a |
| MIDI-T2 | n/a | Yes | n/a | Yes | n/a | n/a | n/a | n/a |
| DeepBrainNet | Yes | n/a | Yes | n/a | Yes | Yes<sup>†</sup> | n/a | n/a |
| PyBrainAge + FS v7.4 ra | Yes | n/a | Yes | n/a | Yes | Yes | Yes | Yes |
| PyBrainAge + FS v7.4 rac | Yes | Yes | Yes | Yes | No | No | n/a | n/a |
| PyBrainAge + FS v8.0 ra | Yes | n/a | Yes | n/a | No | No | Yes | Yes |
| PyBrainAge + FS v8.0 rac | Yes | Yes | Yes | Yes | Yes | Yes | n/a | n/a |

All ULF scans listed here were pre-processed with MRR; “T1 SSR/T2 SSR” indicates additional SynthSR processing. The combined-input columns “HF/ULF T1+T2” denotes FreeSurfer `recon-all (ra)` runs that used a T1 with an accompanying T2 via the `-T2` and `-T2pial` flags and are only applicable to the PyBrainAge model; Single-acquisition ULF (AXI, COR, SAG, ISO) scans are omitted in this table because they were evaluated only with the best-performing model (see Results). ^*†*^ BrainageR and DeepBrainNet accept T1-weighted and SynthSR-derived (T1-like) inputs; here T2 SSR is treated as T1-like; ra = *recon-all*; rac = *recon-all-clinical*.

#### 2.5.1. BrainageR

BrainageR^2^ is a Gaussian Process Regression model trained on T1-weighted scans from 3,377 healthy adults (18-92 years) drawn from public datasets^3^. The model operates on principal components derived from voxel-wise grey-matter, white-matter and CSF tissue-probability maps.

BrainageR performs an initial preprocessing step involving tissue segmentation and spatial normalisation to MNI152 space using SPM12, after which the pretrained model was applied to produce brain-age estimates. The quality of the preprocessing was assessed visually for each scan using the slice screenshots included in the BrainageR output, and all scans passed QC. As BrainageR was trained on T1- weighted MRI scans, we tested it on T1-weighted and SynthSR-derived images only.

#### 2.5.2. SynthBA

SynthBA^4^ (Puglisi et al., 2024) is a brain-age model built on a 3D DenseNet and trained on synthetic MRIs generated from brain-tissue segmentations via a synthetic scan generator (Gopinath et al., 2024) with extensive domain randomisation. Each synthetic scan is produced with randomised contrast, resolution, noise, bias fields, and intensity profiles, enabling the model to learn contrast- and resolution-agnostic features and thus generalise across acquisition protocols without retraining. The training segmentations were sourced from 4,105 healthy subjects, and Gaussian priors for the generator were estimated from 7,060 T1-weighted and 1,689 T2-weighted MRIs drawn from public datasets. Preprocessing involved skull-stripping using SynthStrip (Hoopes et al., 2022) followed by linear registration to the MNI152 template with Advanced Normalization Tools (ANTs, Tustison et al. (2021)). Preprocessed scans underwent quantitative and visual QC (see SI 2.4 for details). All scans passed QC and were used as direct input for SynthBA, DeepBrainNet, and MIDI. In this study, SynthBA was tested on T1-weighted, T2-weighted, and SynthSR-derived volumes.

#### 2.5.3. DeepBrainNet (DBN)

DBN^5^ is a 2D convolutional model based on an Inception-ResNet-v2 backbone, trained on 11,729 T1-weighted scans from a large, multi-study lifespan cohort (ages 3-95 years) with minimal preprocessing (skull-stripping and affine registration) (Bashyam et al., 2020). Each scan is represented by 80 axial slices and slice-level predictions are aggregated (median) to provide a scan-level brain-age estimate. In this study, DBN was tested on T1-weighted and SynthSR-derived volumes.

#### 2.5.4. MIDI

Two pretrained models were used from the MIDI BrainAge release^6^ (Wood et al., 2022). First, the *T2 model* (3D DenseNet-121) trained on 23,302 axial T2-weighted scans from two UK hospital networks (ages 18-95) with minimal preprocessing (DICOM-to-NIfTI, 1 mm resampling, cropping/padding, *z*-score normalisation; no registration or skull-stripping). Second, the *T1 model* (3D DenseNet-121) trained on 1,551 volumetric T1-weighted scans from a UK-based research cohort. We used the `--ensemble` option, which aggregates predictions across multiple random-seed checkpoints of the same T1 model to produce a single estimate.

In this study, the MIDI-T2 model was tested on T2-weighted inputs, and the MIDI-T1 ensemble was applied to T1-weighted and SynthSR-derived volumes. Because input scans were already skull-stripped and linearly registered, the MIDI “skull-stripped” settings were used, and the default preprocessing steps (skull-stripping and registration to MNI152 using ANTs) were omitted.

#### 2.5.5. PyBrainAge

PyBrainAge^7^ represents a more classical, feature-based approach, leveraging a well-established neuroimaging pipeline by using ridge regression on morphometric features extracted with FreeSurfer^8^ (Fischl, 2012). These include cortical thickness, surface area, and volumetric features from the Desikan-Killiany atlas. PyBrainAge was trained on 29,175 T1-weighted scans from healthy participants aged 2-100 years, across 76 different sites (mostly UK Biobank data) (Rutherford et al., 2022a).

To generate PyBrainAge inputs, FreeSurfer was run with a set of planned variations across (i) FreeSurfer version (either v7.4.0 or v8.0.0 beta), (ii) pipeline (`reconall` vs `recon-all-clinical`), and (iii) input scan type. Not all combinations were exhausted: preliminary tests suggested that `recon-all-clinical` behaved similarly across v7.4.0 and v8.0.0, and performed better than `recon-all` on ULF data, so those options were prioritised.

##### In FreeSurfer v7.4.0

`recon-all` was tested on T1-weighted and SynthSR-derived scans as well as pairs of T1-weighted and T2-weighted scans using the `-T2` and `-T2pial` flags. The command `recon-all-clinical` was run on T1-weighted and T2-weighted scans without SynthSR.

##### In FreeSurfer v8.0.0

`recon-all` was tested on T1-weighted scans without SynthSR (HF T1, ULF T1) as well as T1-weighted paired with T2-weighted using the `-T2` and `-T2pial` flags. The command `recon-all-clinical` was tested on T1-weighted, T2-weighted and SynthSR-derived volumes.

##### A note on the FreeSurfer variations

The conventional `recon-all` command in FreeSurfer is an iterative, surface-based workflow optimised for high-resolution T1- weighted research scans (Fischl, 2012). By contrast, `recon-all-clinical` (introduced in v7.4) was specifically developed to extend surface reconstruction and cortical thickness estimation to heterogeneous clinical scans (e.g., T1, T2, FLAIR) acquired at variable resolutions (Gopinath et al., 2025). Its key innovation is the use of domain randomisation during training: a 3D U-Net is trained to predict isotropic signed-distance functions (SDFs) from highly augmented synthetic data that spans a wide range of contrasts, slice thicknesses, noise levels, and orientations. This enables the model to generalise to “in-the-wild” clinical data without retraining. The architecture combines this learning-based SDF prediction (referred to as “SynthDist”^9^) with FreeSurfer’s classical geometry processing to enforce topology and generate standard surfaces. In parallel, SynthSeg (Billot et al., 2023a,b) provides robust volumetric labelling across contrasts and resolutions, while SynthSR is used only for visualisation and not for downstream processing (Gopinath et al., 2025).

##### Harmonising FreeSurfer outputs for PyBrainAge

The label sets from `recon-all` and `recon-all-clinical` are not identical; some structures are absent in the latter, and some common labels (notably the region denoted “CSF”) differ in neuroanatomical extent. Because PyBrainAge expects inputs derived from v7-like `recon-all` segmentations, a small set of targets (right/left vessel, right/left choroid plexus, and CSF) were predicted using a compact predictive model trained only on `recon-all` (v7.4) volumetric segmentations from the high-field data in this study. Any remaining sporadic missingness was handled with k-nearest neighbours imputation. See SI section 3 for details.

##### Quality control of FreeSurfer outputs

Quantitative and visual checks were combined. For `recon-all`, the free FSQC tool^10^ was used, along with the examination of FreeSurfer statistics (e.g., the number of surface holes per hemisphere). At the time of analyses, FSQC was not compatible with `recon-all-clinical`, hence for these cases, visual inspection was run using an in-house custom tool, whilst the quantitative check was run using the SynthSeg QC metrics included in the output, where a CNN-based regressor outputs scores (0–1) approximating Dice values for key brain structures, enabling unreliable segmentations to be flagged without manual ground truth (Billot et al., 2023a,b). See SI section 2.3 for details.

The QC results were mixed. The T1+T2 `recon-all` pipeline performed poorly overall, with 5 scans (v7.4) and 4 scans (v8.0) failing QC and the remaining ULF scans being borderline failures. For `recon-all` applied to SynthSR-derived T2 volumes 13 scans failed QC. Native ULF `recon-all` runs often appeared suboptimal on visual inspection, but none failed QC. Retaining potentially-suboptimal scans was a deliberate choice to account for the expected lower resolution of the ULF scans. See SI 2.2 for details. No other scan failed QC.

### 2.6. Evaluation metrics

Each brain-age model combination was assessed in three ways: (i) validity, (ii) HF-ULF correspondence and (iii) test-retest reliability.

Validity was quantified as the agreement between predicted brain-age and actual (chronological) age. Following recommendations for predictive modelling in neuroimaging (Scheinost et al., 2019), this included the predictive *R*^2^ (computed directly from predicted vs. observed values, as implemented in `scikit-learn` Pedregosa et al. (2011)), alongside Pearson’s correlation (*r*) and mean absolute error (MAE).

Correspondence was quantified as the agreement between brain-age predictions from HF and ULF scans, and test-retest reliability was quantified as agreement between the two ULF scanners, both using the intraclass correlation coefficient (ICC) and the absolute symmetric percent difference (ASPD).

ICC was computed using a two-way mixed-effects model for absolute agreement and reported with 95% confidence intervals. The ASPD, as used in Sorby-Adams et al. (2024), quantifies the percentage difference between two paired measurements*X* and *Y* :

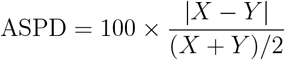

Higher ICC and lower ASPD both indicate better agreement. ICC summarises agreement and consistency while accounting for between-participant variability, whereas ASPD directly measures the magnitude of pairwise differences and is less influenced by the between-participant variance.

To visually assess the agreement between brain-age predictions from different scan types, Bland-Altman plots were used. For each pairing, the difference against the pairwise mean was plotted to visualise bias (mean difference) and the 95% limits of agreement (mean *±* 1.96 SD).

In the main text, correspondence and reliability results are reported for HF versus ULF1 only, since these were acquired on the same day at the same site and thus provide the most direct comparison. Complete results, including HF versus ULF2, are provided in SI section 4.

### 2.7. Model performance composite rankings

There were a total of 40 unique combinations of brain-age model, preprocessing pipeline, scanner and scan type. Of these, 26 pertained to ULF, which are the combinations of interest and are referred to as test runs. To summarise performance across metrics and make these test runs comparable, each performance metric was ranked separately within validity, correspondence, and test-retest reliability. Ranks for MAE and ASPD were inverted to keep directions consistent. For each test run, the median rank across metrics was calculated thus obtaining a single validity ranking (brain-age vs. actual age), correspondence ranking (HF vs. ULF1), and test-retest reliability ranking (ULF1 vs. ULF2).

Due to space constraints, we show validity, correspondence, and reliability plots only for the top four performing models, selected based on validity rankings. Validity was prioritised over correspondence and reliability as it reflects proximity to the ground truth (i.e., age). Full plots for all models are available in SI section 5.

#### 2.7.1. Application of the “best-performing model” to single-acquisition ULF scans

The best-performing model was next applied to the ULF single-acquisition sequences - the anisotropic scans (AXI, COR, SAG) and the isotropic ISO sequence - which had not been tested with any brain-age model in the main analyses. These scans are faster to acquire but lower in resolution than the MRR reconstructions. The preprocessing protocol for the best-performing model (see Methods section 2) was applied to these single-acquisition scans, all of which passed QC.

## 3. Results

Performance metrics for the total 40 combinations of brain-age models, preprocessing, scanner and scan type are presented in Tables 2–3. The rankings for the 26 test runs (pertaining to ULF data only) are presented alongside the raw metrics in the same tables. ULF2 correspondence and reliability results can be found in a larger table in SI section 4.

**Table 2:**
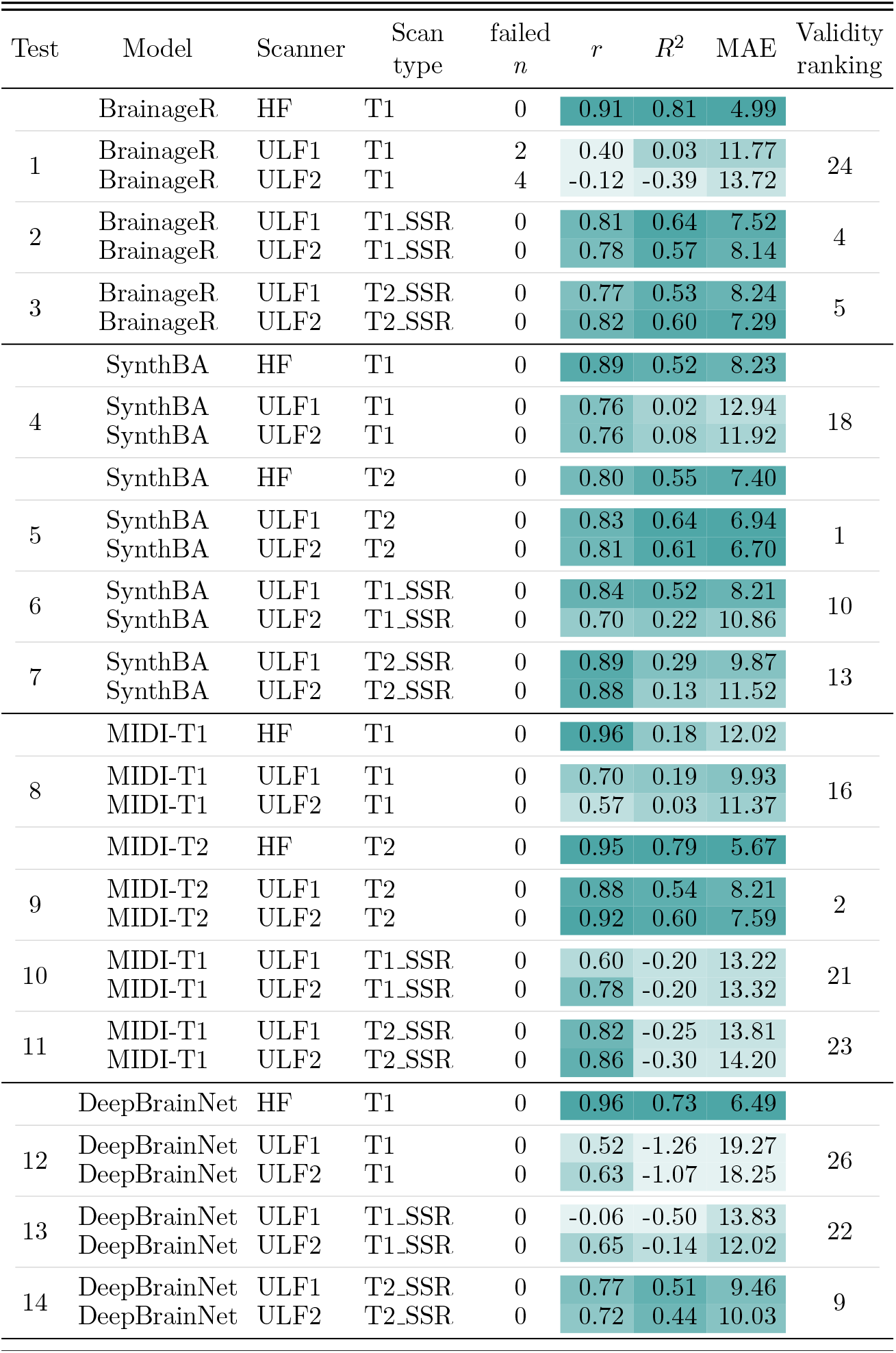
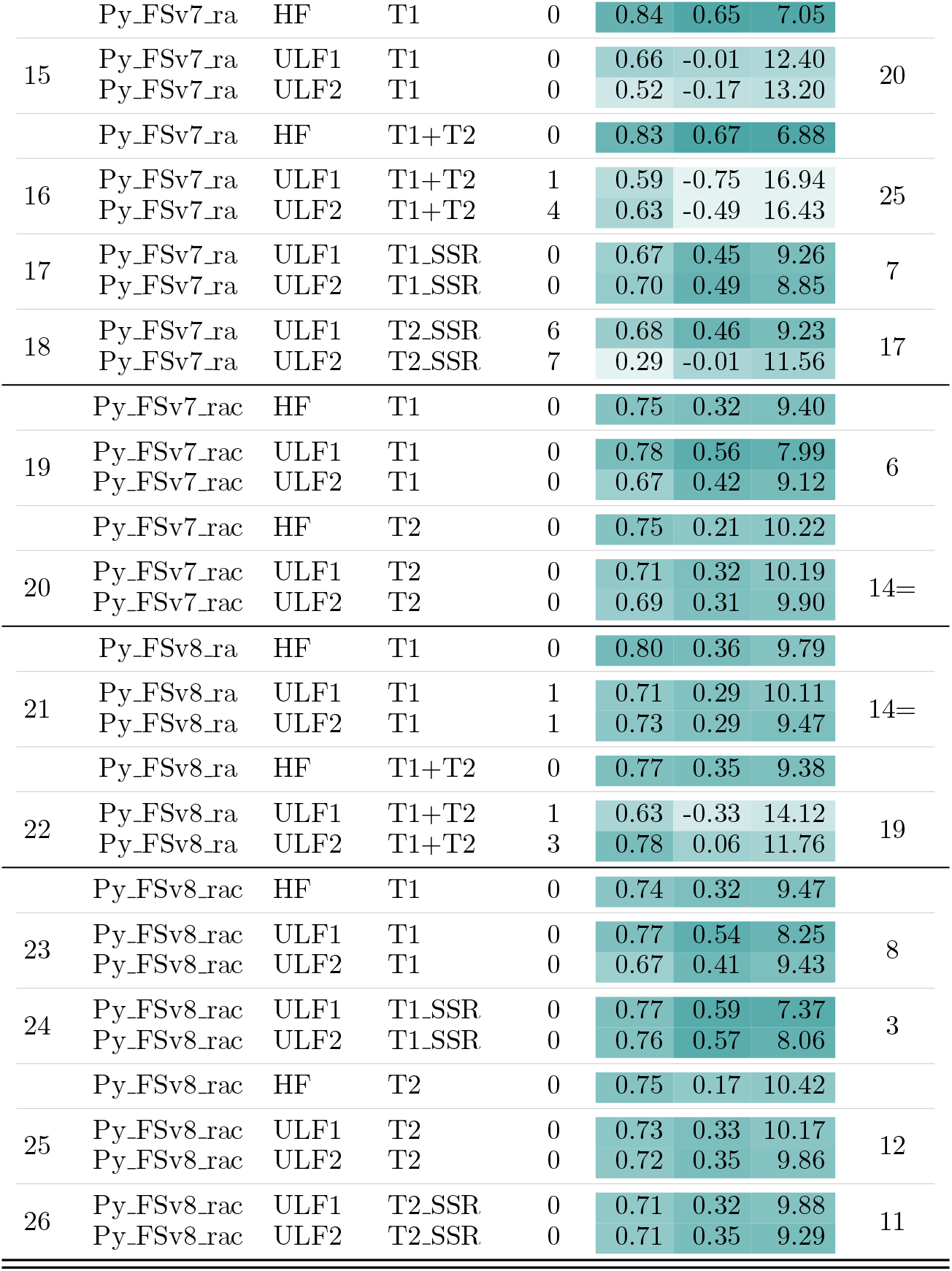
Validity. All conditions start with *n* = 23, except for ULF1 T1 derived conditions (*n* = 22). The number of scans removed due to failed preprocessing/QC is shown in “failed *n*”. The validity ranking is based on the median of six per-metric ranks within each “Test” run (i.e., the three performance metrics, each evaluated for ULF1 and ULF2, are first converted to ranks, and the median of those six ranks is then used for the final ranking). Numerically-low ranks and dark colours in the heatmap indicate better performance whilst numerically-high ranks and light colours indicate poor performance; Abbreviations: HF: high-field; ULF1/ULF2: ultra-low-field at site 1/site 2; SSR: SynthSR; (*r*): Pearson’s correlation coefficient; *R*^2^: coefficient of determination; MAE: mean absolute error; “=“ denotes tied ranks; Py FSv7 ra: PyBrainAge using Freesurfer (FS) v7.4 and (`recon-all`); Py FSv8 rac: PyBrainAge using FS v8.0 and (`recon-all-clinical`).

**Table 3:** Correspondence and reliability. Correspondence denotes agreement between ULF1 scans and a HF homologue (see “HF type” column). Test–retest reliability denotes agreement between ULF1 and ULF2. Starting sample size is *n* = 23 except for ULF1 T1 derived scans *n* = 22. Max scans excluded due to failed preprocessing/QC shown in “max failed *n*” (refer to ULF only; all HF passed). Correspondence and test–retest rankings are based on the median of three per-metric ranks, where each of the metrics, *r*, ASPD and ICC is first converted to a rank and then the median of these three ranks is used. Numerically-low ranks and dark colours in the heatmap indicate good performance whilst numerically-high ranks and light colours indicate poor performance; Abbreviations: HF, high-field; ULF1/ULF2, ultra-low-field at site 1/site 2; SSR, SynthSR; *r*, Pearson correlation; ASPD, absolute symmetric percent difference; ICC, intraclass correlation coefficient; CI, confidence interval; “=“ denotes tied ranks; Py FSv7 ra, PyBrainAge using Freesurfer (FS) v7.4 and (`recon-all`); Py FSv8 rac, PyBrainAge using FS v8.0 and (`recon-all-clinical`).

| Test | Model | Scan type | HF type | max failed $n$ | Correspondence | | | | | Test-retest | | | | | | |
| --- | --- | --- | --- | --- | --- | --- | --- | --- | --- | --- | --- | --- | --- | --- | --- | --- |
| | | | | | $r$ | ASPD(CI) | | ICC(CI) | | Ranking | $r$ | ASPD(CI) | | ICC(CI) | | Ranking |
| 1 | BrainageR | T1 | T1 | 6 | 0.37 | 15.3 | (7.5-34.1) | 0.30 | (-0.15-0.65) | 23 | 0.37 | 3.2 | (1.1-19.7) | 0.38 | (-0.14-0.73) | 25 |
| 2 | BrainageR | T1.SSR | T1 | 0 | 0.84 | 16.6 | (6.8-23.5) | 0.77 | (0.52-0.90) | 7 | 0.97 | 3.3 | (1.0-6.7) | 0.97 | (0.93-0.99) | 5 |
| 3 | BrainageR | T2.SSR | T1 | 0 | 0.86 | 17.3 | (9.1-25.4) | 0.77 | (0.35-0.91) | 8 | 0.97 | 5.0 | (3.1-8.7) | 0.97 | (0.93-0.99) | 8 |
| 4 | SynthBA | T1 | T1 | 0 | 0.84 | 16.7 | (7.3-28.2) | 0.69 | (0.19-0.88) | 9 | 0.98 | 4.0 | (2.3-7.4) | 0.98 | (0.95-0.99) | 2= |
| 5 | SynthBA | T2 | T2 | 0 | 0.93 | 7.6 | (2.4-16.4) | 0.92 | (0.80-0.97) | 1 | 0.99 | 1.7 | (1.0-3.7) | 0.99 | (0.99-1.00) | 1 |
| 6 | SynthBA | T1.SSR | T1 | 0 | 0.68 | 27.1 | (14.7-40.7) | 0.64 | (0.31-0.83) | 21 | 0.86 | 10.9 | (5.0-25.2) | 0.86 | (0.69-0.94) | 19 |
| 7 | SynthBA | T2.SSR | T2 | 0 | 0.81 | 34.6 | (20.0-57.5) | 0.57 | (-0.08-0.85) | 16 | 0.97 | 8.0 | (2.1-17.5) | 0.97 | (0.93-0.99) | 8 |
| 8 | MIDI-T1 | T1 | T1 | 0 | 0.67 | 24.8 | (13.0-36.3) | 0.47 | (0.02-0.75) | 18 | 0.82 | 4.9 | (2.4-9.1) | 0.82 | (0.62-0.92) | 20= |
| 9 | MIDI-T2 | T2 | T2 | 0 | 0.93 | 12.9 | (5.6-22.3) | 0.81 | (0.60-0.91) | 3= | 0.98 | 3.7 | (2.5-5.4) | 0.98 | (0.94-0.99) | 2= |
| 10 | MIDI-T1 | T1.SSR | T1 | 0 | 0.62 | 16.5 | (5.7-28.6) | 0.57 | (0.20-0.80) | 15 | 0.83 | 4.6 | (3.1-9.0) | 0.81 | (0.60-0.92) | 20= |
| 11 | MIDI-T1 | T2.SSR | T2 | 0 | 0.87 | 25.9 | (12.3-31.4) | 0.54 | (-0.03-0.81) | 17 | 0.97 | 3.5 | (1.7-5.9) | 0.97 | (0.92-0.99) | 5 |
| 12 | DeepBrainNet | T1 | T1 | 0 | 0.54 | 65.4 | (29.9-81.6) | 0.10 | (-0.07-0.38) | 25 | 0.88 | 3.0 | (1.9-5.3) | 0.87 | (0.72-0.95) | 16= |
| 13 | DeepBrainNet | T1.SSR | T1 | 0 | -0.11 | 23.0 | (16.3-44.5) | -0.09 | (-0.52-0.35) | 26 | -0.08 | 5.6 | (3.3-13.1) | -0.07 | (-0.44-0.35) | 26 |
| 14 | DeepBrainNet | T2.SSR | T1 | 0 | 0.85 | 23.1 | (9.2-42.1) | 0.57 | (0.06-0.82) | 14 | 0.96 | 3.9 | (3.1-5.9) | 0.96 | (0.90-0.98) | 10 |
| 15 | Py_FSv7_ra | T1 | T1 | 0 | 0.66 | 26.2 | (11.9-48.0) | 0.43 | (-0.08-0.75) | 19 | 0.79 | 6.1 | (2.6-15.3) | 0.79 | (0.56-0.91) | 22 |
| 16 | Py_FSv7_ra | T1+T2 | T1 | 5 | 0.71 | 33.7 | (21.8-55.6) | 0.40 | (-0.11-0.75) | 22 | 0.89 | 5.1 | (3.7-7.6) | 0.90 | (0.75-0.96) | 15 |
| 17 | Py_FSv7_ra | T1.SSR | T1 | 0 | 0.74 | 18.0 | (6.9-28.3) | 0.70 | (0.41-0.86) | 10= | 0.87 | 9.6 | (5.3-15.4) | 0.88 | (0.73-0.95) | 18 |
| 18 | Py_FSv7_ra | T2.SSR | T1 | 13 | 0.71 | 9.8 | (4.9-32.5) | 0.67 | (0.30-0.86) | 10= | 0.62 | 13.1 | (6.0-20.2) | 0.63 | (0.13-0.87) | 24 |
| 19 | Py_FSv7_rac | T1 | T1 | 0 | 0.93 | 24.5 | (18.7-29.0) | 0.66 | (-0.06-0.91) | 12= | 0.93 | 7.2 | (2.9-11.2) | 0.93 | (0.85-0.97) | 11= |
| 20 | Py_FSv7_rac | T2 | T2 | 0 | 0.87 | 12.3 | (9.1-16.1) | 0.81 | (0.56-0.92) | 3= | 0.92 | 5.2 | (3.3-8.0) | 0.92 | (0.82-0.97) | 13= |
| 21 | Py_FSv8_ra | T1 | T1 | 2 | 0.89 | 28.9 | (24.0-37.1) | 0.42 | (-0.04-0.80) | 20 | 0.65 | 11.3 | (6.2-14.0) | 0.65 | (0.32-0.84) | 23 |
| 22 | Py_FSv8_ra | T1+T2 | T1 | 4 | 0.73 | 43.4 | (25.8-51.3) | 0.23 | (-0.05-0.62) | 24 | 0.88 | 3.9 | (2.3-6.6) | 0.88 | (0.72-0.95) | 16= |
| 23 | Py_FSv8_rac | T1 | T1 | 0 | 0.92 | 24.4 | (16.2-30.5) | 0.66 | (-0.07-0.91) | 12= | 0.92 | 7.2 | (3.0-11.2) | 0.92 | (0.81-0.97) | 13= |
| 24 | Py_FSv8_rac | T1.SSR | T1 | 0 | 0.87 | 13.0 | (7.5-17.7) | 0.81 | (0.50-0.92) | 5 | 0.93 | 4.3 | (2.2-7.4) | 0.93 | (0.85-0.97) | 11= |
| 25 | Py_FSv8_rac | T2 | T2 | 0 | 0.88 | 20.5 | (6.3-27.0) | 0.72 | (-0.04-0.91) | 6 | 0.97 | 5.5 | (2.3-7.1) | 0.97 | (0.93-0.99) | 8 |
| 26 | Py_FSv8_rac | T2.SSR | T2 | 0 | 0.93 | 12.2 | (4.2-14.4) | 0.89 | (0.55-0.96) | 2 | 0.98 | 4.2 | (1.8-5.8) | 0.97 | (0.94-0.99) | 5 |

Performance metrics varied widely. For validity, the HF metrics (representing the benchmark) showed numerically better performance and tighter ranges compared to ULF (HF: *r* = 0.74–0.96, *R*^2^ = 0.17–0.81, MAE = 4.99–12.02 years; ULF: *r* = 0.12–0.92, *R*^2^ = 1.26–0.64, MAE = 6.7–19.27 years). For HF-ULF correspondence, the ranges were *r* = − 0.11–0.93, ASPD = 7.6–65.4, ICC = − 0.09–0.92, and for test-retest reliability *r* = − 0.08–0.99, ASPD = 1.7–13.1, ICC = − 0.07–0.99. The top four brain-age model combinations, selected based on validity rankings, were SynthBA on T2 scans, MIDI on T2 scans, PyBrainAge using `recon-all- clinical` on T1 scans with SynthSR, and BrainageR on T1 scans with SynthSR, in descending rank. For these four combinations, validity plots are shown in Figure 2, and correspondence, test-retest reliability and Bland–Altman plots are shown in Figure 3. For a complete set of plots, see SI section 5.

**Figure 2.**
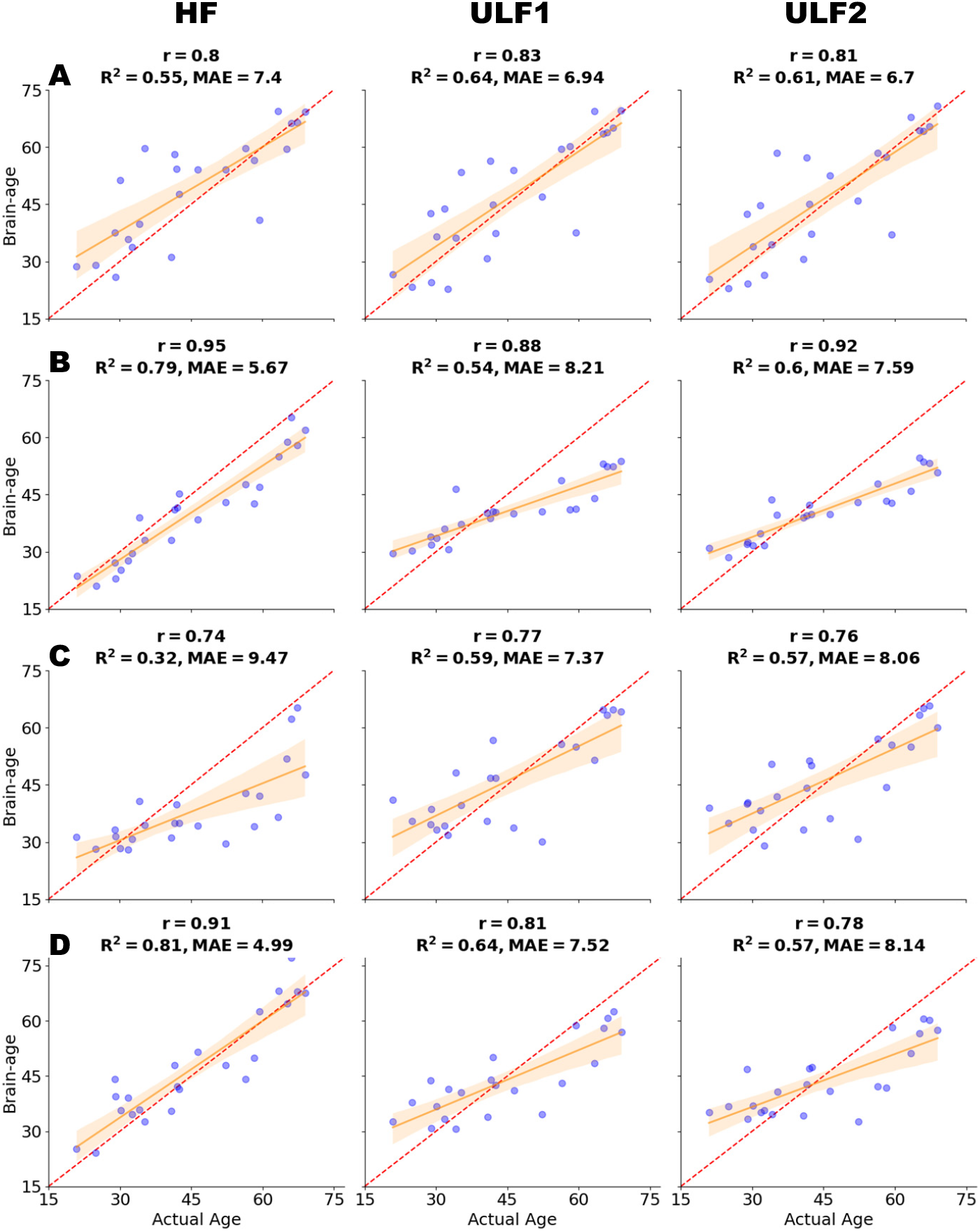
Validity plots for the top four performing brain-age models. Rows (A–D) show model/scan combinations: (A) SynthBA on T2; (B) MIDI (T2 model) on T2; (C) PyBrainAge on T1 SSR (FreeSurfer v8, `recon-all-clinical`); and (D) BrainageR on T1 SSR. Columns are HF (high-field), ULF1, and ULF2 (ultra-low-field sites 1/2). Each panel plots estimated brain age versus actual (chronological) age; the red dashed diagonal is the identity line. The solid line is the fitted regression with a confidence interval. Annotations report the Pearson correlation *r*, the coefficient of determination *R*^2^, and the mean absolute error (MAE).

**Figure 3.**
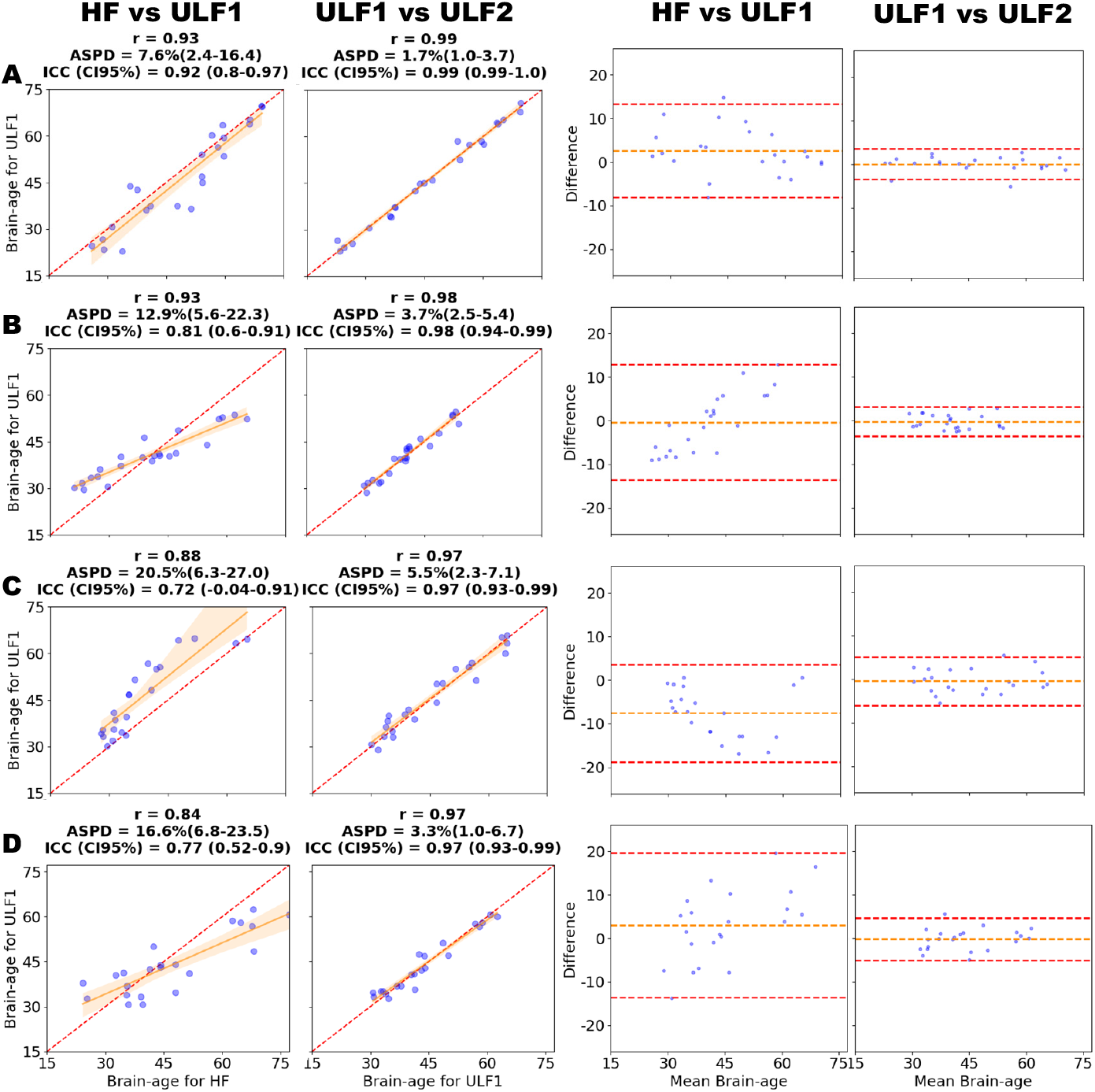
Correspondence and reliability plots for the top four performing brain-age models. Rows (A–D) show model and scan type combinations: (A) SynthBA on T2; (B) MIDI (T2 model) on T2; (C) PyBrainAge on T1 SSR (FreeSurfer v8, `recon-all-clinical`); and (D) BrainageR on T1 - SSR. Columns are: (1) Correspondence (HF vs. ULF1), (2) test-retest reliability (ULF1 vs. ULF2), (3) Bland–Altman for HF vs. ULF1, and (4) Bland–Altman for ULF1 vs. ULF2. Correspondence panels (columns 1–2) show scatter plots comparing estimated brain-age between scanner pairs; annotations report the Pearson correlation *r, R*^2^, absolute symmetric percent difference (ASPD) with 95% confidence interval (CI), and intraclass correlation coefficient (ICC) with 95% CI. Bland– Altman panels (columns 3–4) plot the pairwise difference versus the pairwise mean; the central orange dashed line indicates the mean bias and the outer dashed red lines show the 95% limits of agreement. Abbreviations: HF = high-field; ULF1/ULF2 = ultra-low-field sites 1/2; SSR = SynthSR.

For validity and correspondence plots, perfect performance would be indicated if datapoints aligned diagonally along the line of identity. Whilst the baseline validity performance of HF approached this pattern closely for the top four performing model combinations (*r* = 0.74–0.95, *R*^2^ = 0.32–0.81, MAE = 4.99–9.47 years), both ULF1 and ULF2 plots showed a broadly similar pattern, albeit with estimates falling somewhat further from the line of identity (*r* = 0.76–0.92, *R*^2^ = 0.54–0.64, MAE = 6.7–8.21 years).

Correspondence showed good results, with strong performance for SynthBA (*r* = 0.93, ASPD = 7.6%; 95% CI: 2.4–16.4; ICC = 0.92, 95% CI: 0.80–0.97) and moderate-to-strong for the other three top-performing models (*r* = 0.84–0.93, ASPD = 12.9–20.5%, ICC = 0.72–0.81). However, for PyBrainAge the ICC estimate was 0.72, but the 95% CI included zero (− 0.04–0.91), indicating non-significance. For SynthBA, the Bland–Altman plot did not reveal any obvious age-related bias. In contrast, the plots for MIDI and BrainageR indicated that ULF scans tended to underestimate older ages and overestimate younger ages relative to HF, while PyBrainAge showed a systematic overestimation relative to HF. Test-retest reliability for the top-performing model combinations was strong (*r* = 0.97–0.99, ASPD = 1.7–5.5%, ICC = 0.97–0.99), and Bland–Altman plots did not reveal any noticeable age-related bias. The complete correspondence and reliability results, including HF versus ULF2, are provided in SI section 4.

The best-performing model combination based on validity rankings was SynthBA on T2 scans, which also ranked first in both correspondence and test-retest reliability rankings. As the best-performing model, SynthBA was further tested on singleacquisition scans; these results are shown in Table 4 and the plots can be found in SI section 5.2.1. Validity metrics were weak-to-strong *r* = 0.65–0.83, *R*^2^ = − 0.70– 0.62, MAE = 6.–16.74 years, while test-retest reliability was strong (*r* = 0.86–0.99, ASPD = 1.7–6.7%, ICC = 0.86–0.99).

**Table 4:**
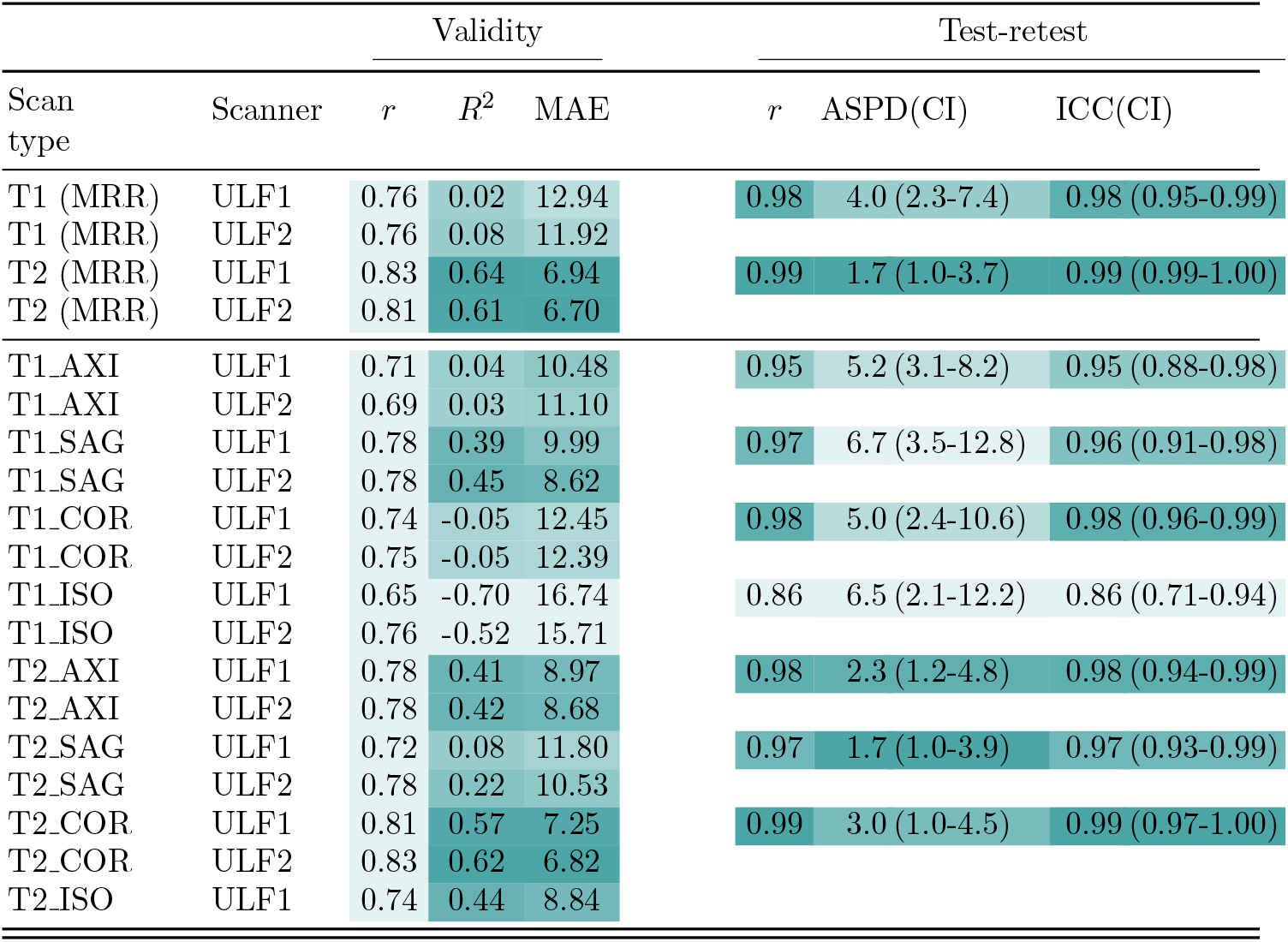
Best-performing model: SynthBA. The first four rows recapitulate the validity and testretest reliability results for SynthBA using T1 and T2 scans (MRR, without SSR), for reference. The remaining rows are hitherto unreported results: the performance metrics for the single acquisition ULF scans (no MRR/SSR) using SynthBA. Abbreviations: HF, high-field; ULF1/ULF2, ultra-low-field at site 1/site 2; SSR, SynthSR; *r*, Pearson correlation;*R*^2^: coefficient of determination; MAE: mean absolute error; ASPD, absolute symmetric percent difference; ICC, intraclass correlation coefficient; CI, confidence interval.

| Scan type | Scanner | Validity |  |  | Test-retest |  |  |
| --- | --- | --- | --- | --- | --- | --- | --- |
| | | $r$ | $R^2$ | MAE | $r$ | ASPD(CI) | ICC(CI) |
| T1 (MRR) | ULF1 | 0.76 | 0.02 | 12.94 | 0.98 | 4.0 (2.3-7.4) | 0.98 (0.95-0.99) |
| T1 (MRR) | ULF2 | 0.76 | 0.08 | 11.92 |  |  |  |
| T2 (MRR) | ULF1 | 0.83 | 0.64 | 6.94 | 0.99 | 1.7 (1.0-3.7) | 0.99 (0.99-1.00) |
| T2 (MRR) | ULF2 | 0.81 | 0.61 | 6.70 |  |  |  |
| T1_AXI | ULF1 | 0.71 | 0.04 | 10.48 | 0.95 | 5.2 (3.1-8.2) | 0.95 (0.88-0.98) |
| T1_AXI | ULF2 | 0.69 | 0.03 | 11.10 |  |  |  |
| T1_SAG | ULF1 | 0.78 | 0.39 | 9.99 | 0.97 | 6.7 (3.5-12.8) | 0.96 (0.91-0.98) |
| T1_SAG | ULF2 | 0.78 | 0.45 | 8.62 |  |  |  |
| T1_COR | ULF1 | 0.74 | -0.05 | 12.45 | 0.98 | 5.0 (2.4-10.6) | 0.98 (0.96-0.99) |
| T1_COR | ULF2 | 0.75 | -0.05 | 12.39 |  |  |  |
| T1_ISO | ULF1 | 0.65 | -0.70 | 16.74 | 0.86 | 6.5 (2.1-12.2) | 0.86 (0.71-0.94) |
| T1_ISO | ULF2 | 0.76 | -0.52 | 15.71 |  |  |  |
| T2_AXI | ULF1 | 0.78 | 0.41 | 8.97 | 0.98 | 2.3 (1.2-4.8) | 0.98 (0.94-0.99) |
| T2_AXI | ULF2 | 0.78 | 0.42 | 8.68 |  |  |  |
| T2_SAG | ULF1 | 0.72 | 0.08 | 11.80 | 0.97 | 1.7 (1.0-3.9) | 0.97 (0.93-0.99) |
| T2_SAG | ULF2 | 0.78 | 0.22 | 10.53 |  |  |  |
| T2_COR | ULF1 | 0.81 | 0.57 | 7.25 | 0.99 | 3.0 (1.0-4.5) | 0.99 (0.97-1.00) |
| T2_COR | ULF2 | 0.83 | 0.62 | 6.82 |  |  |  |
| T2_ISO | ULF1 | 0.74 | 0.44 | 8.84 |  |  |  |

### 3.1. A closer look at PyBrainAge and the impact of FreeSurfer variations

Within the PyBrainAge iterations, three additional findings were observed. First, the FreeSurfer software version (v7.4.0 vs. v8.0.0) appeared to influence results. Using the standard `recon-all` command, v8.0.0 gave higher validity rankings than v7.4.0 (e.g., with T1 scans in ULF1, *R*^2^ = − 0.01 [rank 20] vs. *R*^2^ = 0.29 [rank 14]). By contrast, version differences seemed to have less impact when using the `reconall-clinical` command, which was introduced in v7.4.0 (e.g., *R*^2^ = 0.56 [rank 6] in v7.4.0 vs. *R*^2^ = 0.54 [rank 8] in v8.0.0). Second, the use of `recon-all-clinical` instead of `recon-all`, or SynthSR instead of no treatment, improved ULF brain-age estimation. For example, PyBrainAge with `recon-all` and T1 in ULF1 achieved *R*^2^ = − 0.01 (rank 20), improving to *R*^2^ = 0.56 (rank 6) with `recon-all-clinical` and *R*^2^ = 0.45 (rank 7) with SynthSR. The best performance was achieved when both were applied in combination (*R*^2^ = 0.59, rank 3, v8.0.0). Third, adding T2 to aid T1 segmentation (using the `-T2` and `-T2-pial` flags) unexpectedly reduced performance (e.g., rank 20 dropped to rank 25 in v7.4.0; rank 14 dropped to rank 19 in v8.0.0).

## 4. Discussion

In this study, we tested the validity, correspondence and test-retest reliability of brain-age estimates derived from ULF MRI, using five diverse brain-age models and a range of preprocessing pipelines. This design resulted in an extensive set of combinations, allowing us to benchmark performance systematically.

Our key finding is that ULF brain-age estimates can achieve high validity and reliability from T1 or T2 scans, comparable to HF benchmarks. To illustrate this, we highlight the top four performing model combinations, which capture broader patterns observed across the full set of pipelines. These were selected based on validity rankings and consisted of (in descending rank order): SynthBA on T2 scans, MIDI on T2 scans, PyBrainAge using `recon-all-clinical` on T1 scans with SynthSR, and BrainageR on T1 scans with SynthSR. This convergence across evaluation axes is notable: validity showed that ULF estimates closely tracked actual (chronological) age (with *r* and *R*^2^ values ranging from 0.77 to 0.88 and 0.54 to 0.64, respectively), correspondence demonstrated strong agreement with HF estimates (ICC ranging from 0.72 to 0.92), and test-retest reliability confirmed that results were highly reproducible across two ULF scanners (ICC ranging from 0.97 to 0.99), supporting the robustness of repeated acquisitions in ULF for this scanner and consistent with prior reports using the same dataset (Váša et al., 2025). Together, this highlights the potential for ULF-derived brain-age estimates to be both accurate and reliable, a critical prerequisite for use as a biomarker in research and clinical settings.

Beyond their strong performance, findings from the top four combinations revealed other patterns. The top models were based on a range of different input data types: raw scans (MIDI, SynthBA), voxel-wise tissue volumes (BrainageR), or morphometric parcellations (PyBrainAge, which uses FreeSurfer segmentations). This suggests that robust features important for brain-age estimation in ULF can be extracted at both fine and coarse scales, aligning with previous ULF fidelity research showing moderately strong HF-to-ULF correspondence for both coarse parcellations (Váša et al., 2025) and fine-grained structures at the vertex level (Pretzsch et al., 2025). Another systematic feature was a consistent negative bias relative to the identity line (Figure 2), reflected in shallower regression slopes and consistent with the known regression-to-the-mean effect (de Lange and Cole, 2020), whereby predictions are biased toward the mean age of the training sample.

Another key finding in this study was that the precise combination of preprocessing, brain-age model and scan type is crucial for adequate brain-age performance in ULF scans, with performance varying considerably across pipelines. This is consistent with recent evidence also showing that the accuracy of ULF-derived anatomical features depends on the processing used (Pretzsch et al., 2025). For example, the application of SynthSR to T1 improved validity ranks from 24th to 4th in BrainageR and from 20th to 7th in PyBrainAge v7.4.0 with `recon-all`. However, in some cases the gains were minimal and in others even detrimental. For example, the two highest validity rankings were SynthBA and MIDI on T2 scans without SynthSR, where their treatment with SynthSR actually reduced performance (ranks reduced from 1st to 13th and 2nd to 23rd, respectively). In addition, in SynthBA, some anisotropic acquisitions performed closely to the MRR-processed isotropic scans (constructed from three orthogonal anisotropic acquisitions; e.g., T2 (MRR): *r* = 0.81, *R*^2^ = 0.61, MAE = 6.70 vs. T2 COR: *r* = 0.83, *R*^2^ = 0.62, MAE = 6.82), further indicating that MRR processing, another form of super-resolution, may not be essential for certain brain-age models. By contrast, extensive image enhancement benefited PyBrainAge, which performed best when combining MRR, SSR, and the `recon-all-clinical` pipeline (incorporating SynthSeg), compared to the classical `recon-all` command. Scan modality also made a difference, whereby each model showed distinct strengths and weaknesses which could not always be readily explained by the modality native to its training. For instance, SynthBA (training included both T1 and T2 scans) and MIDI (trained separately for T1 and T2) excelled with T2 but were weaker with T1 (validity ranks reduced from 1st to 18th and 2nd to 16th, respectively). In contrast, a strong and balanced performance across T1 and T2 modalities was observed for BrainageR once SynthSR was applied (validity rankings of 4th and 5th for T1 SSR and T2 SSR, respectively). SynthSR effectively converts a T2 scan to a T1 contrast which is native to the BrainageR training data.

These findings highlight three key points. First, they suggest that models trained on variable quality HF datasets (e.g., including noisy, lower-quality or synthetic scans, as in MIDI and SynthBA) perform robustly even without SynthSR. By contrast, models trained on datasets that are diverse in site or study but still composed mainly of research-grade scans (e.g., BrainageR, PyBrainAge, DBN) appear more reliant on SynthSR for adequate performance with ULF data. However, confirming this trend would require a dedicated investigation beyond the scope of this study. Second, they suggest that image enhancement techniques like SynthSR warrant careful consideration, as different models may exhibit varying sensitivities, and additional processing can sometimes yield diminishing or even negative returns. Third, they further suggest that robust brain-age estimates are attainable even without acquisition of three orthogonal anisotropic scans, and subsequent preprocessing using MRR, as observed with certain single-acquisition results. Clinically, this is advantageous because physicians often prefer sparse acquisitions in orthogonal planes, which are faster to inspect (Billot et al., 2023b). Beyond workflow, multiple shorter anisotropic acquisitions may also reduce the risk of motion artefacts compared with a single longer high-resolution scan (Baljer et al., 2025b,a), an advantage that may be important in populations for whom lying still is challenging, such as children (Scheinost et al., 2019). Furthermore, our results suggest that even within single-slice acquisitions, some orientations may be preferable: with SynthBA, sagittal slices performed best for T1 (*R*^2^ = 0.45 vs. *R*^2^ = 0.03 axial, *R*^2^ =− 0.05 coronal in ULF2), while coronal slices performed best for T2 (*R*^2^ = 0.62 vs. *R*^2^ = 0.42 axial, *R*^2^ = 0.22 sagittal in ULF2). Although additional benchmarking is needed, such orientation-specific differences could help guide protocol design when only limited acquisitions are feasible.

Consistent with this broader pattern, an in-depth look at findings from PyBrainAge - which relies on FreeSurfer-derived morphometric features - showed that the choice of preprocessing strongly influenced outcomes. For example, both the FreeSurfer version (v7.4 vs v8.0) and command type (`recon-all` vs. `recon-all- clinical`) affected brain-age performance. Such variability is consistent with previous reports of segmentation differences across FreeSurfer versions (Whelan et al., 2016) and further underscores how preprocessing choices can shape ULF brain-age estimation, an effect that has also been observed in other imaging studies (Haller et al., 2016; Botvinik-Nezer et al., 2020; Sederevičius et al., 2021; Pretzsch et al., 2025). It is also worth bearing in mind that FreeSurfer-based processing substantially increases computational time compared with the other brain-age models tested here, which is an important practical consideration for future applications.

### 4.1. Limitations and future work

This study has several limitations. First, the sample size was modest, so chance findings, for example in the form of unstable pipeline rankings and their differences, cannot be excluded. However, although modest sample size limits precision, the repeated-measures design ensured that all comparisons were made within the same individuals, reducing between-subject variability. This increases confidence that observed differences across pipelines reflect genuine methodological sensitivities rather than noise, especially as validity was assessed against chronological age (the ground truth) and correspondence to HF benchmarks. In addition, the fact that ICC confidence intervals rarely crossed zero further supports the presence of a reliable signal. On a related note, the comparisons described in the discussion are based on rankings rather than formal statistical tests. This reflects the exploratory nature of this study where our aim was not to test specific hypotheses but rather assess the general feasibility of brain-age estimation in ULF via systematic evaluation of a landscape of possible model-pipeline-scan combinations.

Second, the possibility of hallucinations, whereby image-enhancement methods introduce spurious anatomical features, cannot be excluded; this has been noted as a general concern for super-resolution approaches (Lin et al., 2023), and may also apply to SynthSR. Notably, SynthSR is explicitly designed to inpaint lesions (including white matter hyperintensities), which could pose problems when applied in older individuals or patient populations. However, the strong performance of pipelines that do not rely on SynthSR (e.g., MIDI with T2) suggests this is unlikely to be the sole explanation of our robust ULF brain-age findings. Nevertheless, emerging approaches such as SuperSynth^11^now offer super-resolution without lesion inpainting.

A further limitation concerns generalisability. The sample, was modestly sized and non-randomly selected, which raises the possibility of bias in the cohort. In addition, although we refer to the scanners as HF and ULF, the results may not extend to other scanner models or field strengths or acquisition parameters (Pretzsch et al., 2025). Finally, while we tested five established brain-age models across a rich combination of pipelines, our analyses were not exhaustive and other widely used frameworks were not included.

These considerations point to aims for future work. A key next step will be to extend this work by testing whether ULF-derived brain-age estimates can serve as predictors of health outcomes, thereby establishing their translational validity. In addition, replication in larger samples, across different scanners, sequence parameters and scanner software variants will be important to assess generalisability. Finally, while it remains to be determined whether brain-age models trained directly on ULF data may improve performance, this is complicated by the rapid evolution of ULF software. Frequent scanner updates and protocol changes create moving targets that may widen the domain gap, highlighting the need for ongoing benchmarking or the development of approaches that are agnostic to such shifts.

## 5. Conclusion

To our knowledge, this is the first study to investigate brain-age estimation using ULF MRI. By systematically evaluating multiple pipelines, we demonstrate both the promise and the challenges of extending brain-age modelling to this new imaging domain. Our results show that ULF can deliver accurate and reliable estimates that closely approach ground truth and HF benchmarks, highlighting brain-age as a feasible biomarker for ULF applications. These findings are consistent with recent studies demonstrating that ULF-derived biomarkers can perform comparably to HF, including in dementia (Sorby-Adams et al., 2024), multiple sclerosis (Arnold et al., 2022), healthy morphometry (Váša et al., 2025), and normative model centiles (see supplement in Dorfschmidt et al. (2025)). Importantly, we also show that preprocessing choices influence outcomes, underscoring the need for careful benchmarking in future applications. Establishing robustness and clinical utility will require validation in larger and more diverse populations, and in clinical contexts. In this regard, brain-age has the potential to be valuable not only in clinical settings but also as a predictor of health outcomes in healthy cohorts (see also Dorfschmidt et al. (2025)). If confirmed more widely, ULF brain-age estimation could provide a practical and scalable tool for clinical decision-making, population research, and long-term patient monitoring, ultimately helping to make advanced neuroimaging biomarkers more accessible worldwide.

## Appendix A. Glossary

ANTs: Advanced Normalization Tools
ASPD: Absolute Symmetric Percent Difference
AXI: Axial
COR: Coronal
CSF: Cerebrospinal Fluid
DBN: DeepBrainNet
FLAIR: Fluid-Attenuated Inversion Recovery
FSQC: FreeSurfer Quality Control
HF: High-Field MRI
ICC: Intraclass Correlation Coefficient
ISO: Isotropic
MAE: Mean Absolute Error
MNI: Montreal Neurological Institute
MRR: Multi-Resolution Registration
MRI: Magnetic Resonance Imaging
MPRAGE: Magnetisation Prepared Rapid Gradient Echo
QC: Quality Control
SAG: Sagittal
SDF: Signed-Distance Function
SNR: Signal-to-Noise Ratio
SPM: Statistical Parametric Mapping
SSR: SynthSR
T1: T1-weighted MRI
T2: T2-weighted MRI
ULF: Ultra-Low-Field MRI

## Acknowledgements

This study was supported by a National Institute for Health and Care Research (NIHR) Biomedical Research Centre Early Career Investigator Award to F.V., and by the Bill and Melinda Gates Foundation UNITY and other BMGF projects [INV-032788; INV-047888; INV-005774 and INV-047885]. TCB and DW were supported by the UK Medical Research Council (MR/W021684/1) and by core funding from the Wellcome/EPSRC Centre for Medical Engineering (WT203148/A/16/Z). SAM was supported by the Engineering and Physical Sciences Research Council (EPSRC) [grant number EP/W524335/1]; the EPSRC-funded Centre for Doctoral Training in Intelligent, Integrated Imaging in Healthcare (i4health) (EP/S021930/1); and the Department of Health’s NIHR-funded Biomedical Research Centre at University College London Hospitals. JEI was supported by NIH grants 1R01AG070988, 1R01EB031114, 1UM1MH130981, 1RF1AG080371, 1R21NS138995.

## CRediT Author Statement

**Francesca Biondo** Conceptualisation, Data Curation, Formal Analysis, Investigation, Methodology, Project Administration, Visualisation, Writing–Original Draft.

**Carly Bennalick**: Data Curation, Formal Analysis, Investigation, Methodology

**Sophie Martin**: Methodology, Writing–Review & Editing

**Lemuel Puglisi**: Methodology, Writing–Review & Editing

**Thomas C. Booth**: Writing–Review & Editing

**David A. Wood**: Writing–Review & Editing

**Juan Eugenio Iglesias**: Writing–Review & Editing

**František Váša**: Conceptualisation, Data Curation, Formal Analysis, Supervision, Funding Acquisition, Project Administration, Writing–Review & Editing.

**James H. Cole**: Conceptualisation, Supervision, Project Administration, Writing– Review & Editing.

## Conflict of interest disclosure

All authors declare no competing interests except: JHC is shareholder/advisor for BrainKey and Claritas HealthTech; LP works for Queen Square Analytics, University College London.

## Ethics approval

*The original study via which the data were collected was approved by the King’s College London human Research Ethics Committee (Reference: HR/DP-21/22-26481)*.

## Data Availability

Data are publicly available except for MRR, which will be released soon. https://openneuro.org/datasets/ds006557

## Footnotes

1 Thttps://www.nitrc.org/projects/mricro

2 https://github.com/james-cole/brainageR

3 https://doi.org/10.5281/zenodo.3476365

4 https://github.com/LemuelPuglisi/SynthBA

5 https://github.com/vishnubashyam/DeepBrainNet

6 https://github.com/MIDIconsortium/BrainAge

7 https://github.com/james-cole/PyBrainAge

8 https://surfer.nmr.mgh.harvard.edu/fswiki/FreeSurferWiki

^9^https://surfer.nmr.mgh.harvard.edu/fswiki/recon-all-clinical

10 https://deep-mi.org/fsqc/dev/

11 https://surfer.nmr.mgh.harvard.edu/fswiki/SuperSynth

## Notes

### Author Declarations

The original study via which the data were collected was approved by the King's College London human Research Ethics Committee (Reference: HR/DP-21/22-26481).

