## Supplementary Information for "Brain-age in ultra-low-field MRI: how well does it work?"

### 1 MRI scan types

The table below lists all MRI scan and scanner combinations. All scans were tested using all five brain-age models in the study, except for the single acquisition scans which were used with the best-performing brain-age model, only. All scan conditions had $n = 23$ scans except for the ULF1 T1 derived scans which had $n = 22$. For ULF2 there was no T2_ISO sequence available.

Table S1: Scan types and sample sizes

| Scanner group | Scan type | Processing (MRR / SynthSR) | # scans (n) |
|---|---|---|---|
| **High-field (3 T)** | | | |
| HF | T1 | – / – | 23 |
| | T2 | – / – | 23 |
| **Ultra-low-field with MRR (64 mT)** | | | |
| ULF1 | T1 | MRR / – | 22 |
| | T2 | MRR / – | 23 |
| ULF2 | T1 | MRR / – | 23 |
| | T2 | MRR / – | 23 |
| ULF1 | T1_SSR | MRR / SynthSR | 22 |
| | T2_SSR | MRR / SynthSR | 23 |
| ULF2 | T1_SSR | MRR / SynthSR | 23 |
| | T2_SSR | MRR / SynthSR | 23 |
| **Ultra-low-field single acquisition (64 mT)** | | | |
| ULF1 | T1_AXI | – / – | 23 |
| | T2_AXI | – / – | 23 |
| | T1_SAG | – / – | 23 |
| | T2_SAG | – / – | 23 |
| | T1_COR | – / – | 23 |
| | T2_COR | – / – | 23 |
| | T1_ISO | – / – | 23 |
| | T2_ISO | – / – | 23 |
| ULF2 | T1_AXI | – / – | 23 |
| | T2_AXI | – / – | 23 |
| | T1_SAG | – / – | 23 |
| | T2_SAG | – / – | 23 |
| | T1_COR | – / – | 23 |
| | T2_COR | – / – | 23 |
| | T1_ISO | – / – | 23 |

HF: high-field; ULF1/ULF2: ultra-low-field at site 1/site 2; MRR: multi-resolution registration; SSR: SynthSR; AXI: axial; SAG: sagittal; COR: coronal; ISO: isotropic.

### 2 Quality Control

#### 2.1 SynthSR QC: examples

Representative examples of raw ULF scans and their SynthSR outputs are included (Figure S1). Visual QC was performed in MRIcro.[1]

[1] https://www.nitrc.org/projects/mricro

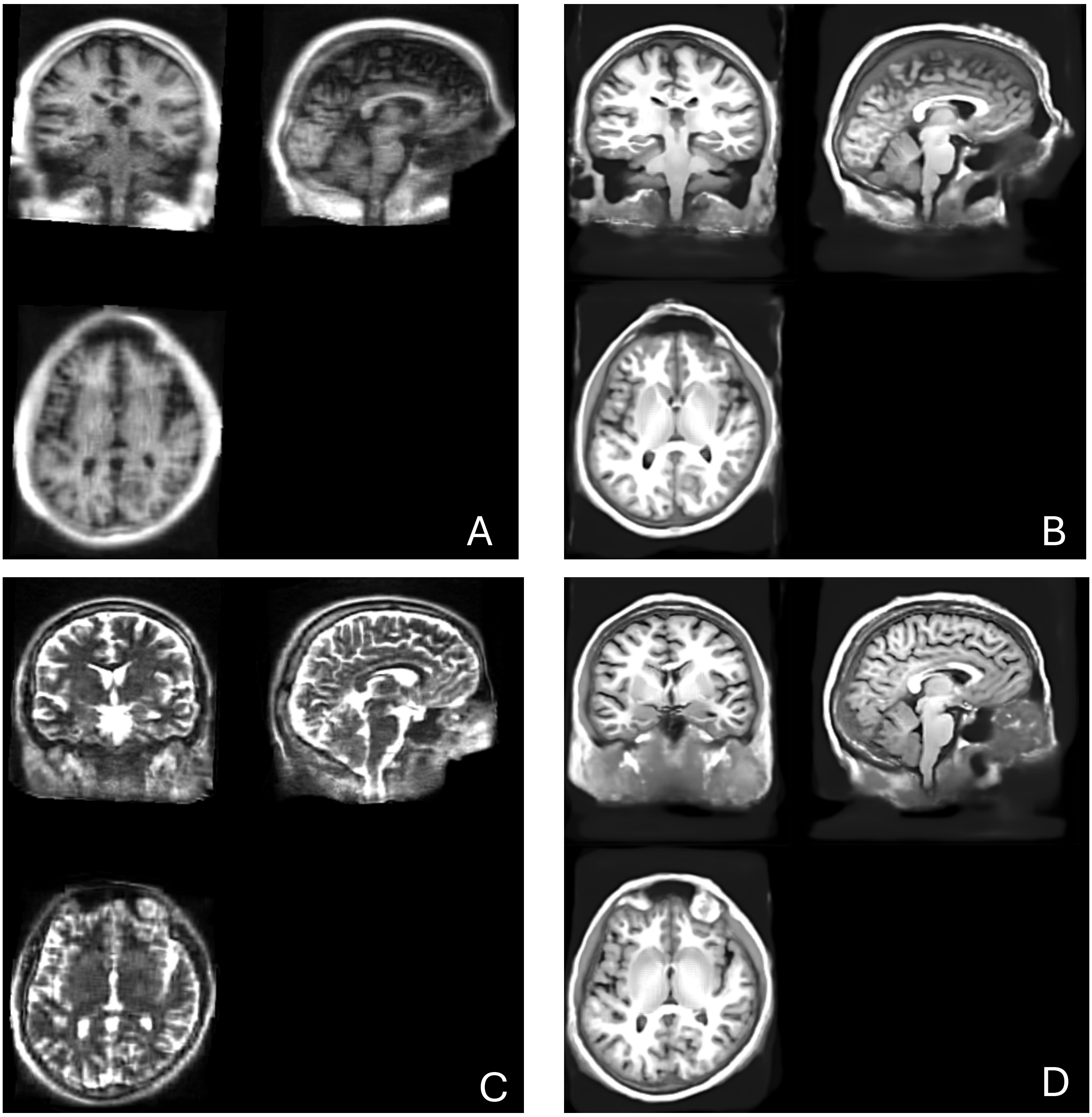

Figure S1: MRIcro screenshots illustrating SynthSR on ULF MRR data (same participant). (A) ULF1 T1; (B) ULF1 T1_SSR (T1 after SynthSR); (C) ULF1 T2; (D) ULF1 T2_SSR (T2 after SynthSR).

Applying SynthSR to high-field (HF) images is, in principle, unnecessary given that SynthSR is intended for heterogeneous clinical scans. Yet we tested it for completeness. Applying SynthSR to HF images produced a visibly degraded appearance (e.g., blurring/artefacts) particularly for T1-weighted images. These outputs were *not* used in any analyses; examples are included for reference only (Figure S2).

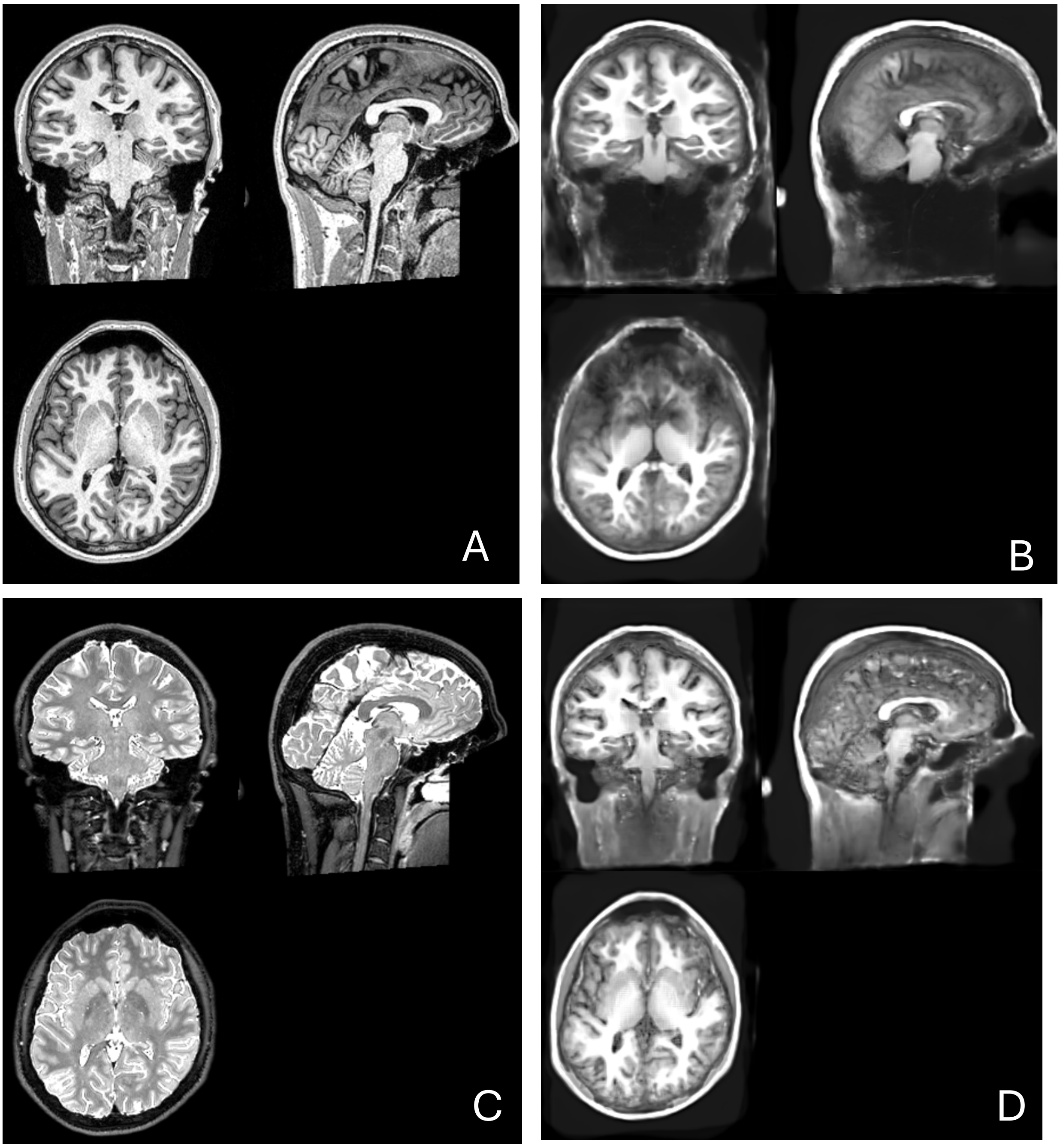

Figure S2: MRIcro screenshots illustrating SynthSR on HF data (same participant). (A) HF T1; (B) HF T1 after SynthSR; (C) HF T2; (D) HF T2 after SynthSR. HF SynthSR outputs were *not* used in any analyses.

#### 2.2 PyBrainAge QC: FreeSurfer `recon-all`

The FSQC toolbox[2] was used to assist visual quality control of `recon-all` outputs. For each participant, FSQC screenshot panels were reviewed to inspect FreeSurfer segmentations and skull-stripping results.

Quantitatively, FreeSurfer's surface topology statistics (e.g., number of surface holes/defects per hemisphere) were inspected. These were used trigger targeted re-reviews: scans with a high number of defects were double-checked visually. FSQC was applied only to `recon-all` outputs.

[2] `https://deep-mi.org/fsqc/dev/`

###### 2.2.1 Examples

In this section, typical QC screenshots for `recon-all` (FreeSurfer v7.4.0) are shown.

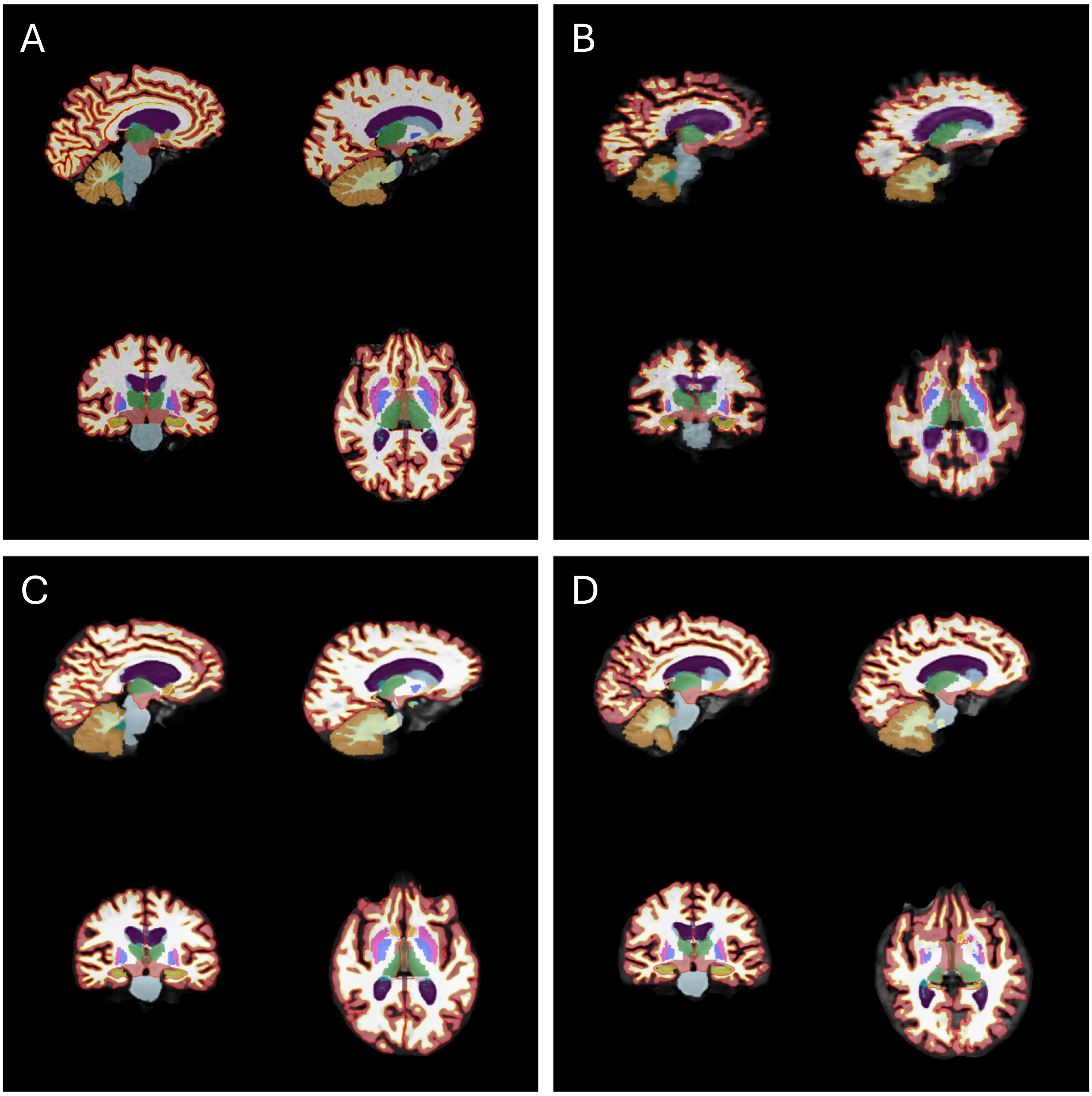

Figure S3: Quality control screenshots for `recon-all` outputs. Panel A shows HF T1; Panel B shows ULF T1; Panel C shows ULF T1_SSR and Panel D shows ULF T2_SSR. All from the same participant.

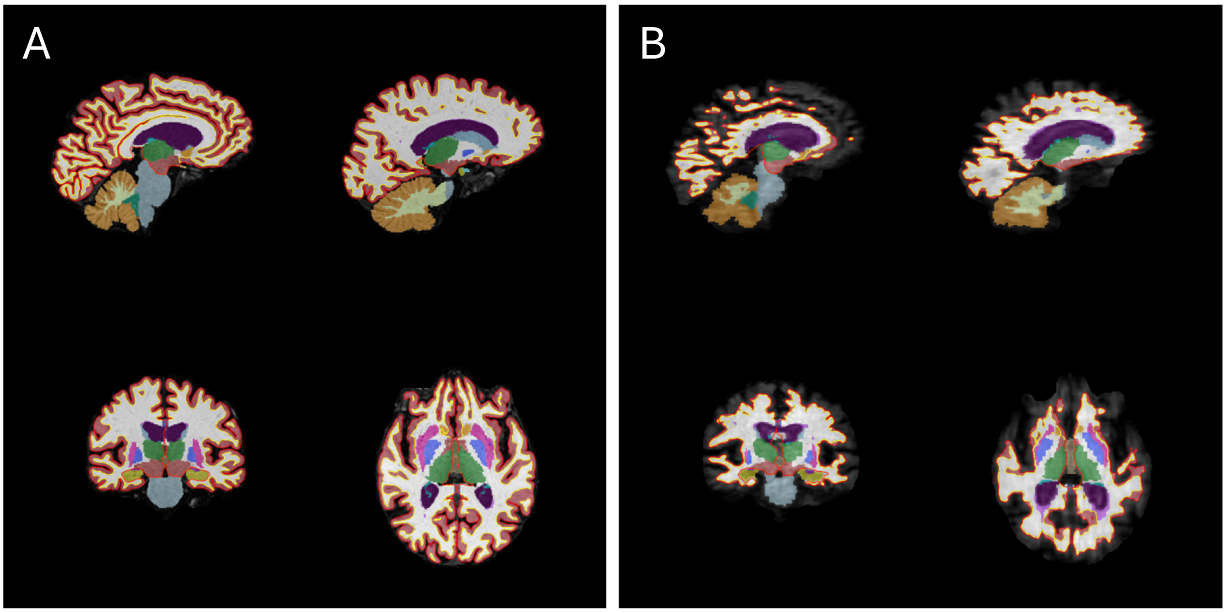

Figure S4: Quality control screenshots for `recon-all` outputs using the T1+T2 setting. Panel A shows HF T1+T2; Panel B shows ULF T1+T2; All from the same participant.

All scans shown in Figure S3 and Figure S4 above passed QC. Clearly, some segmentations appear suboptimal (e.g. erosion/missingness at the edges Figure S4B); this was expected for native ULF data and reflects a deliberate trade-off given the study's focus on feasibility and reliability at ultra-low field. To avoid excluding the entire set of ULF scans, a more permissive QC threshold was applied for ULF compared to HF and only scans that were clear outliers within each ULF batch were removed.

#### 2.3 PyBrainAge QC: FreeSurfer `recon-all-clinical`

At the time of analysis, FSQC was not compatible with `recon-all-clinical` outputs. Hence, we created a lightweight custom toolbox to support visual QC of `recon-all-clinical` outputs. For each scan, the toolbox loads the SynthSeg label map (`synthseg.mgz`) and overlays it on the normalised image (`synthSR.norm.mgz`). It then exports a single composite panel comprising nine slices – three sagittal, three axial, and three coronal – for rapid inspection of segmentation.

Five scans were visually inspected per `recon-all-clinical` batch (see batch list below). These were selected at random. A further five scans were visually inspected. These were the five poorest cases according to the quantitative QC (lowest SynthSeg QC scores; see below). Therefore, in total, 10 of 23 scans per batch (or 10 of 22 for ULF1 T1) were visually reviewed. No scans were excluded on the basis of this review. This hybrid approach provided broad coverage while also targeting the most problematic cases identified by the quantitative QC (see below).

Table S2: QC batches for `recon-all-clinical`

| Scanner group | Sequence | Abbrev. | v7.4 | v8.0 |
|---|---|---|---|---|
| HF | T1 | HF T1 | X | X |
| HF | T2 | HF T2 | X | X |
| ULF1 | T1 | ULF1 T1 | X | X |
| ULF1 | T2 | ULF1 T2 | X | X |
| ULF2 | T1 | ULF2 T1 | X | X |
| ULF2 | T2 | ULF2 T2 | X | X |
| ULF1 | T1_SSR | ULF1 T1_SSR | – | X |
| ULF1 | T2_SSR | ULF1 T2_SSR | – | X |
| ULF2 | T1_SSR | ULF2 T1_SSR | – | X |
| ULF2 | T2_SSR | ULF2 T2_SSR | – | X |

**Notes:** "v7.4" denotes FreeSurfer v7.4.0; "v8.0" denotes FreeSurfer v8.0.0. SSR indicates SynthSR-derived inputs. HF = high-field; ULF1/ULF2 = ultra-low-field scanner site 1/2. An "X" indicates a separate batch (i.e. total 16 batches); "–" indicates not applicable / not run.

For quantitative QC, the `synthseg.qc.csv` file generated by the `recon-all-clinical` pipeline was used. This file is directly generated by the automated quality control (QC) module of SynthSeg and reports scores (0–1) reflecting the quality of SynthSeg segmentation for eight summary labels (general grey matter, general white matter, CSF, cerebellum, brainstem, thalamus, putamen+pallidum, hippocampus+amygdala). In more detail, after the input MRI is segmented, the resulting label map is passed to a CNN-based regressor, which outputs QC scores between 0 and 1. These scores are designed to approximate the Dice values that would have been obtained if manual ground truth were available, thereby enabling unreliable segmentations to be flagged or excluded without manual inspection (Billot et al., 2023a,b). The eight Dice scores were aggregated per scan into a composite "worst" score by ranking scans across the eight features and identifying the five lowest-scoring scans within each batch. These were then visually inspected (see description above), alongside the random sample.

##### 2.3.1 Examples

Shown here are typical QC screenshots for `recon-all-clinical` (FreeSurfer v8.0.0).

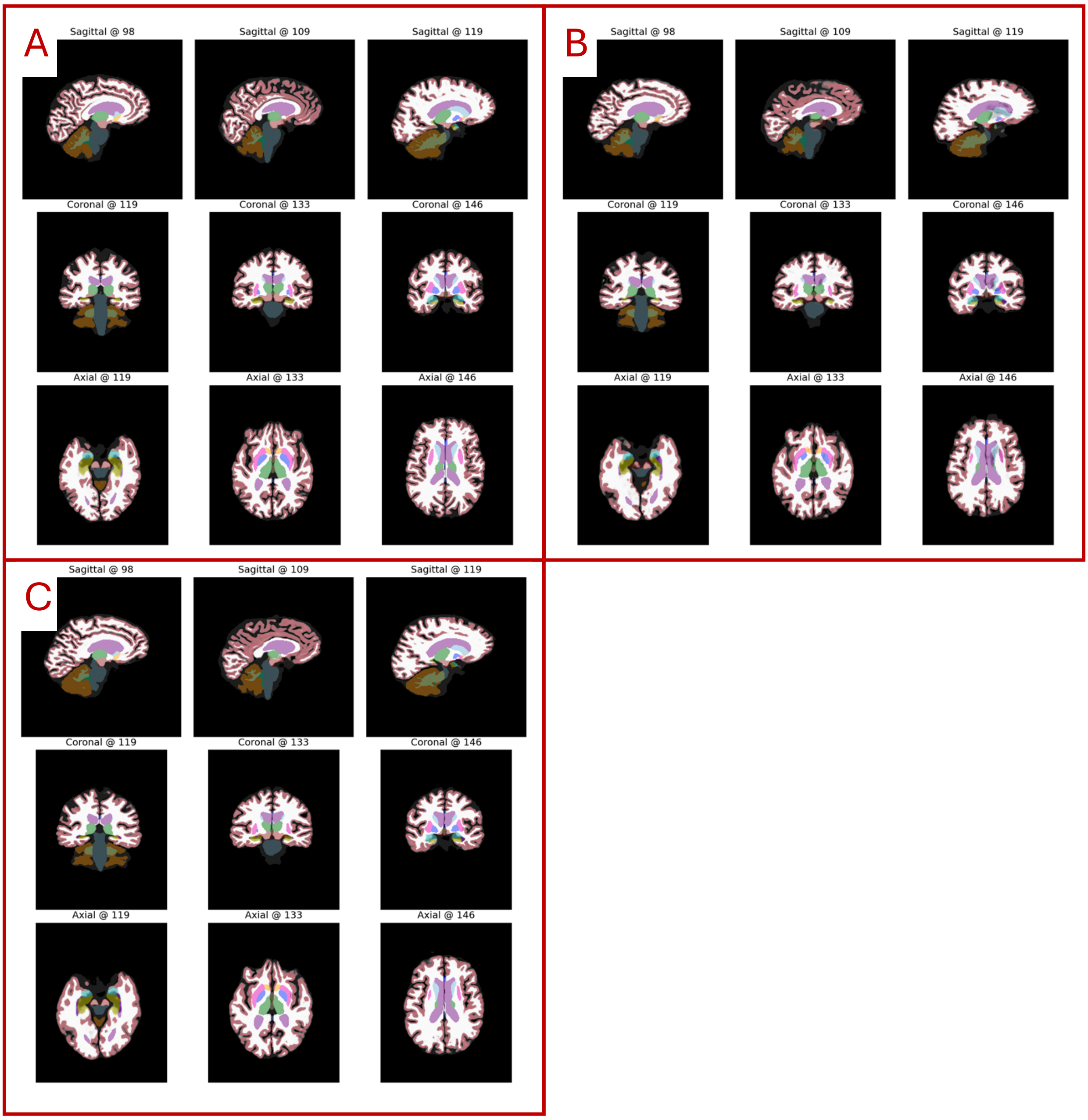

Figure S5: Quality control screenshots for `recon-all-clinical` outputs. Panel A shows HF T1; Panel B shows ULF T1; Panel C shows ULF T1_SSR. All from the same participant.

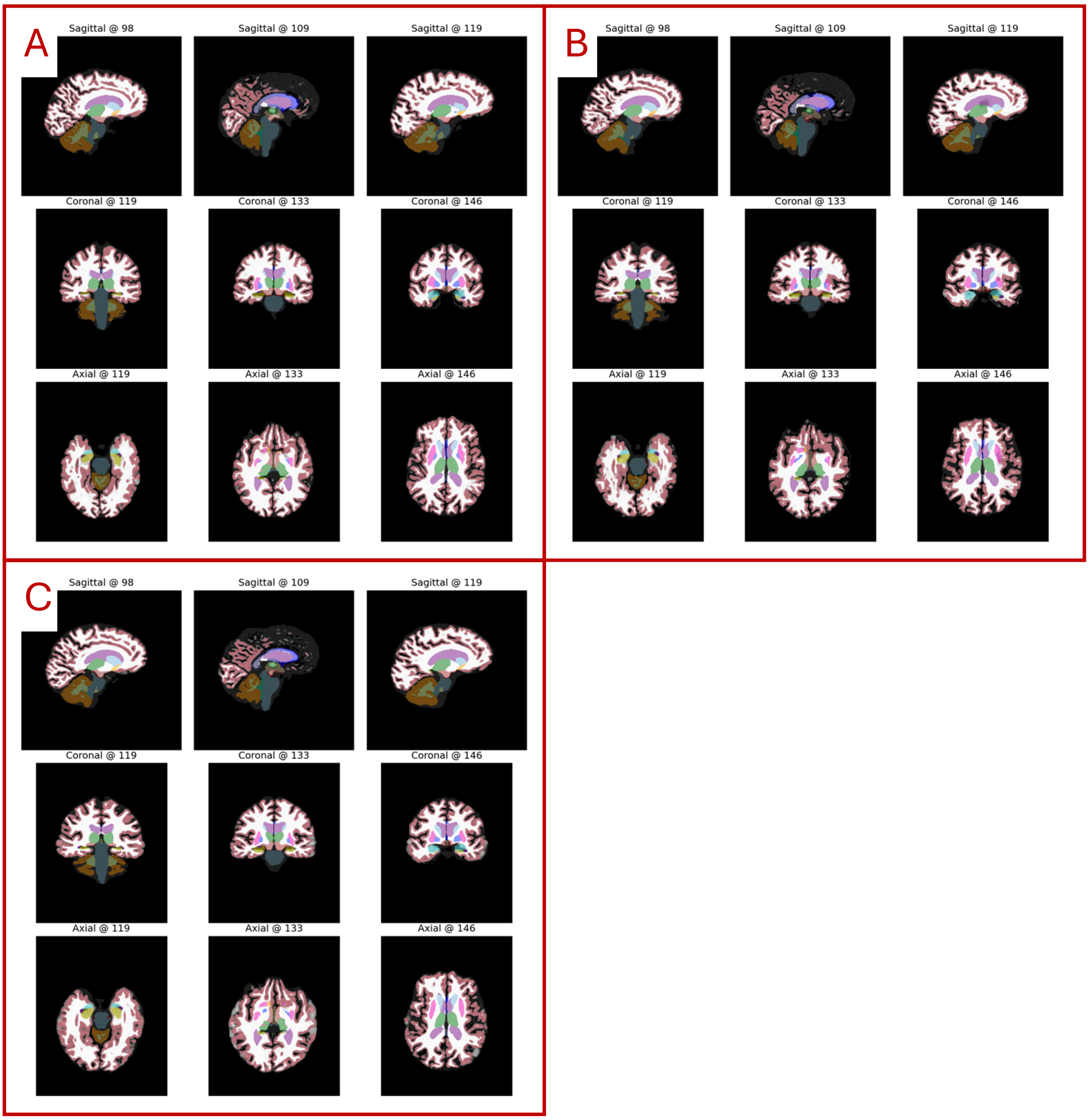

Figure S6: Quality control screenshots for `recon-all-clinical` outputs. Panel A shows HF T2; Panel B shows ULF T2; Panel C shows ULF T2_SSR. All from the same participant.

#### 2.4 QC examples for deep-learning models: SynthBA, DBN, and MIDI

The deep-learning brain-age models used minimal preprocessing: skull-stripping with SynthStrip and linear registration to the MNI152 template using ANTs. For visual QC, the same lightweight toolbox used for `recon-all-clinical` outputs (see Supplementary Section 2.3) was adapted to generate slice screenshots for ease of inspection. All volumes (HF, ULF, and ULF_SSR) passed this visual QC. Some representative examples are shown below.

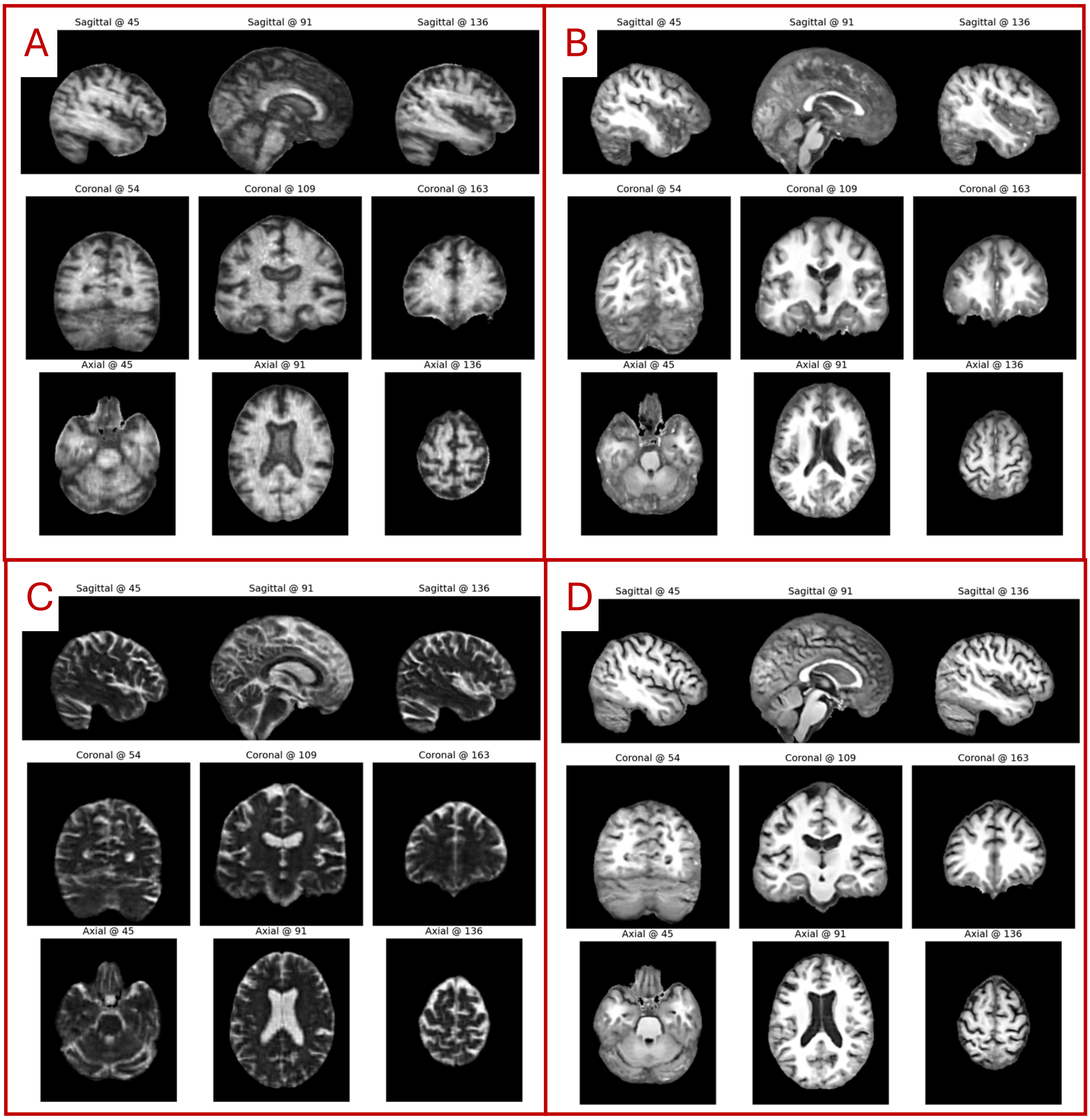

Figure S7: ULF scans after minimal preprocessing (same participant). (A) ULF1 T1; (B) ULF1 T1_SSR; (C) ULF1 T2; (D) ULF1 T2_SSR. This figure illustrates how SynthSR improves the appearance of the ULF scans. In addition, T2 scans are transformed into T1-weighted-like images with SynthSR.

### 3 PyBrainAge

#### 3.1 Predicting missing FreeSurfer labels across pipelines

**Rationale** The label sets from `recon-all` and `recon-all-clinical` are not identical (e.g., vessels and choroid plexus are absent in the latter; CSF differs in extent). Because PyBrainAge expects `recon-all` (v6.0.0-like) features, a lightweight model was trained on `recon-all` outputs to predict five targets needed by PyBrainAge in datasets processed with `recon-all-clinical`.

**Inputs (predictors) and targets** Predictors were volumetric measures from `recon-all` (v7.4.0) on the high-field scans (n=23), comprising subcortical and global volumes.[3] The five *targets* were `Left-vessel`, `Right-vessel`, `Left-choroid-plexus`, `Right-choroid-plexus`, and `CSF`.

**Model and training** A random forest regressor was used (`RandomForestRegressor`, scikit-learn; `random_state=42`) with default hyperparameters. Data were split subject-wise into 80% training and 20% testing. Predictions were constrained to be non-negative by post hoc clipping to zero.

**Evaluation metrics** On the held-out test set the mean squared error (MSE), coefficient of determination ($R^2$), and Pearson's correlation ($r$) for each target are reported.

Table S3: Performance of the label-prediction model on held-out `recon-all` (v7.4.0) high-field scans.

| Target | MSE | $R^2$ | $r$ |
|---|---|---|---|
| Left-vessel | 237.53 | 0.587 | 0.767 |
| Right-vessel | 343.56 | 0.425 | 0.702 |
| Left choroid plexus | 132,391.01 | 0.326 | 0.710 |
| Right choroid plexus | 62,556.81 | 0.385 | 0.716 |
| CSF | 199,065.46 | 0.782 | 0.960 |

**Use in downstream analyses** The predicted values were used solely to complete the PyBrainAge feature set when processing with `recon-all-clinical`. Given the small absolute size and noisier delineation of vessels and choroid plexus, we judged this model-based completion preferable to simple imputation, acknowledging the modest accuracy in performance for those structures.

**Limitations and future directions** We acknowledge that performance for the small ROIs (vessels, choroid plexus) is limited, likely due to the small absolute size of these structures, higher anatomical variability, and the small training sample. In future work we plan to: (1) retrain PyBrainAge directly on `recon-all-clinical` segmentations, or, (2) develop a stronger label-prediction model (to replace the model above) using larger samples, appropriate version (native v6.0.0) and incorporating `recon-all-clinical` features to improve transfer; and (3) other calibration strategies between pipelines to reduce domain shift.

#### 3.2 Predicting sparse missing FreeSurfer segmentations

For sporadic missing values in FreeSurfer outputs (across any pipeline), we applied $k$-nearest neighbours imputation (`KNNImputer`, $k = 5$) to the full feature matrix required for PyBrainAge. Table S4 lists the count of imputed values per batch across all features and participants.

[3]The exact list is: `Left-Putamen`, `Left-Pallidum`, `3rd-Ventricle`, `4th-Ventricle`, `Brain-Stem`, `Left-Hippocampus`, `Left-Amygdala`, `Left-Accumbens-area`, `Left-VentralDC`, `Right-Lateral-Ventricle`, `Right-Inf-Lat-Vent`, `Right-Cerebellum-White-Matter`, `Right-Cerebellum-Cortex`, `Right-Thalamus`, `Right-Caudate`, `Right-Putamen`, `Right-Pallidum`, `Right-Hippocampus`, `Right-Amygdala`, `Right-Accumbens-area`, `Right-VentralDC`, `SubCortGrayVol`, `TotalGrayVol`, `SupraTentorialVol`, `SupraTentorialVolNotVent`, `EstimatedTotalIntraCranialVol`.

Table S4: Number of imputed values per run; Note: 'n/a' marks batches that were not performed.

| **Batch** | **v7.4.0** | **v8.0.0** |
|---|---|---|
| FS T1 | 16 | 1 |
| FS T1+T2 | 16 | 1 |
| FS T1_SSR | 5 | 'n/a' |
| FS T2_SSR | 15 | 'n/a' |
| FS `recon-all-clinical` | 0 | 1 |
| FS `recon-all-clinical` SSR | 'n/a' | 3 |

#### 3.3 Unexpected ULF performance relative to HF

In some cases, ULF scans outperformed HF scans for identical preprocessing combinations within PyBrainAge. For instance, using FreeSurfer v8.0.0, HF data achieved $R^2 = 0.32$, whereas ULF1 reached $R^2 = 0.54$, and ULF1 with SynthSR reached $R^2 = 0.59$.

While unexpected, one plausible explanation for this is that PyBrainAge overfit the training data, which were processed with FreeSurfer v6.0 and may align more closely with outputs from FreeSurfer v7.4.0 than with v8.0.0 (although we did not test this). This is further corroborated by performance drops in high-field when changing FreeSurfer versions (e.g., from $R^2 = 0.65$ in v7.4.0 to $R^2 = 0.36$ in v8.0.0) or when switching from `recon-all` to `recon-all-clinical` ($R^2 = 0.65$ vs. $R^2 = 0.32$, respectively). In ULF, these drops may be less pronounced because of the gains provided by the `recon-all-clinical` pipeline, and/or SynthSR. However, there could be other reasons which we cannot rule out in this study.

### 4 Reliability: complete Correspondence results

In this section, the complete correspondence results are shown. In the main paper only the HF vs ULF1 results were presented, as ULF1 scans were acquired on the same day as HF, whereas ULF2 scans were acquired up to 36 days later. Both HF vs ULF1 and HF vs ULF2 are included in Table S5 below. The Correspondence rank in this version of the table has been recalculated to incorporate both comparisons. This recalculation slightly alters the ranking to the one presented in the main text.

Table S5: Extended correspondence and test-retest reliability. *Correspondence* denotes agreement between ULF scans and a HF homologue ( "HF type" column). Starting sample size is $n$ = 23 except for ULF1 T1 derived scans $n$ = 22. Max scans excluded due to failed preprocessing/QC shown in "max failed $n$" (refer to ULF1 and ULF2 only; all HF passed). The correspondence ranking is based on the median of six per-metric ranks within each Test run (i.e., the three performance metrics $r$, ASPD, and ICC, each evaluated for ULF1 and ULF2, are first converted to ranks, and the median of those six ranks is then used for the final ranking). Numerically-low ranks and dark colours in the heatmap indicate good performance whilst numerically-high ranks and light colours indicate poor performance; Abbreviations: HF, high-field; ULF1/ULF2, ultra-low-field at site 1/site 2; SSR, SynthSR; $r$, Pearson correlation; ASPD, absolute symmetric percent difference; ICC, intraclass correlation coefficient; CI, confidence interval;"=" denotes tied ranks; Py_FSv7_ra, PyBrainAge using FreeSurfer (FS) v7.4 (`recon-all`); Py_FSv8_rac, PyBrainAge using FS v8.0 (`recon-all-clinical`);

| Test | Model | Scan type | Scanner | HF type | failed $n$ | $r$ | ASPD(CI) | ICC(CI) | Ranking |
|---|---|---|---|---|---|---|---|---|---|
| 1 | BrainageR | T1 | ULF1 | T1 | 2 | 0.37 | 15.3 (7.5-34.1) | 0.30 (-0.15-0.65) | 25 |
| 1 | BrainageR | T1 | ULF2 | T1 | 4 | 0.04 | 25.3 (7.1-40.0) | 0.03 (-0.44-0.48) | 25 |
| 2 | BrainageR | T1_SSR | ULF1 | T1 | 0 | 0.84 | 16.6 (6.8-23.5) | 0.77 (0.52-0.90) | 7 |
| 2 | BrainageR | T1_SSR | ULF2 | T1 | 0 | 0.86 | 13.2 (6.6-22.5) | 0.76 (0.52-0.89) | 7 |
| 3 | BrainageR | T2_SSR | ULF1 | T1 | 0 | 0.86 | 17.3 (9.1-25.4) | 0.77 (0.35-0.91) | 6 |
| 3 | BrainageR | T2_SSR | ULF2 | T1 | 0 | 0.89 | 15.7 (8.1-22.0) | 0.80 (0.33-0.93) | 6 |
| 4 | SynthBA | T1 | ULF1 | T1 | 0 | 0.84 | 16.7 (7.3-28.2) | 0.69 (0.19-0.88) | 10 |
| 4 | SynthBA | T1 | ULF2 | T1 | 0 | 0.85 | 22.1 (11.3-36.3) | 0.68 (0.12-0.88) | 10 |
| 5 | SynthBA | T2 | ULF1 | T2 | 0 | 0.93 | 7.6 (2.4-16.4) | 0.92 (0.80-0.97) | 1 |
| 5 | SynthBA | T2 | ULF2 | T2 | 0 | 0.93 | 8.0 (2.2-17.3) | 0.92 (0.79-0.97) | 1 |
| 6 | SynthBA | T1_SSR | ULF1 | T1 | 0 | 0.68 | 27.1 (14.7-40.7) | 0.64 (0.31-0.83) | 11= |
| 6 | SynthBA | T1_SSR | ULF2 | T1 | 0 | 0.86 | 13.9 (6.2-32.8) | 0.82 (0.60-0.92) | 11= |
| 7 | SynthBA | T2_SSR | ULF1 | T2 | 0 | 0.81 | 34.6 (20.0-57.5) | 0.57 (-0.08-0.85) | 17 |
| 7 | SynthBA | T2_SSR | ULF2 | T2 | 0 | 0.78 | 47.7 (24.8-56.2) | 0.53 (-0.09-0.83) | 17 |
| 8 | MIDI-T1 | T1 | ULF1 | T1 | 0 | 0.67 | 24.8 (13.0-36.3) | 0.47 (0.02-0.75) | 19= |
| 8 | MIDI-T1 | T1 | ULF2 | T1 | 0 | 0.54 | 26.9 (12.0-38.4) | 0.37 (0.00-0.67) | 19= |
| 9 | MIDI-T2 | T2 | ULF1 | T2 | 0 | 0.93 | 12.9 (5.6-22.3) | 0.81 (0.60-0.91) | 3 |
| 9 | MIDI-T2 | T2 | ULF2 | T2 | 0 | 0.96 | 12.7 (5.3-19.7) | 0.84 (0.66-0.93) | 3 |
| 10 | MIDI-T1 | T1_SSR | ULF1 | T1 | 0 | 0.62 | 16.5 (5.7-28.6) | 0.57 (0.20-0.80) | 11= |
| 10 | MIDI-T1 | T1_SSR | ULF2 | T1 | 0 | 0.83 | 13.3 (7.0-21.5) | 0.79 (0.57-0.91) | 11= |
| 11 | MIDI-T1 | T2_SSR | ULF1 | T2 | 0 | 0.87 | 25.9 (12.3-31.4) | 0.54 (-0.03-0.81) | 18 |
| 11 | MIDI-T1 | T2_SSR | ULF2 | T2 | 0 | 0.90 | 27.4 (9.7-33.3) | 0.52 (-0.06-0.81) | 18 |
| 12 | DeepBrainNet | T1 | ULF1 | T1 | 0 | 0.54 | 65.4 (29.9-81.6) | 0.10 (-0.07-0.38) | 26 |
| 12 | DeepBrainNet | T1 | ULF2 | T1 | 0 | 0.68 | 60.9 (27.7-78.8) | 0.12 (-0.08-0.42) | 26 |
| 13 | DeepBrainNet | T1_SSR | ULF1 | T1 | 0 | -0.11 | 23.0 (16.3-44.5) | -0.09 (-0.52-0.35) | 23 |
| 13 | DeepBrainNet | T1_SSR | ULF2 | T1 | 0 | 0.65 | 25.4 (13.7-34.9) | 0.35 (-0.06-0.65) | 23 |
| 14 | DeepBrainNet | T2_SSR | ULF1 | T1 | 0 | 0.85 | 23.1 (9.2-42.1) | 0.57 (0.06-0.82) | 16 |
| 14 | DeepBrainNet | T2_SSR | ULF2 | T1 | 0 | 0.82 | 26.7 (9.7-47.4) | 0.52 (0.03-0.79) | 16 |
| 15 | Py_FSv7_ra | T1 | ULF1 | T1 | 0 | 0.66 | 26.2 (11.9-48.0) | 0.43 (-0.08-0.75) | 19= |
| 15 | Py_FSv7_ra | T1 | ULF2 | T1 | 0 | 0.66 | 27.1 (13.1-43.9) | 0.41 (-0.09-0.73) | 19= |
| 16 | Py_FSv7_ra | T1+T2 | ULF1 | T1 | 1 | 0.71 | 33.7 (21.8-55.6) | 0.40 (-0.11-0.75) | 22 |
| 16 | Py_FSv7_ra | T1+T2 | ULF2 | T1 | 4 | 0.74 | 34.3 (13.2-52.4) | 0.41 (-0.11-0.76) | 22 |
| 17 | Py_FSv7_ra | T1_SSR | ULF1 | T1 | 0 | 0.74 | 18.0 (6.9-28.3) | 0.70 (0.41-0.86) | 9 |
| 17 | Py_FSv7_ra | T1_SSR | ULF2 | T1 | 0 | 0.83 | 12.6 (7.0-20.9) | 0.79 (0.58-0.91) | 9 |
| 18 | Py_FSv7_ra | T2_SSR | ULF1 | T1 | 6 | 0.71 | 9.8 (4.9-32.5) | 0.67 (0.30-0.86) | 15 |
| 18 | Py_FSv7_ra | T2_SSR | ULF2 | T1 | 7 | 0.55 | 17.4 (7.1-28.5) | 0.49 (0.05-0.78) | 15 |

Continued on next page

| Test | Model | Scan type | Scanner | HF type | failed $n$ | $r$ | ASPD(CI) | ICC(CI) | Ranking |
|---|---|---|---|---|---|---|---|---|---|
| 19 | Py_FSv7_rac | T1 | ULF1 | T1 | 0 | 0.93 | 24.5 (18.7-29.0) | 0.66 (-0.06-0.91) | 13 |
| 19 | Py_FSv7_rac | T1 | ULF2 | T1 | 0 | 0.84 | 26.0 (11.7-32.7) | 0.60 (-0.09-0.87) | 13 |
| 20 | Py_FSv7_rac | T2 | ULF1 | T2 | 0 | 0.87 | 12.3 (9.1-16.1) | 0.81 (0.56-0.92) | 4 |
| 20 | Py_FSv7_rac | T2 | ULF2 | T2 | 0 | 0.90 | 11.6 (5.4-19.2) | 0.84 (0.54-0.94) | 4 |
| 21 | Py_FSv8_ra | T1 | ULF1 | T1 | 1 | 0.89 | 28.9 (24.0-37.1) | 0.42 (-0.04-0.80) | 21 |
| 21 | Py_FSv8_ra | T1 | ULF2 | T1 | 1 | 0.76 | 33.0 (25.6-37.2) | 0.34 (-0.07-0.72) | 21 |
| 22 | Py_FSv8_ra | T1+T2 | ULF1 | T1 | 1 | 0.73 | 43.4 (25.8-51.3) | 0.23 (-0.05-0.62) | 24 |
| 22 | Py_FSv8_ra | T1+T2 | ULF2 | T1 | 3 | 0.72 | 41.5 (28.1-46.4) | 0.24 (-0.06-0.63) | 24 |
| 23 | Py_FSv8_rac | T1 | ULF1 | T1 | 0 | 0.92 | 24.4 (16.2-30.5) | 0.66 (-0.07-0.91) | 14 |
| 23 | Py_FSv8_rac | T1 | ULF2 | T1 | 0 | 0.83 | 26.4 (8.1-31.9) | 0.59 (-0.09-0.86) | 14 |
| 24 | Py_FSv8_rac | T1_SSR | ULF1 | T1 | 0 | 0.87 | 13.0 (7.5-17.7) | 0.81 (0.50-0.92) | 5 |
| 24 | Py_FSv8_rac | T1_SSR | ULF2 | T1 | 0 | 0.89 | 9.1 (6.3-22.9) | 0.81 (0.40-0.93) | 5 |
| 25 | Py_FSv8_rac | T2 | ULF1 | T2 | 0 | 0.88 | 20.5 (6.3-27.0) | 0.72 (-0.04-0.91) | 8 |
| 25 | Py_FSv8_rac | T2 | ULF2 | T2 | 0 | 0.89 | 21.4 (10.9-25.6) | 0.70 (-0.07-0.91) | 8 |
| 26 | Py_FSv8_rac | T2_SSR | ULF1 | T2 | 0 | 0.93 | 12.2 (4.2-14.4) | 0.89 (0.55-0.96) | 2 |
| 26 | Py_FSv8_rac | T2_SSR | ULF2 | T2 | 0 | 0.94 | 9.9 (7.4-17.2) | 0.87 (0.31-0.96) | 2 |

### 5 All brain-age model analyses plots

In this section, all brain-age model analyses plots are shown, organised by model.

#### 5.1 BrainageR

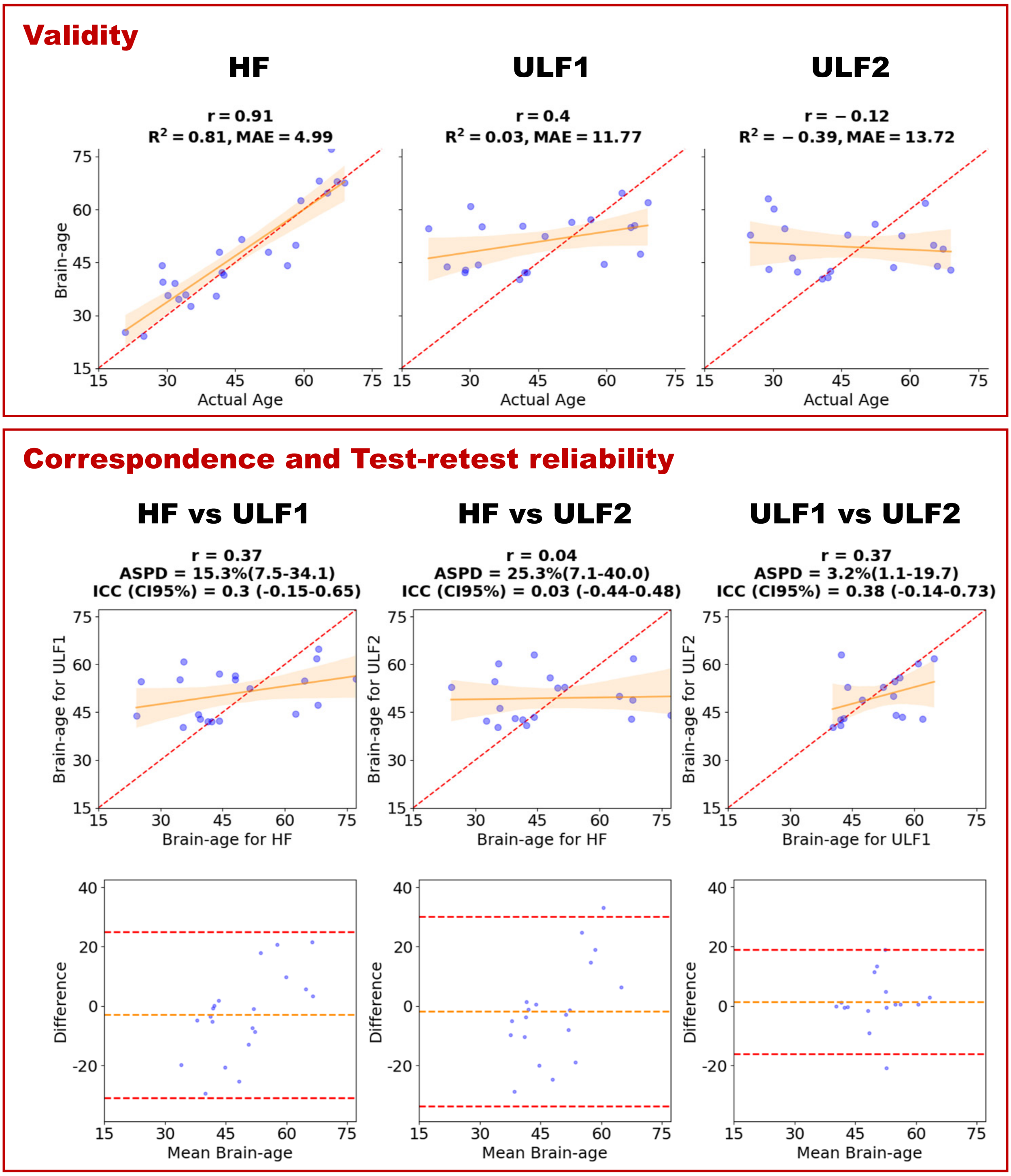

Figure S8: **BrainageR on T1-weighted scans**. Top panel (validity): three scatter plots of brain-age vs. actual age for (1) HF T1, (2) ULF1 T1, and (3) ULF2 T1. Bottom panel (correspondence and test-retest reliability): the upper row shows, in this order from the left, HF-ULF correspondence scatter plots for (1) HF T1 vs. ULF1 T1, (2) HF T1 vs. ULF2 T1, and (3) test-retest reliability (ULF1 T1 vs. ULF2 T1). The lower row shows the corresponding Bland–Altman plots for the same pairings. Scatter plots include the identity line (red dashed) and a least-squares fit with a 95% confidence band. Abbreviations: Pearson correlation $r$; coefficient of determination $R^2$; mean absolute error (MAE); absolute symmetric percent difference (ASPD) with 95% confidence interval (CI); and intraclass correlation coefficient (ICC) with 95% CI; HF = high-field; ULF1/ULF2 = ultra-low-field sites 1/2.

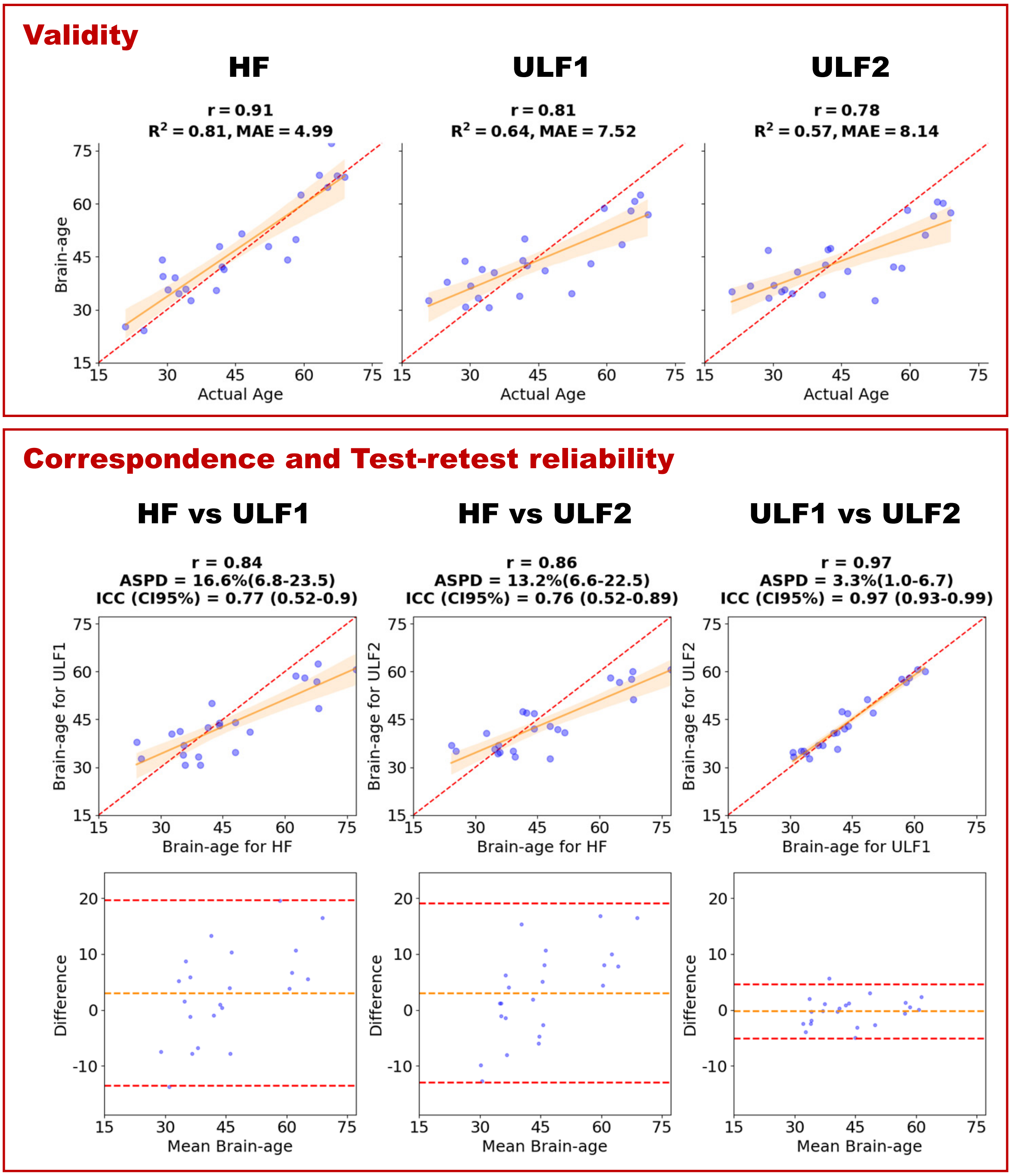

Figure S9: **BrainageR on T1-weighted scans with SynthSR (SSR)**. Top panel (validity): three scatter plots of brain-age vs. actual age for (1) HF T1, (2) ULF1 T1_SSR, and (3) ULF2 T1_SSR. Bottom panel (correspondence and test-retest reliability): the upper row shows, in this order from the left, HF-ULF correspondence scatter plots for (1) HF T1 vs. ULF1 T1_SSR, (2) HF T1 vs. ULF2 T1_SSR, and (3) test-retest reliability (ULF1 T1_SSR vs. ULF2 T1_SSR). The lower row shows the corresponding Bland–Altman plots for the same pairings. Scatter plots include the identity line (red dashed) and a least-squares fit with a 95% confidence band. Abbreviations: Pearson correlation $r$; coefficient of determination $R^2$; mean absolute error (MAE); absolute symmetric percent difference (ASPD) with 95% confidence interval (CI); and intraclass correlation coefficient (ICC) with 95% CI; HF = high-field; ULF1/ULF2 = ultra-low-field sites 1/2.

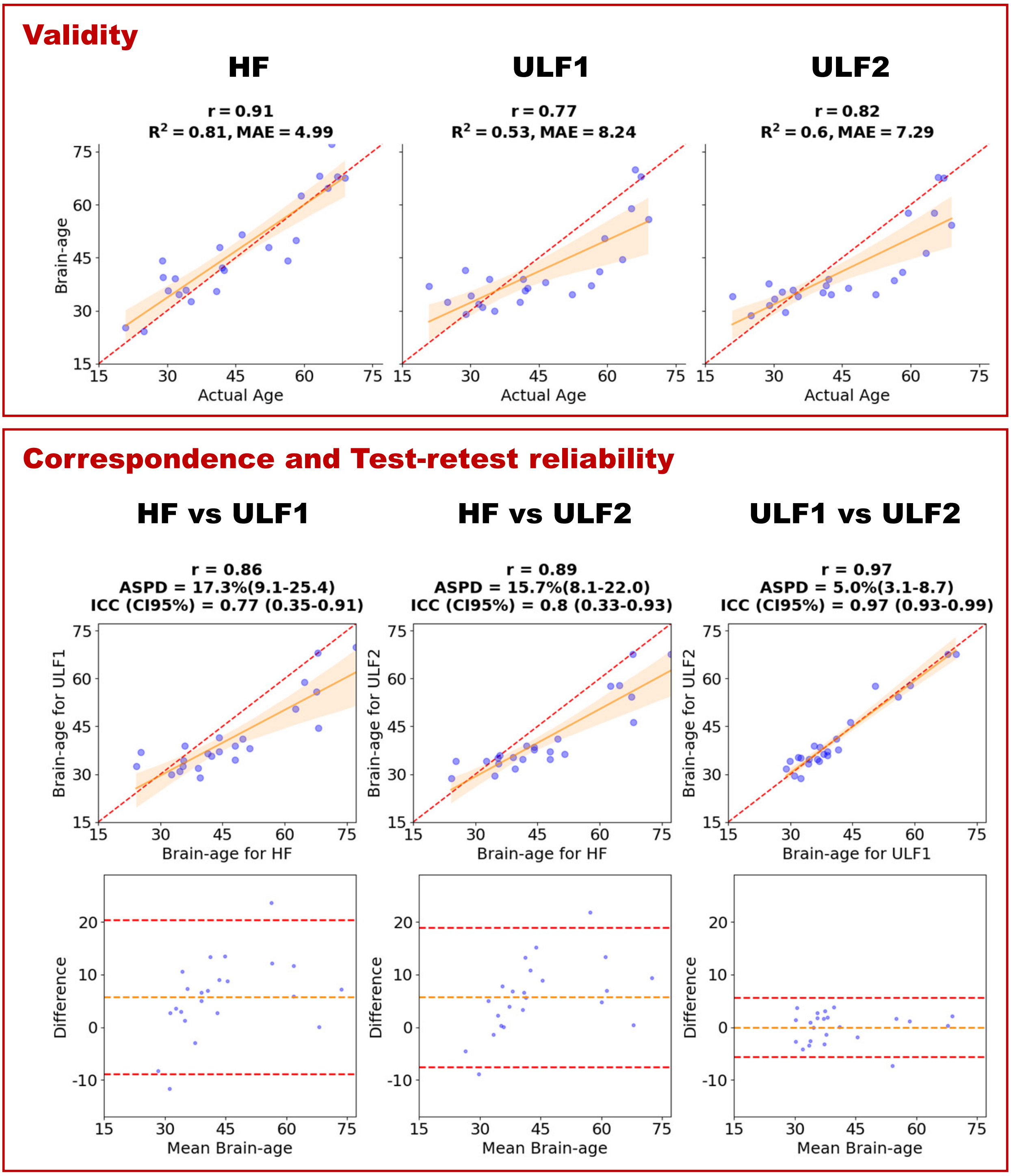

Figure S10: **BrainageR on T2-weighted scans with SynthSR (SSR)**. Top panel (validity): three scatter plots of brain-age vs. actual age for (1) HF T1, (2) ULF1 T2_SSR, and (3) ULF2 T2_SSR. Bottom panel (correspondence and test-retest reliability): the upper row shows, in this order from the left, HF-ULF correspondence scatter plots for (1) HF T1 vs. ULF1 T2_SSR, (2) HF T1 vs. ULF2 T2_SSR, and (3) test-retest reliability (ULF1 T2_SSR vs. ULF2 T2_SSR). The lower row shows the corresponding Bland–Altman plots for the same pairings. Scatter plots include the identity line (red dashed) and a least-squares fit with a 95% confidence band. Abbreviations: Pearson correlation $r$; coefficient of determination $R^2$; mean absolute error (MAE); absolute symmetric percent difference (ASPD) with 95% confidence interval (CI); and intraclass correlation coefficient (ICC) with 95% CI; HF = high-field; ULF1/ULF2 = ultra-low-field sites 1/2.

#### 5.2 SynthBA

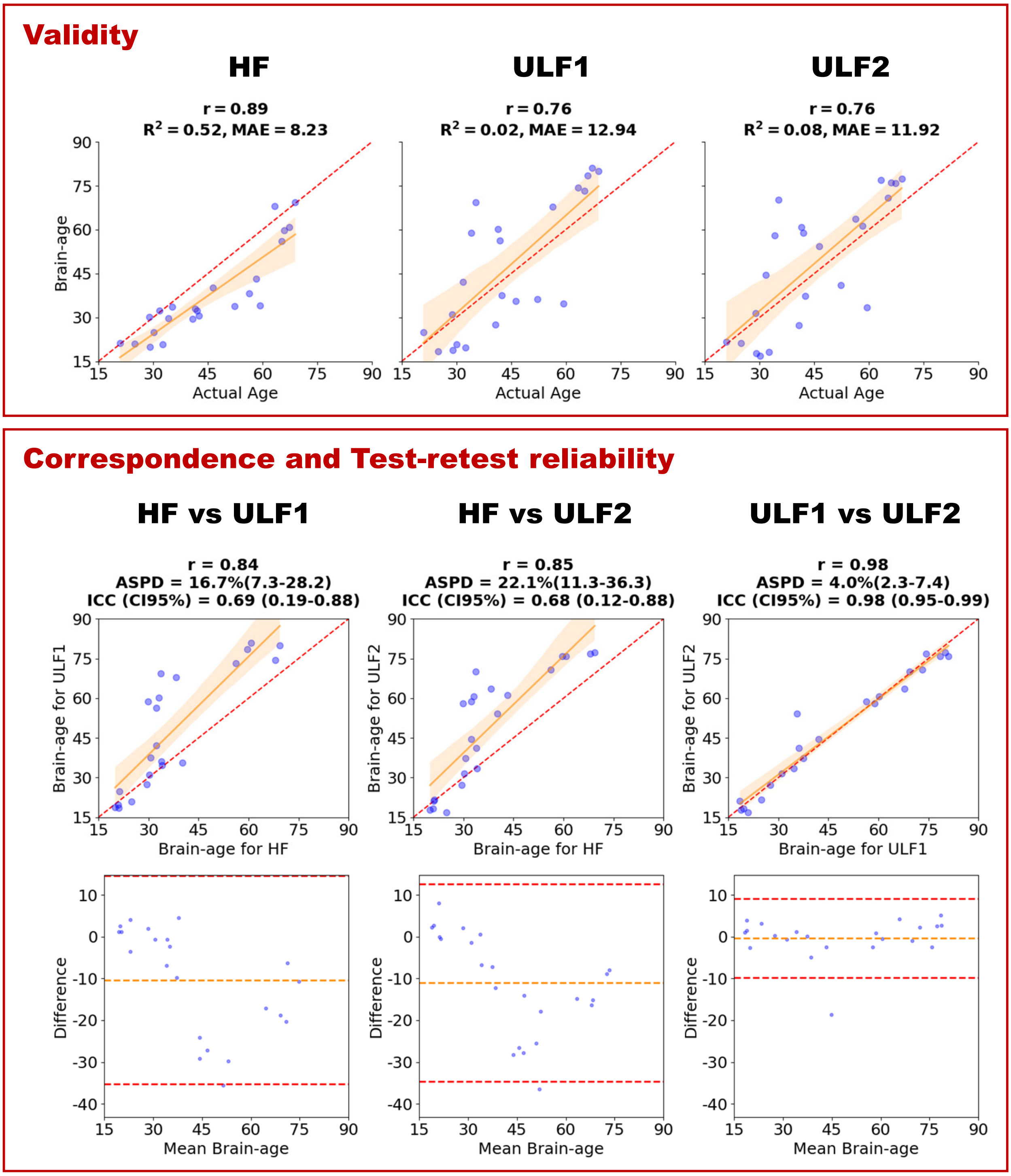

Figure S11: **SynthBA on T1-weighted scans**. Top panel (validity): three scatter plots of brain-age vs. actual age for (1) HF T1, (2) ULF1 T1, and (3) ULF2 T1. Bottom panel (correspondence and test-retest reliability): the upper row shows, in this order from the left, HF-ULF correspondence scatter plots for (1) HF T1 vs. ULF1 T1, (2) HF T1 vs. ULF2 T1, and (3) test-retest reliability (ULF1 T1 vs. ULF2 T1). The lower row shows the corresponding Bland–Altman plots for the same pairings. Scatter plots include the identity line (red dashed) and a least-squares fit with a 95% confidence band. Abbreviations: Pearson correlation $r$; coefficient of determination $R^2$; mean absolute error (MAE); absolute symmetric percent difference (ASPD) with 95% confidence interval (CI); and intraclass correlation coefficient (ICC) with 95% CI; HF = high-field; ULF1/ULF2 = ultra-low-field sites 1/2.

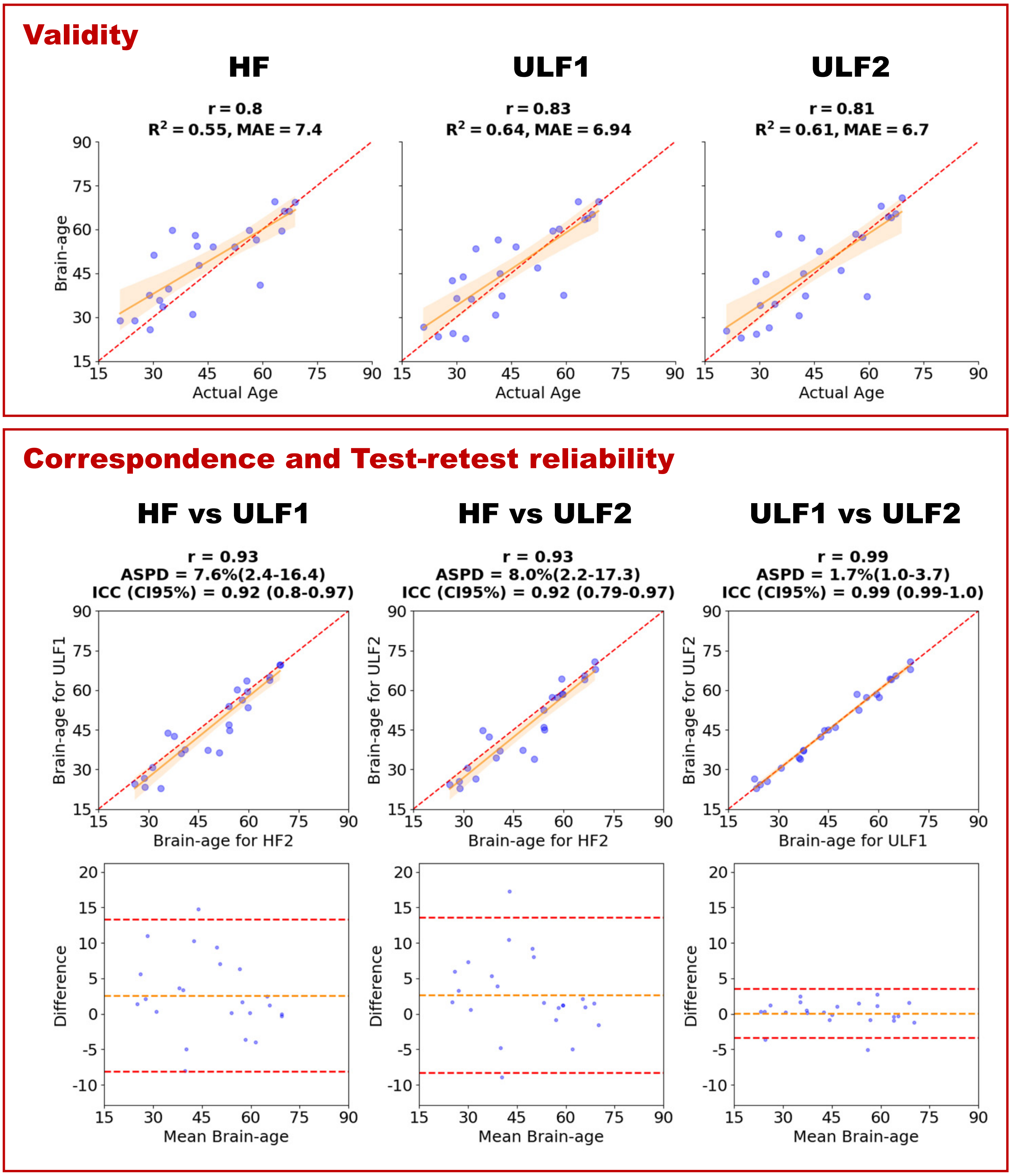

Figure S12: **SynthBA on T2-weighted scans**. Top panel (validity): three scatter plots of brain-age vs. actual age for (1) HF T2, (2) ULF1 T2, and (3) ULF2 T2. Bottom panel (correspondence and test-retest reliability): the upper row shows, in this order from the left, HF-ULF correspondence scatter plots for (1) HF T2 vs. ULF1 T2, (2) HF T2 vs. ULF2 T2, and (3) test-retest reliability (ULF1 T2 vs. ULF2 T2). The lower row shows the corresponding Bland–Altman plots for the same pairings. Scatter plots include the identity line (red dashed) and a least-squares fit with a 95% confidence band. Abbreviations: Pearson correlation $r$; coefficient of determination $R^2$; mean absolute error (MAE); absolute symmetric percent difference (ASPD) with 95% confidence interval (CI); and intraclass correlation coefficient (ICC) with 95% CI; HF = high-field; ULF1/ULF2 = ultra-low-field sites 1/2.

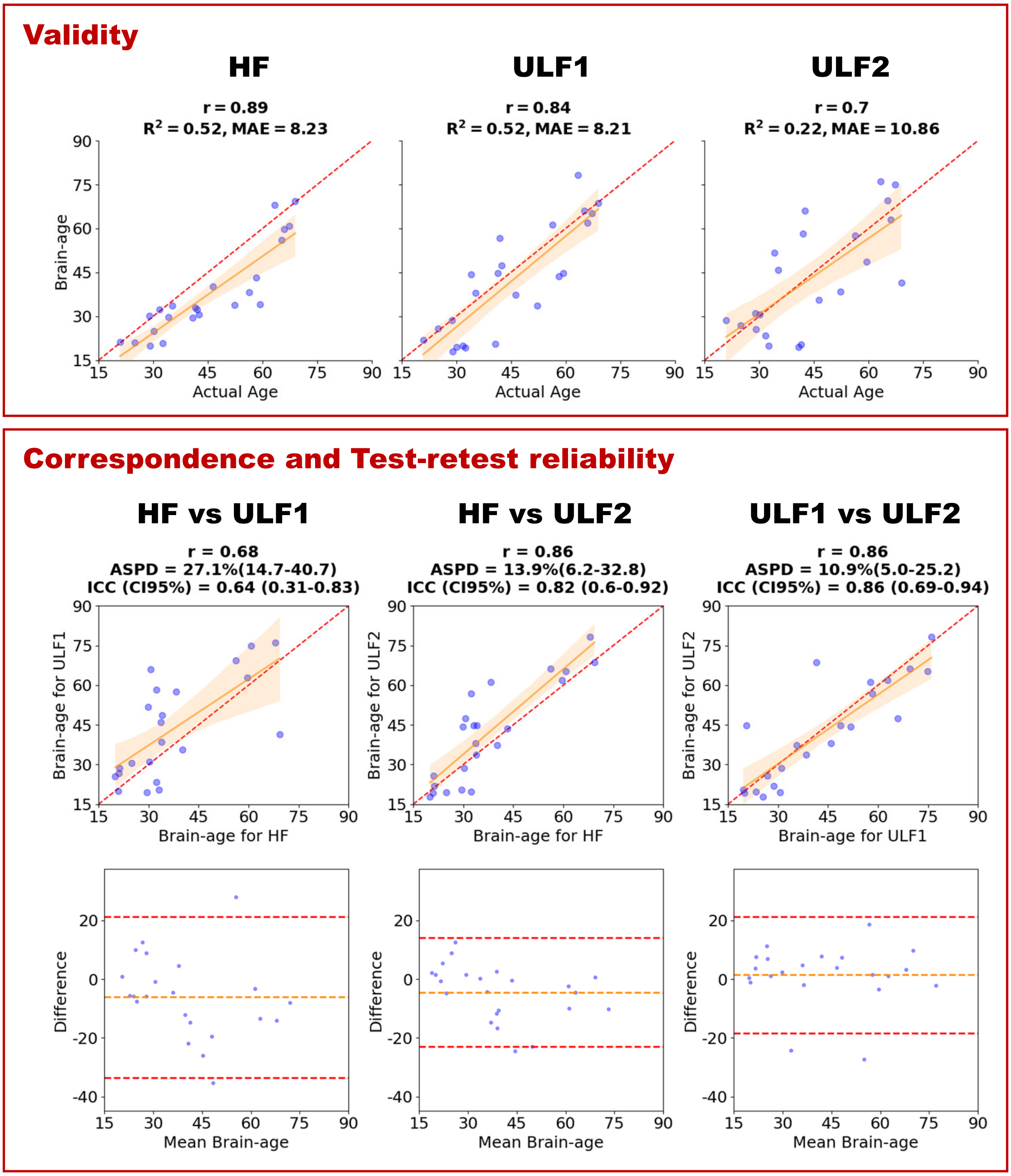

Figure S13: **SynthBA on T1-weighted scans with SynthSR (SSR)**. Top panel (validity): three scatter plots of brain-age vs. actual age for (1) HF T1, (2) ULF1 T1_SSR, and (3) ULF2 T1_SSR. Bottom panel (correspondence and test-retest reliability): the upper row shows, in this order from the left, HF-ULF correspondence scatter plots for (1) HF T1 vs. ULF1 T1_SSR, (2) HF T1 vs. ULF2 T1_SSR, and (3) test-retest reliability (ULF1 T1_SSR vs. ULF2 T1_SSR). The lower row shows the corresponding Bland–Altman plots for the same pairings. Scatter plots include the identity line (red dashed) and a least-squares fit with a 95% confidence band. Abbreviations: Pearson correlation $r$; coefficient of determination $R^2$; mean absolute error (MAE); absolute symmetric percent difference (ASPD) with 95% confidence interval (CI); and intraclass correlation coefficient (ICC) with 95% CI; HF = high-field; ULF1/ULF2 = ultra-low-field sites 1/2.

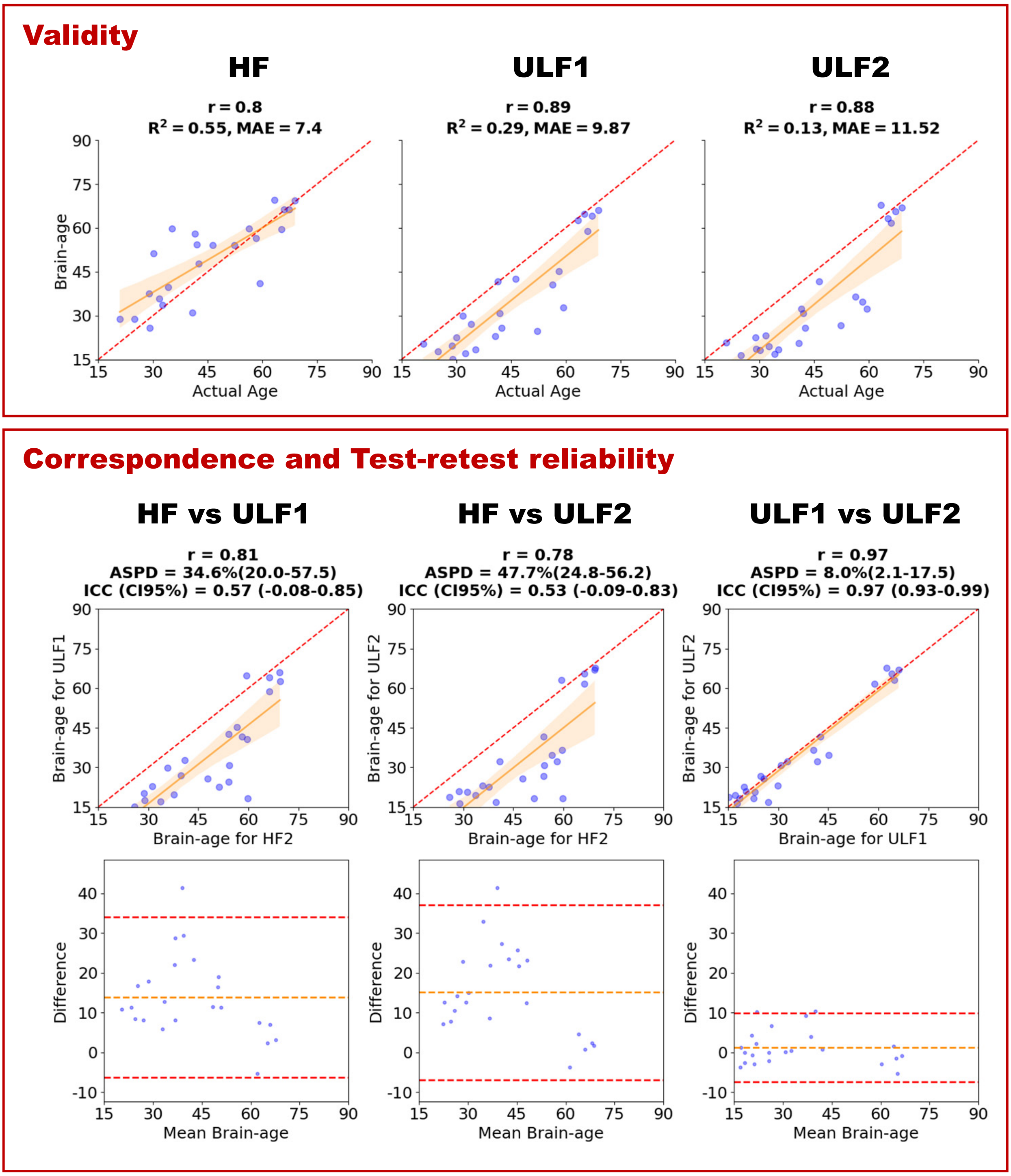

Figure S14: **SynthBA on T2-weighted scans with SynthSR (SSR)**. Top panel (validity): three scatter plots of brain-age vs. actual age for (1) HF T2, (2) ULF1 T2_SSR, and (3) ULF2 T2_SSR. Bottom panel (correspondence and test-retest reliability): the upper row shows, in this order from the left, HF-ULF correspondence scatter plots for (1) HF T2 vs. ULF1 T2_SSR, (2) HF T2 vs. ULF2 T2_SSR, and (3) test-retest reliability (ULF1 T2_SSR vs. ULF2 T2_SSR). The lower row shows the corresponding Bland–Altman plots for the same pairings. Scatter plots include the identity line (red dashed) and a least-squares fit with a 95% confidence band. Abbreviations: Pearson correlation $r$; coefficient of determination $R^2$; mean absolute error (MAE); absolute symmetric percent difference (ASPD) with 95% confidence interval (CI); and intraclass correlation coefficient (ICC) with 95% CI; HF = high-field; ULF1/ULF2 = ultra-low-field sites 1/2.

##### 5.2.1 Best-performing model: SynthBA with single acquisition scans

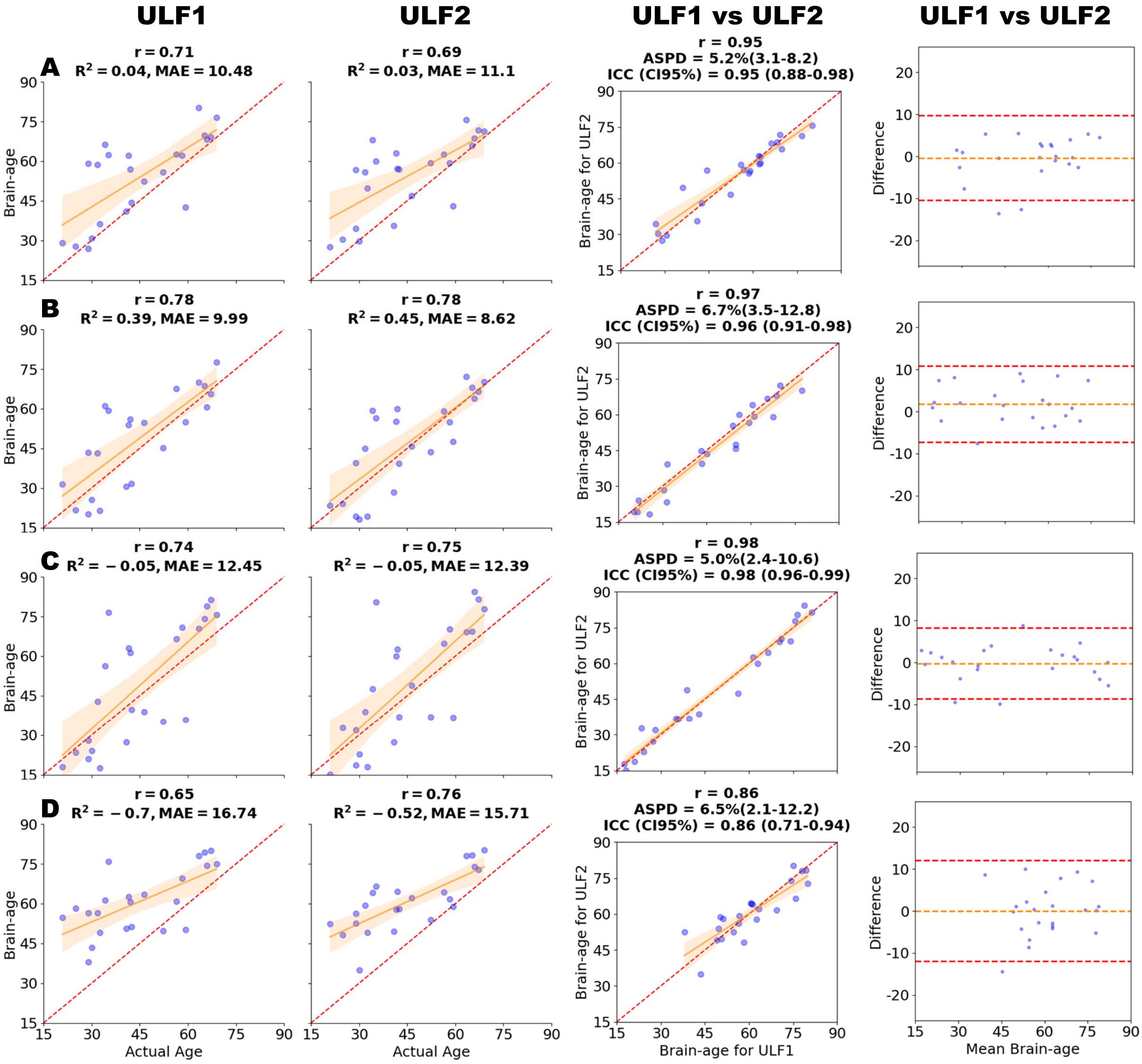

Figure S15: **SynthBA on single-acquisition T1-weighted scans.** Rows (A–D) show acquisition types: (A) axial, (B) sagittal, (C) coronal, and (D) isotropic. Columns are: (1) validity at ULF1 (brain-age vs. actual age), (2) validity at ULF2 (brain-age vs. actual age), (3) test-retest reliability (ULF1 vs. ULF2), and (4) Bland–Altman plots (ULF1 vs. ULF2). Abbreviations: Pearson correlation $r$; coefficient of determination $R^2$; mean absolute error (MAE); absolute symmetric percent difference (ASPD) with 95% confidence interval (CI); and intraclass correlation coefficient (ICC) with 95% CI; HF = high-field; ULF1/ULF2 = ultra-low-field sites 1/2.

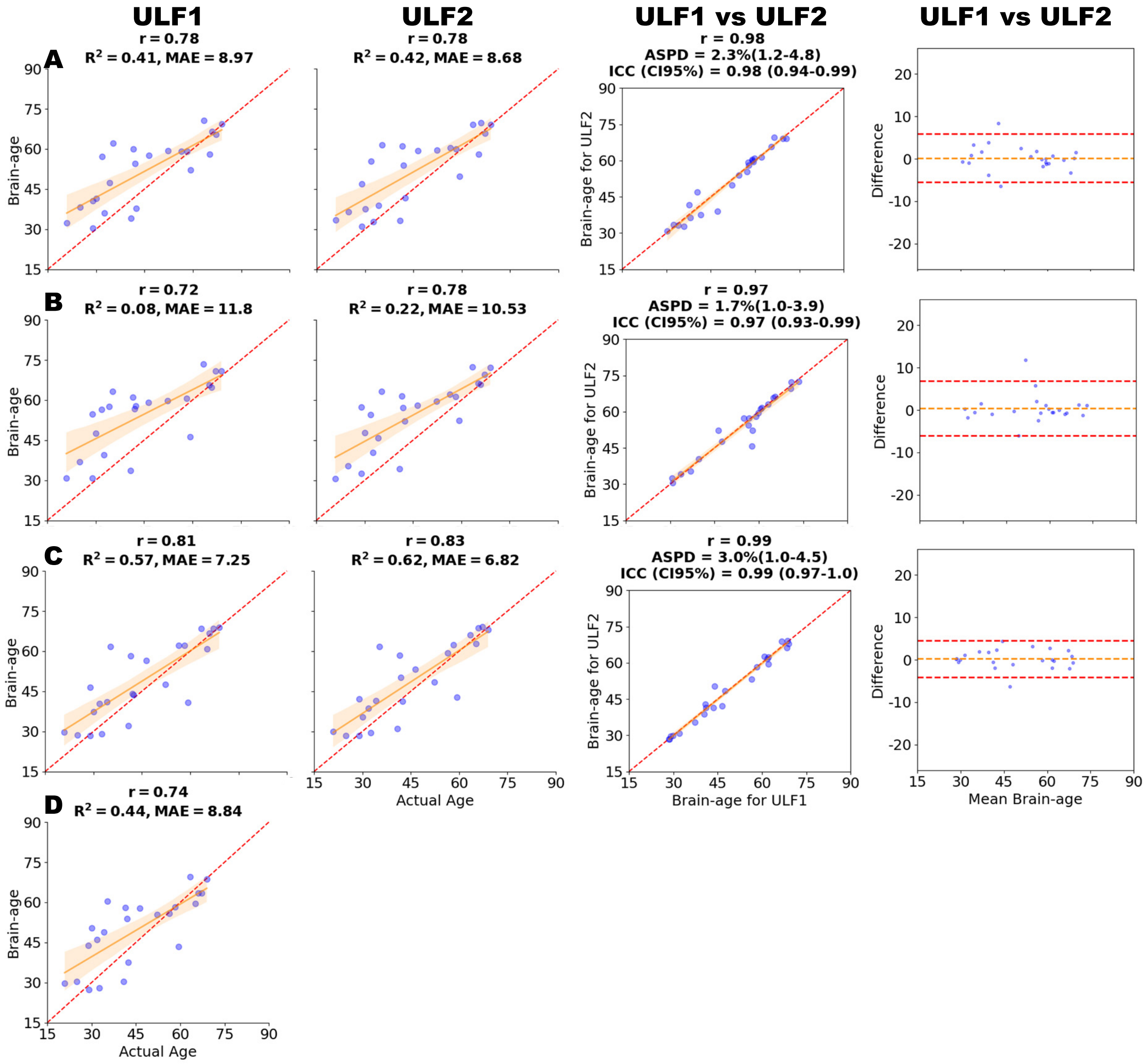

Figure S16: **SynthBA on single-acquisition T2-weighted scans.** Rows (A–D) show acquisition types: (A) axial, (B) sagittal, (C) coronal, and (D) isotropic. Columns are: (1) validity at ULF1 (brain-age vs. actual age), (2) validity at ULF2 (brain-age vs. actual age), (3) test-retest reliability (ULF1 vs. ULF2), and (4) Bland–Altman analysis (ULF1 vs. ULF2). There were no isotropic scans for ULF2 hence blank spaces in row (D). Abbreviations: Pearson correlation $r$; coefficient of determination $R^2$; mean absolute error (MAE); absolute symmetric percent difference (ASPD) with 95% confidence interval (CI); and intraclass correlation coefficient (ICC) with 95% CI; HF = high-field; ULF1/ULF2 = ultra-low-field sites 1/2.

#### 5.3 MIDI

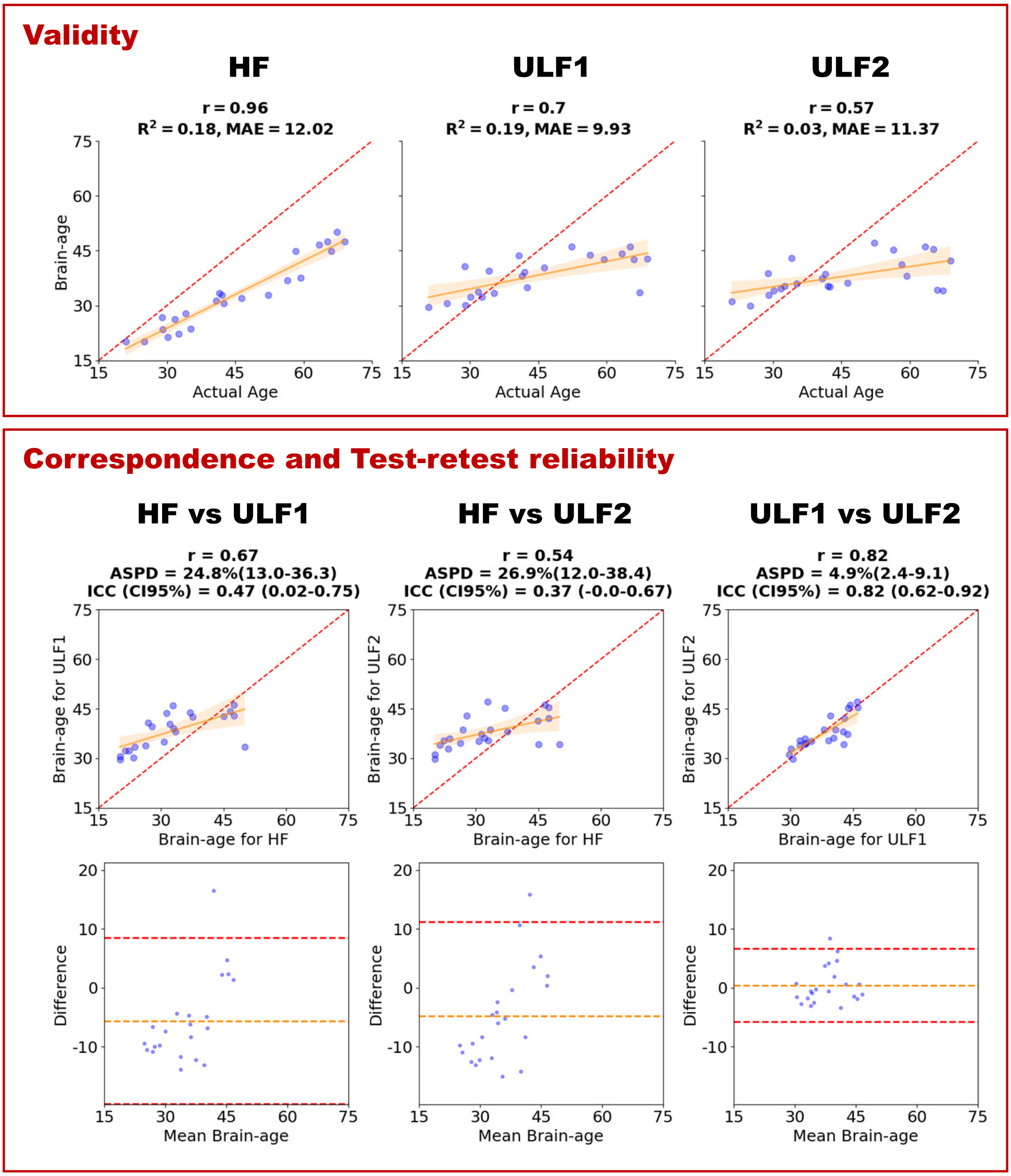

Figure S17: **MIDI (T1 ensemble model) on T1-weighted scans**. Top panel (validity): three scatter plots of brain-age vs. actual age for (1) HF T1, (2) ULF1 T1, and (3) ULF2 T1. Bottom panel (correspondence and test-retest reliability): the upper row shows, in this order from the left, HF-ULF correspondence scatter plots for (1) HF T1 vs. ULF1 T1, (2) HF T1 vs. ULF2 T1, and (3) test-retest reliability (ULF1 T1 vs. ULF2 T1). The lower row shows the corresponding Bland–Altman plots for the same pairings. Scatter plots include the identity line (red dashed) and a least-squares fit with a 95% confidence band. Abbreviations: Pearson correlation $r$; coefficient of determination $R^2$; mean absolute error (MAE); absolute symmetric percent difference (ASPD) with 95% confidence interval (CI); and intraclass correlation coefficient (ICC) with 95% CI; HF = high-field; ULF1/ULF2 = ultra-low-field sites 1/2.

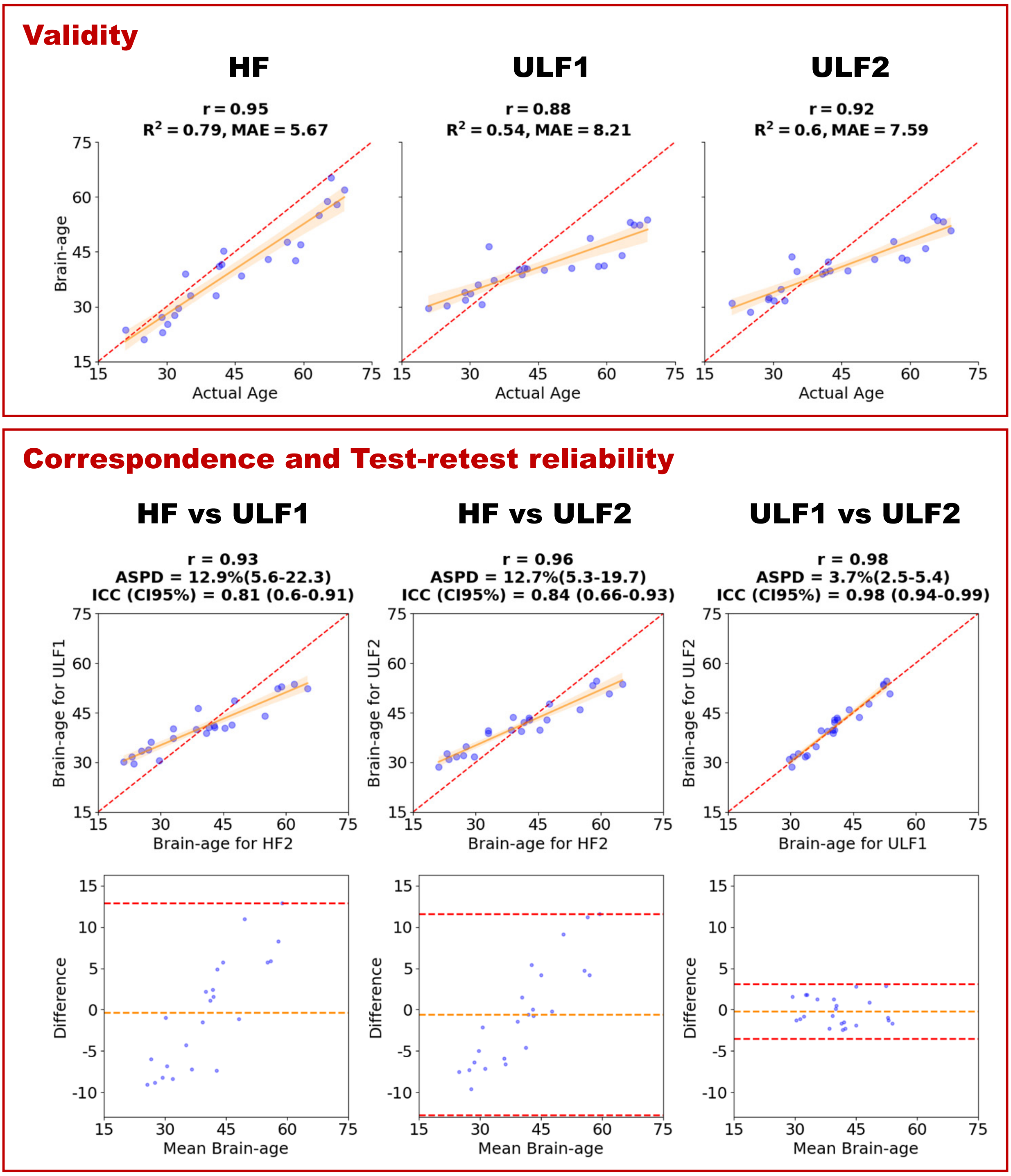

Figure S18: **MIDI (T2 model) on T2-weighted scans**. Top panel (validity): three scatter plots of brain-age vs. actual age for (1) HF T2, (2) ULF1 T2, and (3) ULF2 T2. Bottom panel (correspondence and test-retest reliability): the upper row shows, in this order from the left, HF-ULF correspondence scatter plots for (1) HF T2 vs. ULF1 T2, (2) HF T2 vs. ULF2 T2, and (3) test-retest reliability (ULF1 T2 vs. ULF2 T2). The lower row shows the corresponding Bland–Altman plots for the same pairings. Scatter plots include the identity line (red dashed) and a least-squares fit with a 95% confidence band. Abbreviations: Pearson correlation $r$; coefficient of determination $R^2$; mean absolute error (MAE); absolute symmetric percent difference (ASPD) with 95% confidence interval (CI); and intraclass correlation coefficient (ICC) with 95% CI; HF = high-field; ULF1/ULF2 = ultra-low-field sites 1/2.

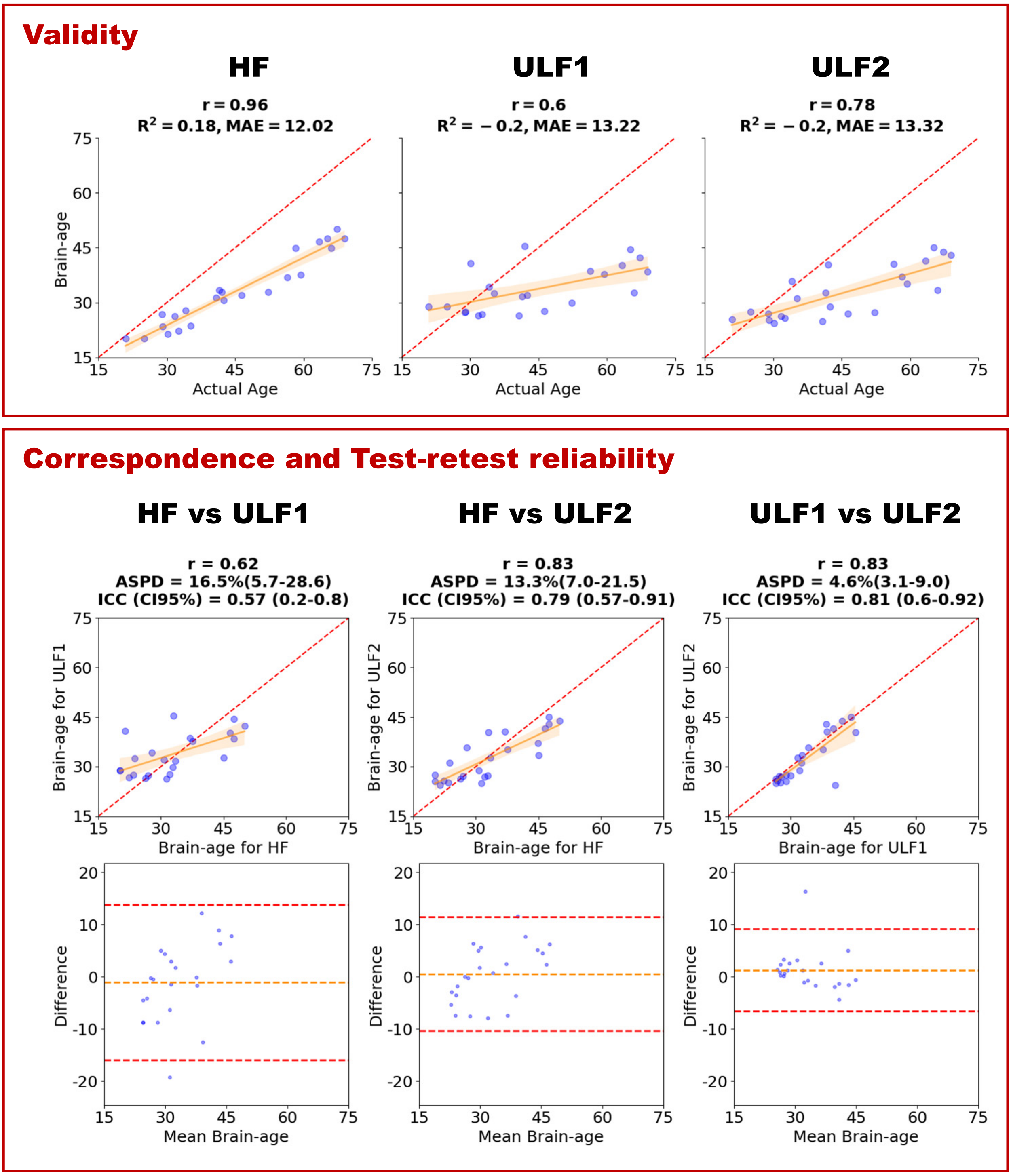

Figure S19: **MIDI (T1 ensemble model) on T1-weighted scans with SynthSR (SSR).** Top panel (validity): three scatter plots of brain-age vs. actual age for (1) HF T1, (2) ULF1 T1_SSR, and (3) ULF2 T1_SSR. Bottom panel (correspondence and test-retest reliability): the upper row shows, in this order from the left, HF-ULF correspondence scatter plots for (1) HF T1 vs. ULF1 T1_SSR, (2) HF T1 vs. ULF2 T1_SSR, and (3) test-retest reliability (ULF1 T1_SSR vs. ULF2 T1_SSR). The lower row shows the corresponding Bland–Altman plots for the same pairings. Scatter plots include the identity line (red dashed) and a least-squares fit with a 95% confidence band. Abbreviations: Pearson correlation $r$; coefficient of determination $R^2$; mean absolute error (MAE); absolute symmetric percent difference (ASPD) with 95% confidence interval (CI); and intraclass correlation coefficient (ICC) with 95% CI; HF = high-field; ULF1/ULF2 = ultra-low-field sites 1/2.

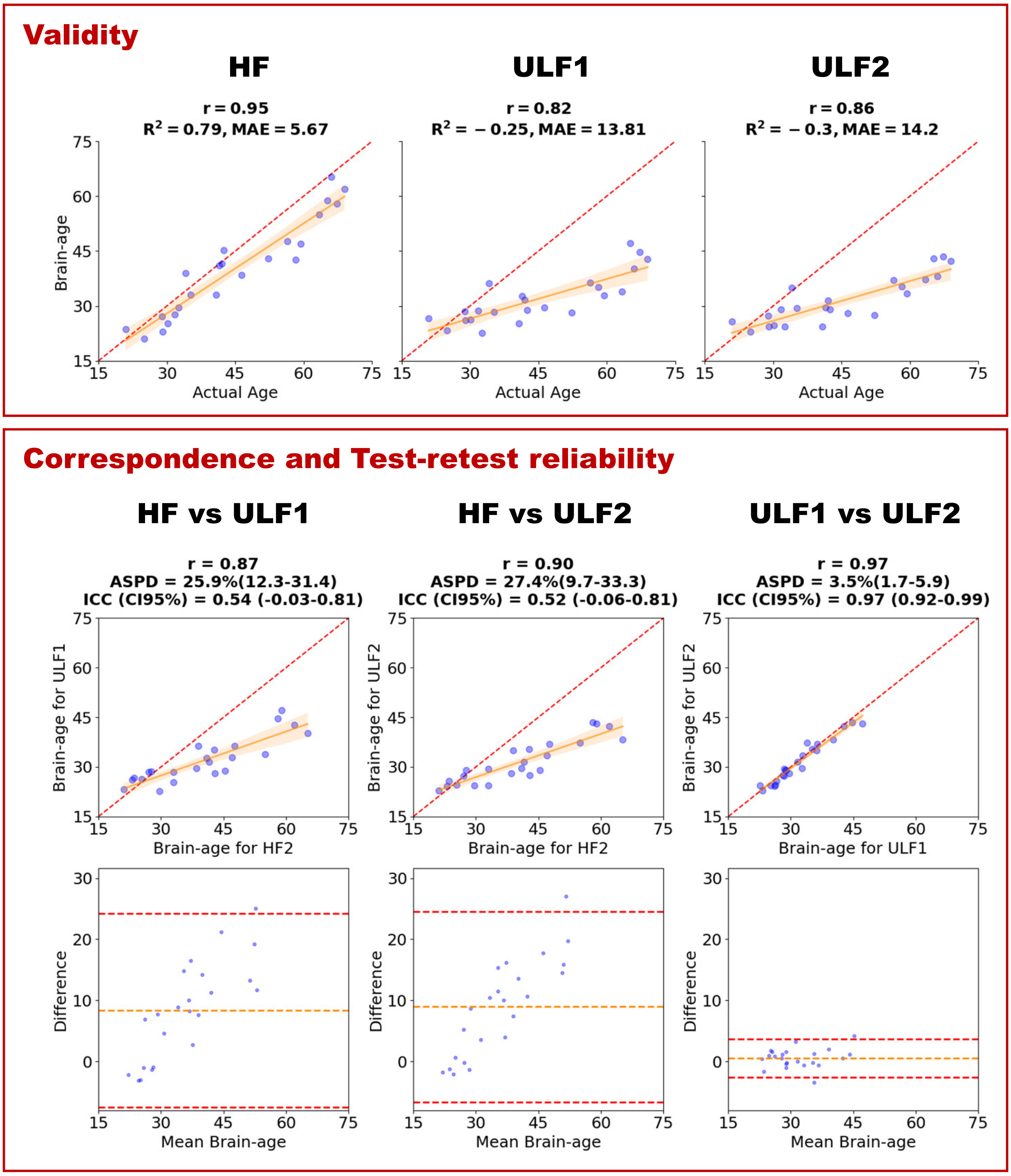

Figure S20: **MIDI (T1 ensemble model) on T2-weighted scans with SynthSR (SSR)**. Top panel (validity): three scatter plots of brain-age vs. actual age for (1) HF T2 (using MIDI-T2 model), (2) ULF1 T2_SSR, and (3) ULF2 T2_SSR. Bottom panel (correspondence and test-retest reliability): the upper row shows, in this order from the left, HF-ULF correspondence scatter plots for (1) HF T2 (using MIDI-T2 model) vs. ULF1 T2_SSR, (2) HF T2 (using MIDI-T2 model) vs. ULF2 T2_SSR, and (3) test-retest reliability (ULF1 T2_SSR vs. ULF2 T2_SSR). The lower row shows the corresponding Bland–Altman plots for the same pairings. Scatter plots include the identity line (red dashed) and a least-squares fit with a 95% confidence band. Abbreviations: Pearson correlation $r$; coefficient of determination $R^2$; mean absolute error (MAE); absolute symmetric percent difference (ASPD) with 95% confidence interval (CI); and intraclass correlation coefficient (ICC) with 95% CI; HF = high-field; ULF1/ULF2 = ultra-low-field sites 1/2.

#### 5.4 DeepBrainNet

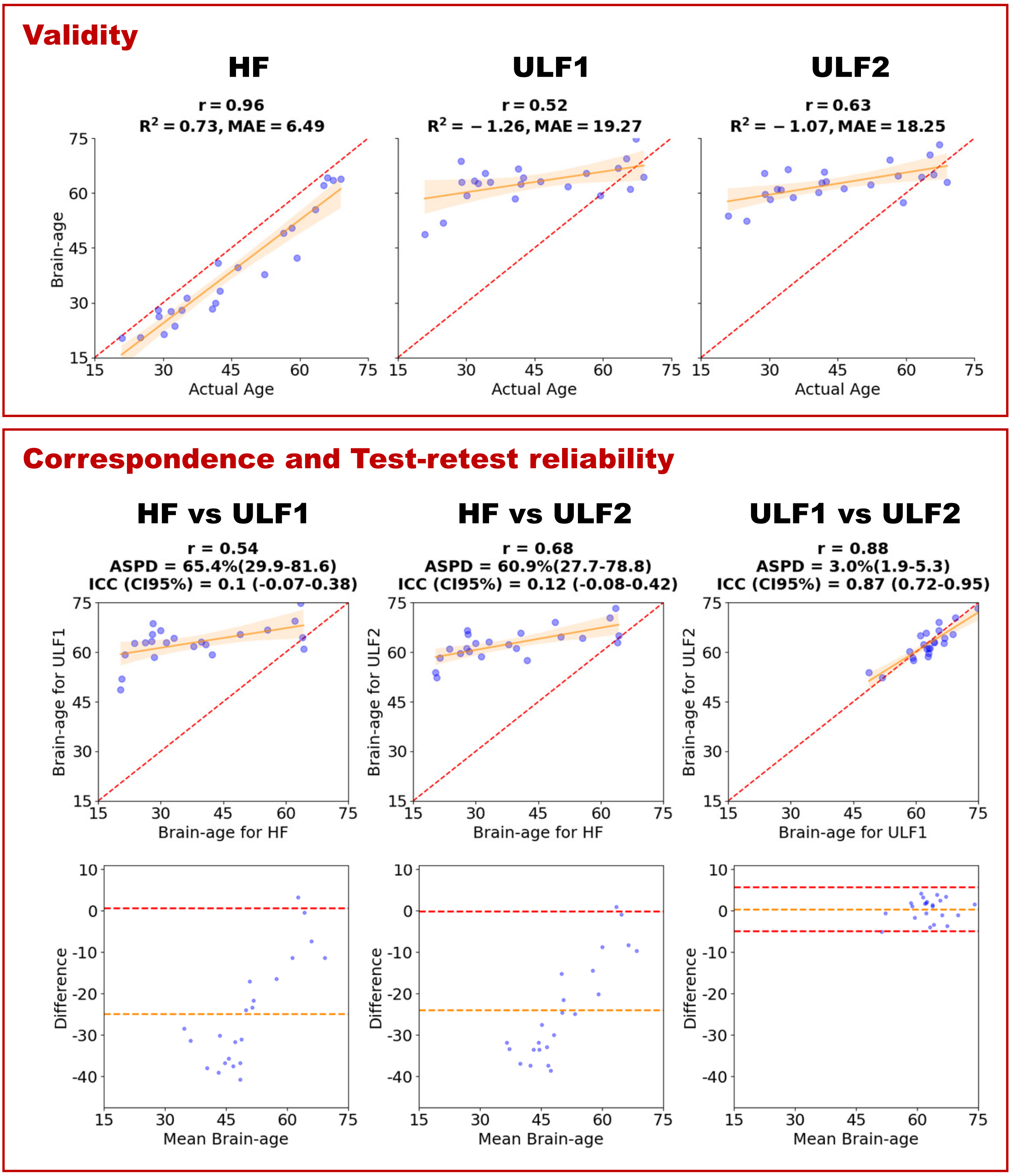

Figure S21: **DeepBrainNet on T1-weighted scans**. Top panel (validity): three scatter plots of brain-age vs. actual age for (1) HF T1, (2) ULF1 T1, and (3) ULF2 T1. Bottom panel (correspondence and test-retest reliability): the upper row shows, in this order from the left, HF-ULF correspondence scatter plots for (1) HF T1 vs. ULF1 T1, (2) HF T1 vs. ULF2 T1, and (3) test-retest reliability (ULF1 T1 vs. ULF2 T1). The lower row shows the corresponding Bland–Altman plots for the same pairings. Scatter plots include the identity line (red dashed) and a least-squares fit with a 95% confidence band. Abbreviations: Pearson correlation $r$; coefficient of determination $R^2$; mean absolute error (MAE); absolute symmetric percent difference (ASPD) with 95% confidence interval (CI); and intraclass correlation coefficient (ICC) with 95% CI; HF = high-field; ULF1/ULF2 = ultra-low-field sites 1/2.

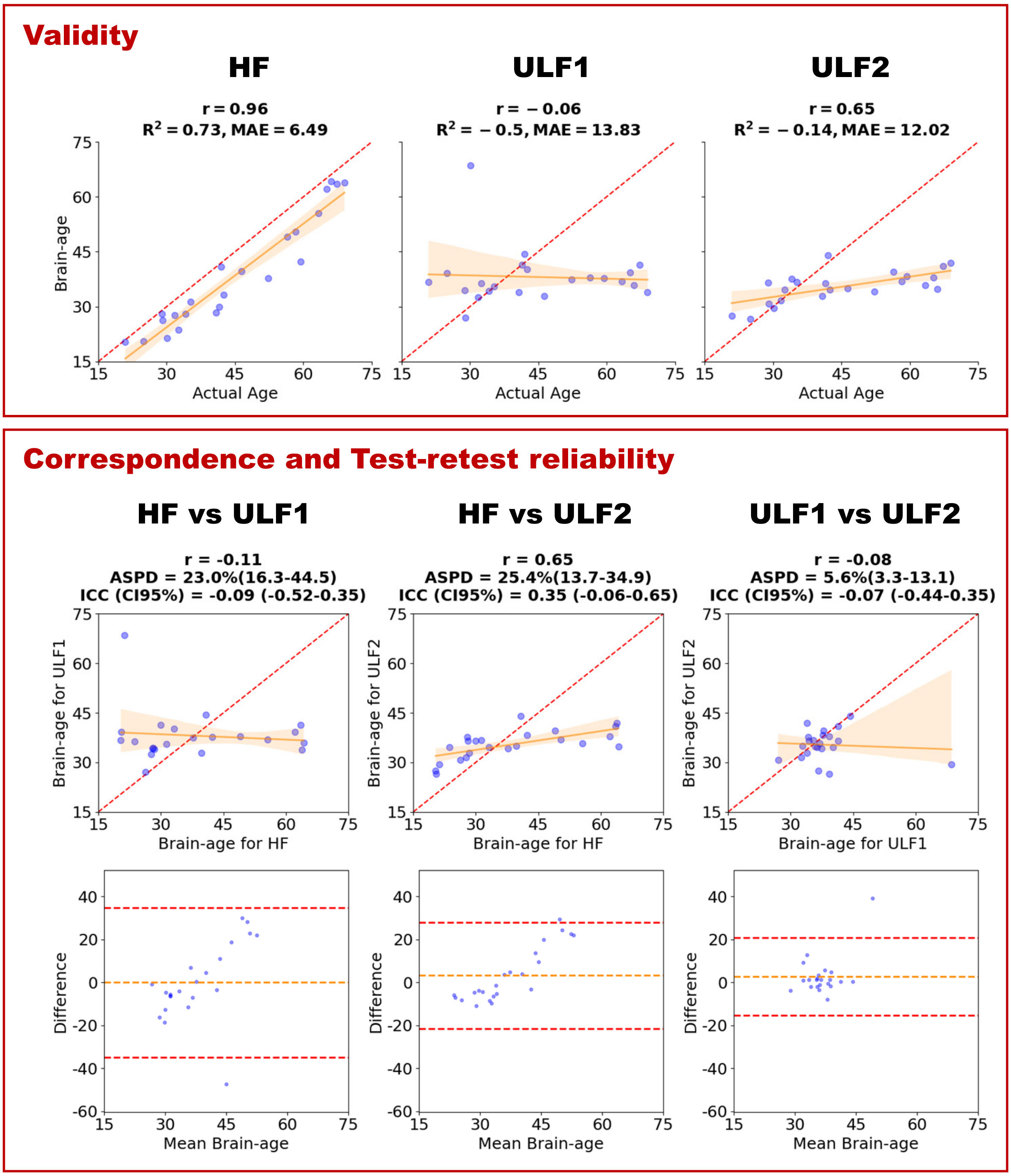

Figure S22: **DeepBrainNet on T1-weighted scans with SynthSR (SSR)**. Top panel (validity): three scatter plots of brain-age vs. actual age for (1) HF T1, (2) ULF1 T1_SSR, and (3) ULF2 T1_SSR. Bottom panel (correspondence and test-retest reliability): the upper row shows, in this order from the left, HF-ULF correspondence scatter plots for (1) HF T1 vs. ULF1 T1_SSR, (2) HF T1 vs. ULF2 T1_SSR, and (3) test-retest reliability (ULF1 T1_SSR vs. ULF2 T1_SSR). The lower row shows the corresponding Bland–Altman plots for the same pairings. Scatter plots include the identity line (red dashed) and a least-squares fit with a 95% confidence band. Abbreviations: Pearson correlation $r$; coefficient of determination $R^2$; mean absolute error (MAE); absolute symmetric percent difference (ASPD) with 95% confidence interval (CI); and intraclass correlation coefficient (ICC) with 95% CI; HF = high-field; ULF1/ULF2 = ultra-low-field sites 1/2.

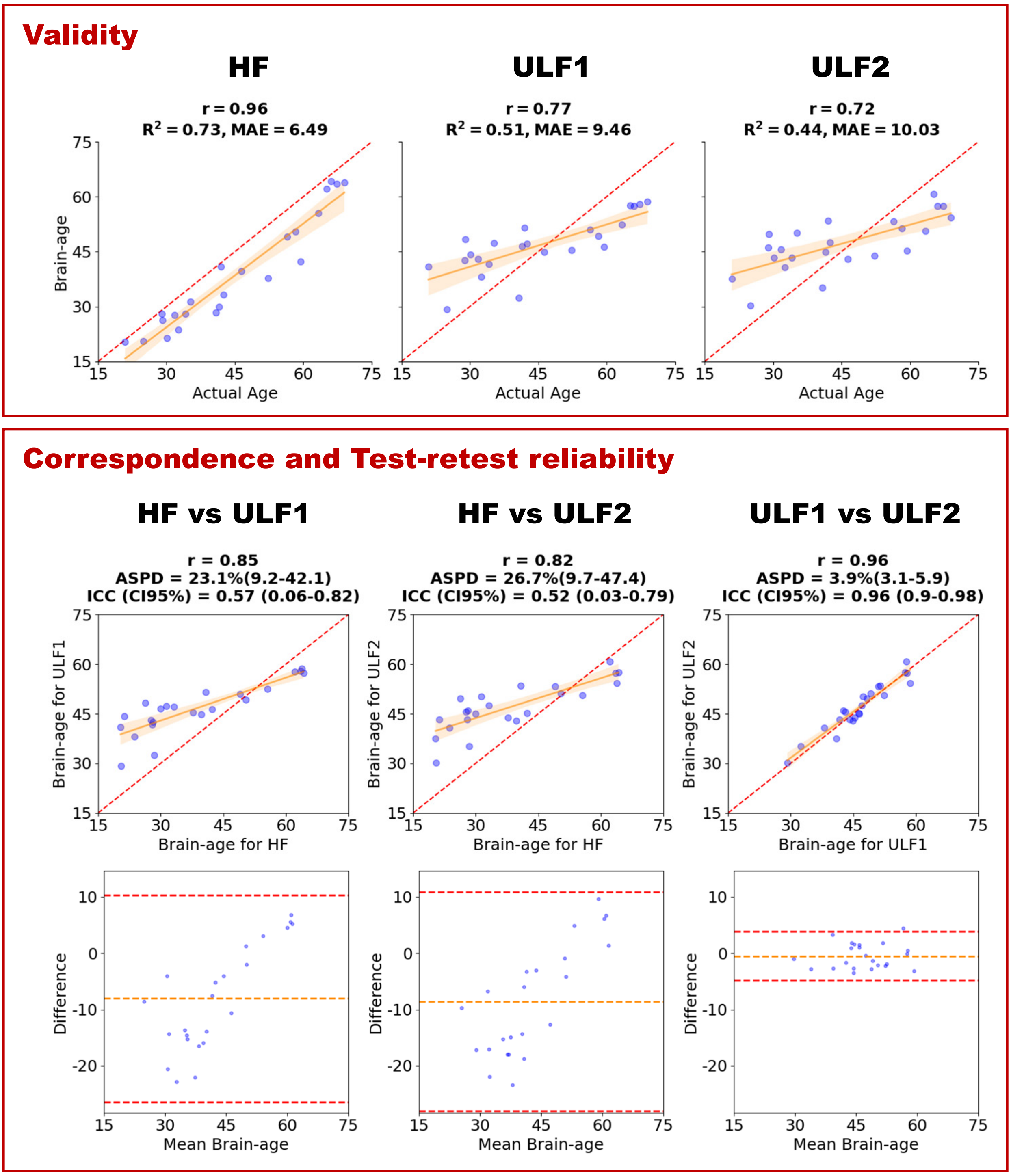

Figure S23: **DeepBrainNet on T2-weighted scans with SynthSR (SSR))**. Top panel (validity): three scatter plots of brain-age vs. actual age for (1) HF T1, (2) ULF1 T2_SSR, and (3) ULF2 T2_SSR. Bottom panel (correspondence and test-retest reliability): the upper row shows, in this order from the left, HF-ULF correspondence scatter plots for (1) HF T1 vs. ULF1 T2_SSR, (2) HF T1 vs. ULF2 T2_SSR, and (3) test-retest reliability (ULF1 T2_SSR vs. ULF2 T2_SSR). The lower row shows the corresponding Bland–Altman plots for the same pairings. Scatter plots include the identity line (red dashed) and a least-squares fit with a 95% confidence band. Abbreviations: Pearson correlation $r$; coefficient of determination $R^2$; mean absolute error (MAE); absolute symmetric percent difference (ASPD) with 95% confidence interval (CI); and intraclass correlation coefficient (ICC) with 95% CI; HF = high-field; ULF1/ULF2 = ultra-low-field sites 1/2.

#### 5.5 PyBrainAge v7.4.0

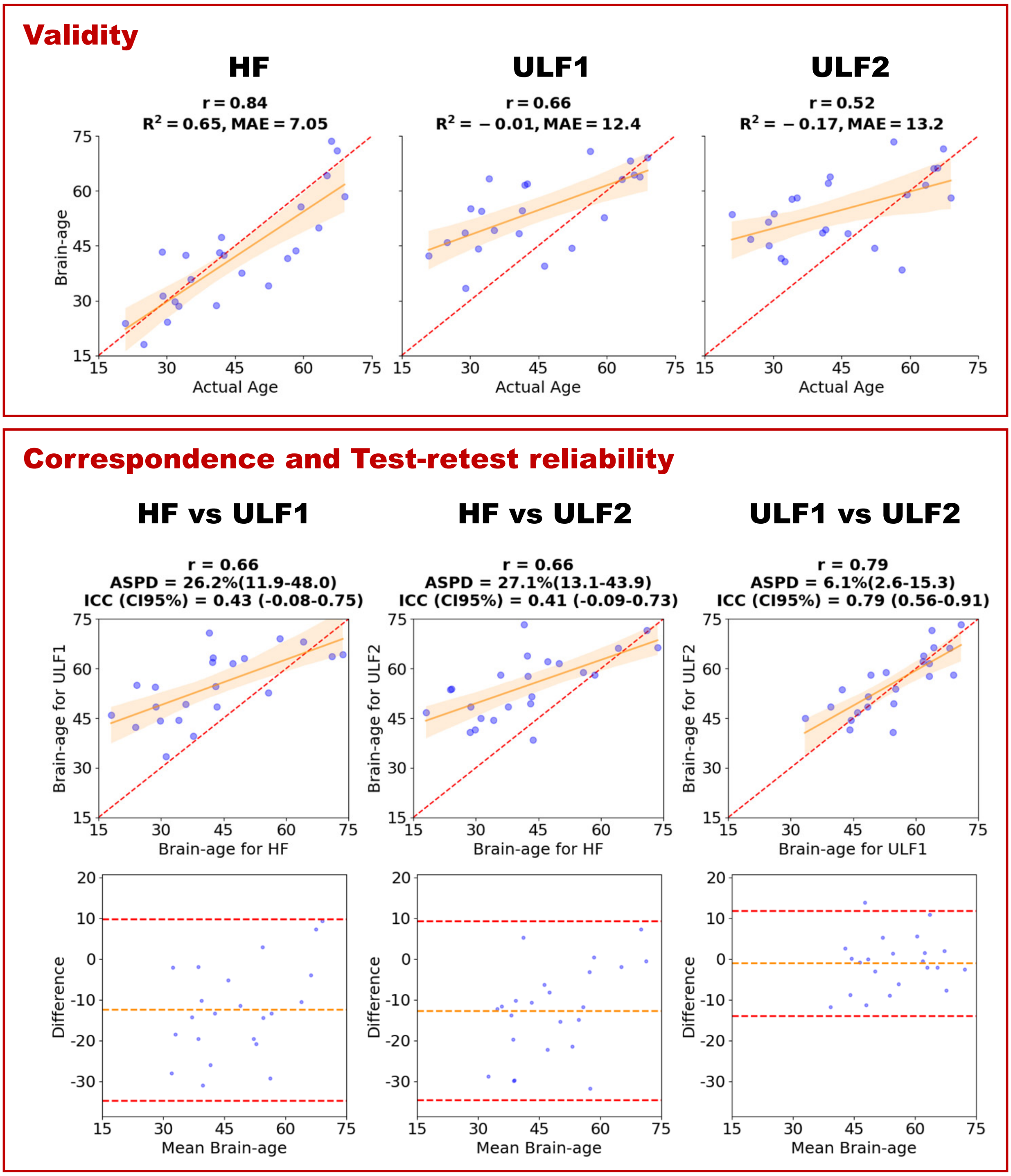

Figure S24: **PyBrainAge on T1-weighted scans using FreeSurfer v7.4 recon-all**. Top panel (validity): three scatter plots of brain-age vs. actual age for (1) HF T1, (2) ULF1 T1, and (3) ULF2 T1. Bottom panel (correspondence and test-retest reliability): the upper row shows, in this order from the left, HF-ULF correspondence scatter plots for (1) HF T1 vs. ULF1 T1, (2) HF T1 vs. ULF2 T1, and (3) test-retest reliability (ULF1 T1 vs. ULF2 T1). The lower row shows the corresponding Bland–Altman plots for the same pairings. Scatter plots include the identity line (red dashed) and a least-squares fit with a 95% confidence band. Abbreviations: Pearson correlation $r$; coefficient of determination $R^2$; mean absolute error (MAE); absolute symmetric percent difference (ASPD) with 95% confidence interval (CI); and intraclass correlation coefficient (ICC) with 95% CI; HF = high-field; ULF1/ULF2 = ultra-low-field sites 1/2.

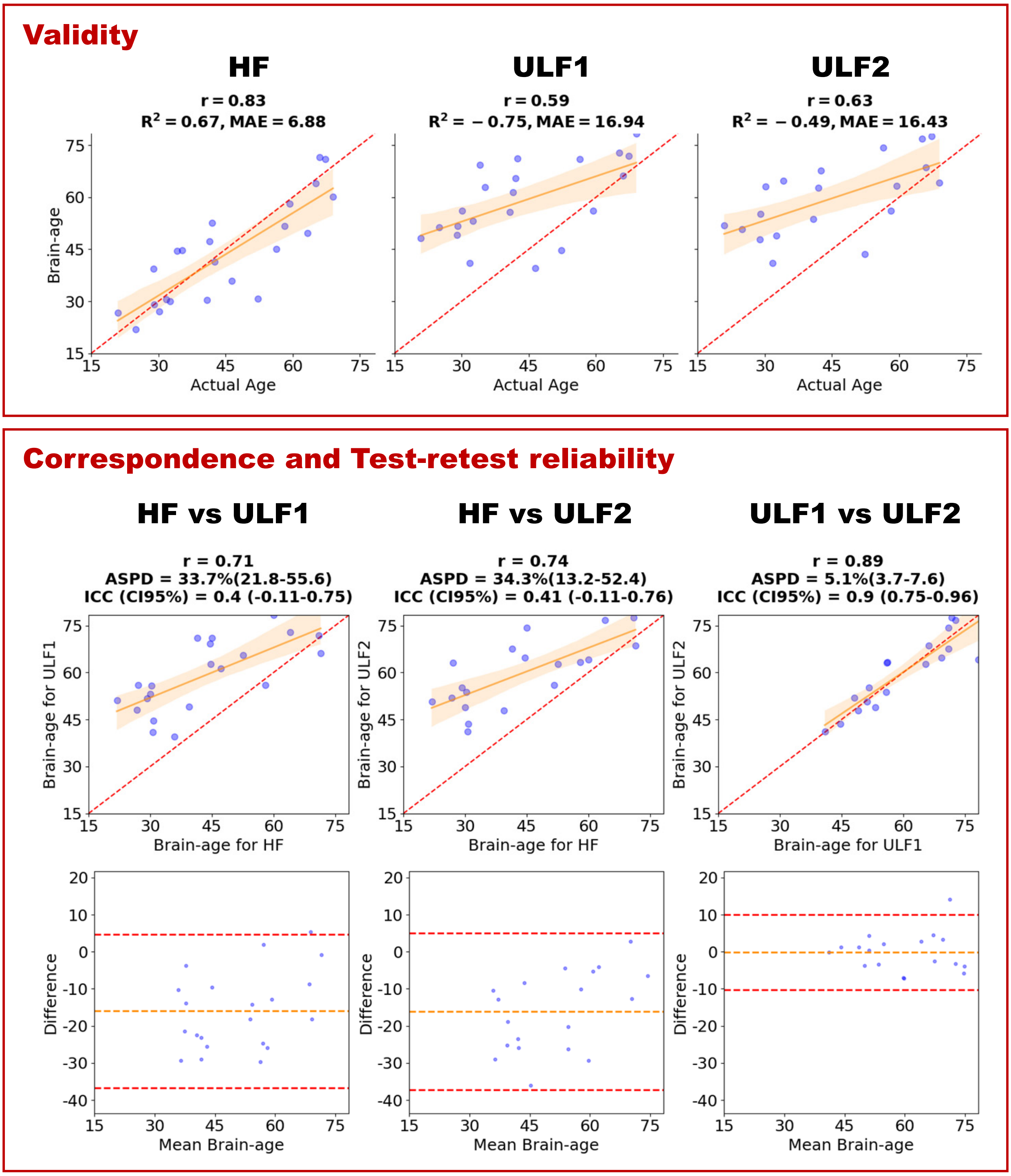

Figure S25: **PyBrainAge on T1+T2-weighted scans using FreeSurfer v7.4 recon-all (with –T2 and –T2pial flags)**. Top panel (validity): three scatter plots of brain-age vs. actual age for (1) HF T1, (2) ULF1 T1, and (3) ULF2 T1. Bottom panel (correspondence and test-retest reliability): the upper row shows, in this order from the left, HF-ULF correspondence scatter plots for (1) HF T1 vs. ULF1 T1, (2) HF T1 vs. ULF2 T1, and (3) test-retest reliability (ULF1 T1 vs. ULF2 T1). The lower row shows the corresponding Bland–Altman plots for the same pairings. Scatter plots include the identity line (red dashed) and a least-squares fit with a 95% confidence band. [Note: re-running the analyses without the –T2pial flag did not change the results]. Abbreviations: Pearson correlation $r$; coefficient of determination $R^2$; mean absolute error (MAE); absolute symmetric percent difference (ASPD) with 95% confidence interval (CI); and intraclass correlation coefficient (ICC) with 95% CI; HF = high-field; ULF1/ULF2 = ultra-low-field sites 1/2.

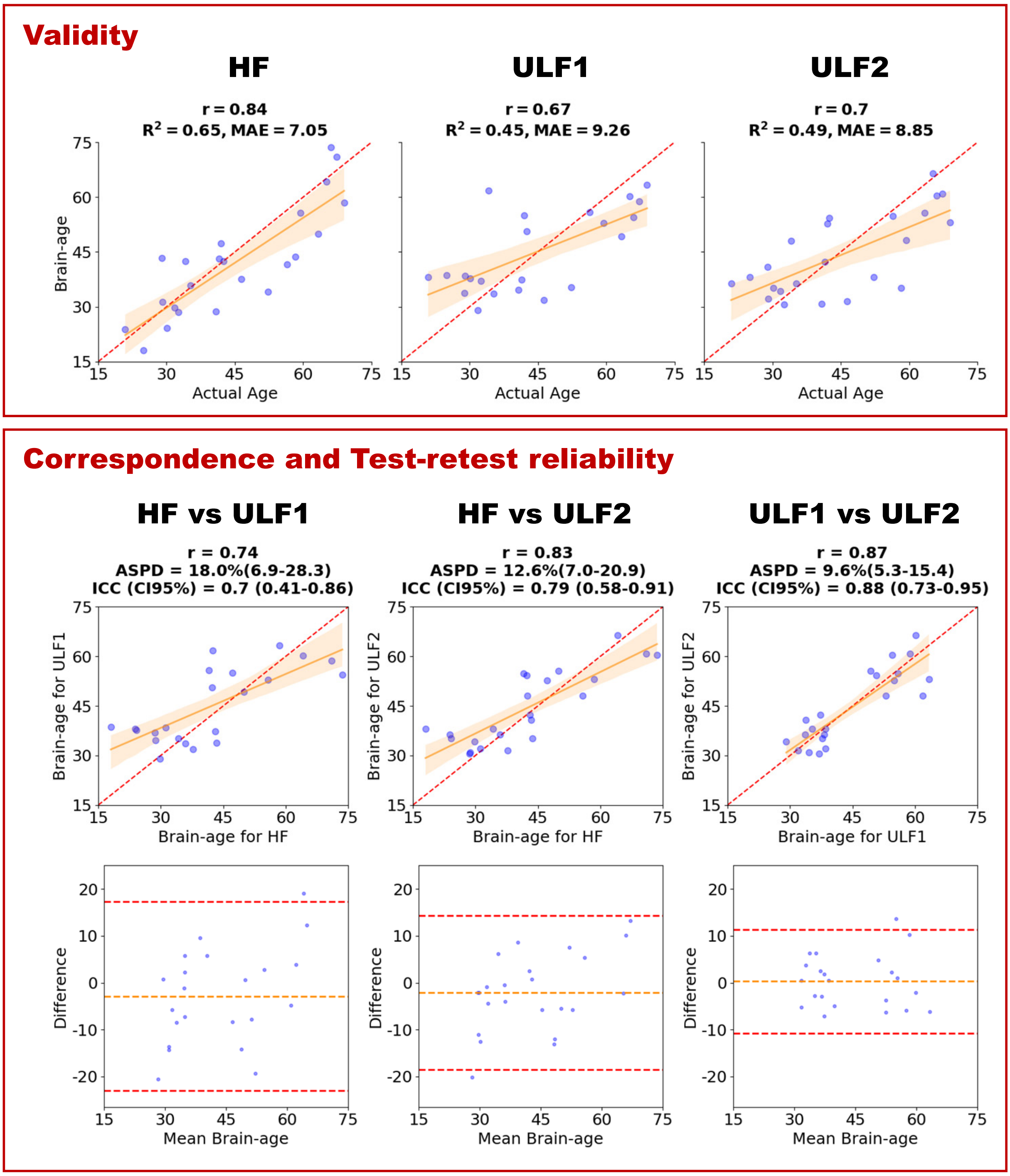

Figure S26: **PyBrainAge on T1-weighted scans with SynthSR (SSR) using FreeSurfer v7.4 recon-all**. Top panel (validity): three scatter plots of brain-age vs. actual age for (1) HF T1, (2) ULF1 T1_SSR, and (3) ULF2 T1_SSR. Bottom panel (correspondence and test-retest reliability): the upper row shows, in this order from the left, HF-ULF correspondence scatter plots for (1) HF T1 vs. ULF1 T1_SSR, (2) HF T1 vs. ULF2 T1_SSR, and (3) test-retest reliability (ULF1 T1_SSR vs. ULF2 T1_SSR). The lower row shows the corresponding Bland–Altman plots for the same pairings. Scatter plots include the identity line (red dashed) and a least-squares fit with a 95% confidence band. Abbreviations: Pearson correlation $r$; coefficient of determination $R^2$; mean absolute error (MAE); absolute symmetric percent difference (ASPD) with 95% confidence interval (CI); and intraclass correlation coefficient (ICC) with 95% CI; HF = high-field; ULF1/ULF2 = ultra-low-field sites 1/2.

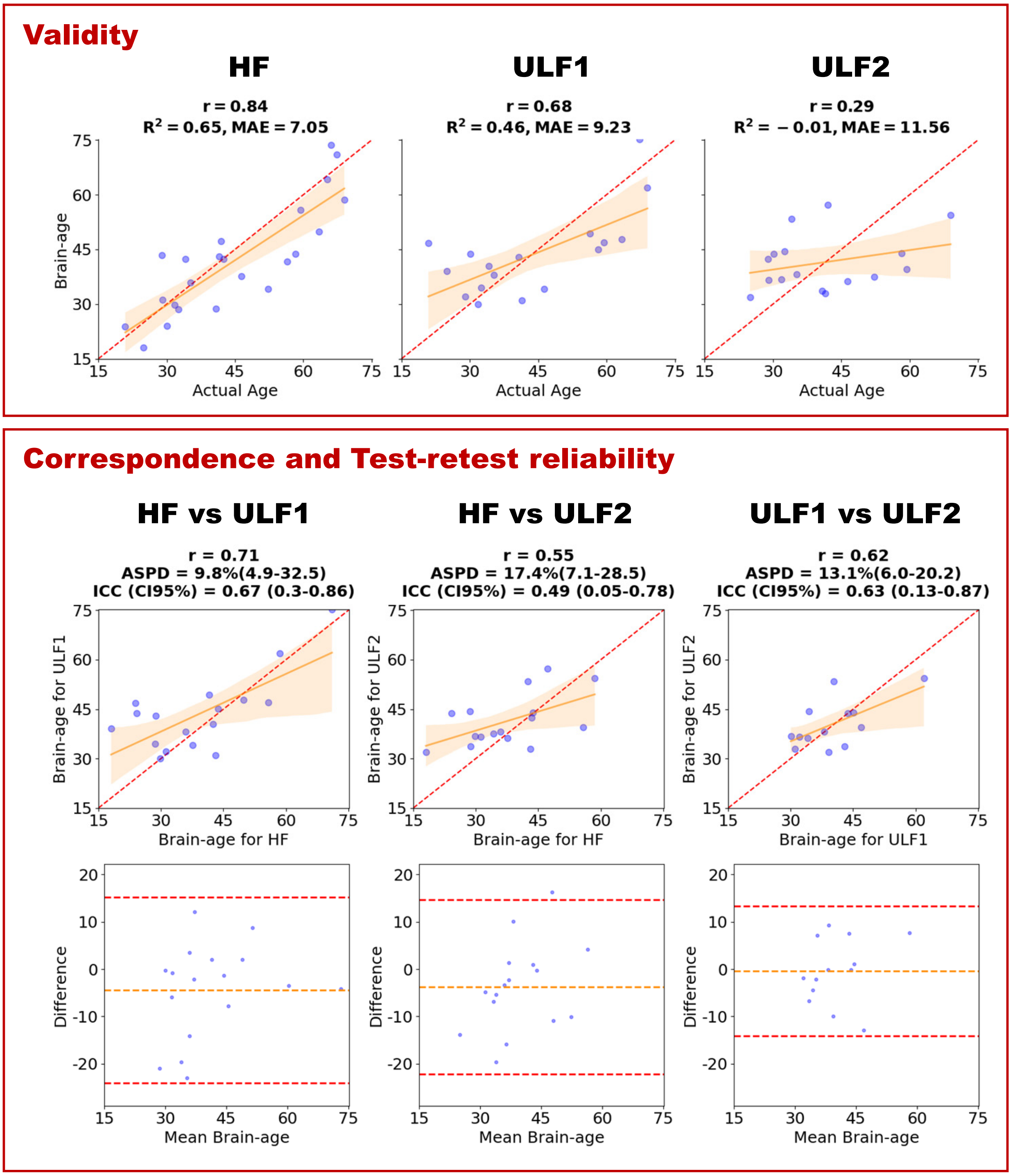

Figure S27: **PyBrainAge on T2-weighted scans with SynthSR (SSR) using FreeSurfer v7.4 recon-all**. Top panel (validity): three scatter plots of brain-age vs. actual age for (1) HF T1, (2) ULF1 T2_SSR, and (3) ULF2 T2_SSR. Bottom panel (correspondence and test-retest reliability): the upper row shows, in this order from the left, HF-ULF correspondence scatter plots for (1) HF T1 vs. ULF1 T2_SSR, (2) HF T1 vs. ULF2 T2_SSR, and (3) test-retest reliability (ULF1 T2_SSR vs. ULF2 T2_SSR). The lower row shows the corresponding Bland–Altman plots for the same pairings. Scatter plots include the identity line (red dashed) and a least-squares fit with a 95% confidence band. Abbreviations: Pearson correlation $r$; coefficient of determination $R^2$; mean absolute error (MAE); absolute symmetric percent difference (ASPD) with 95% confidence interval (CI); and intraclass correlation coefficient (ICC) with 95% CI; HF = high-field; ULF1/ULF2 = ultra-low-field sites 1/2.

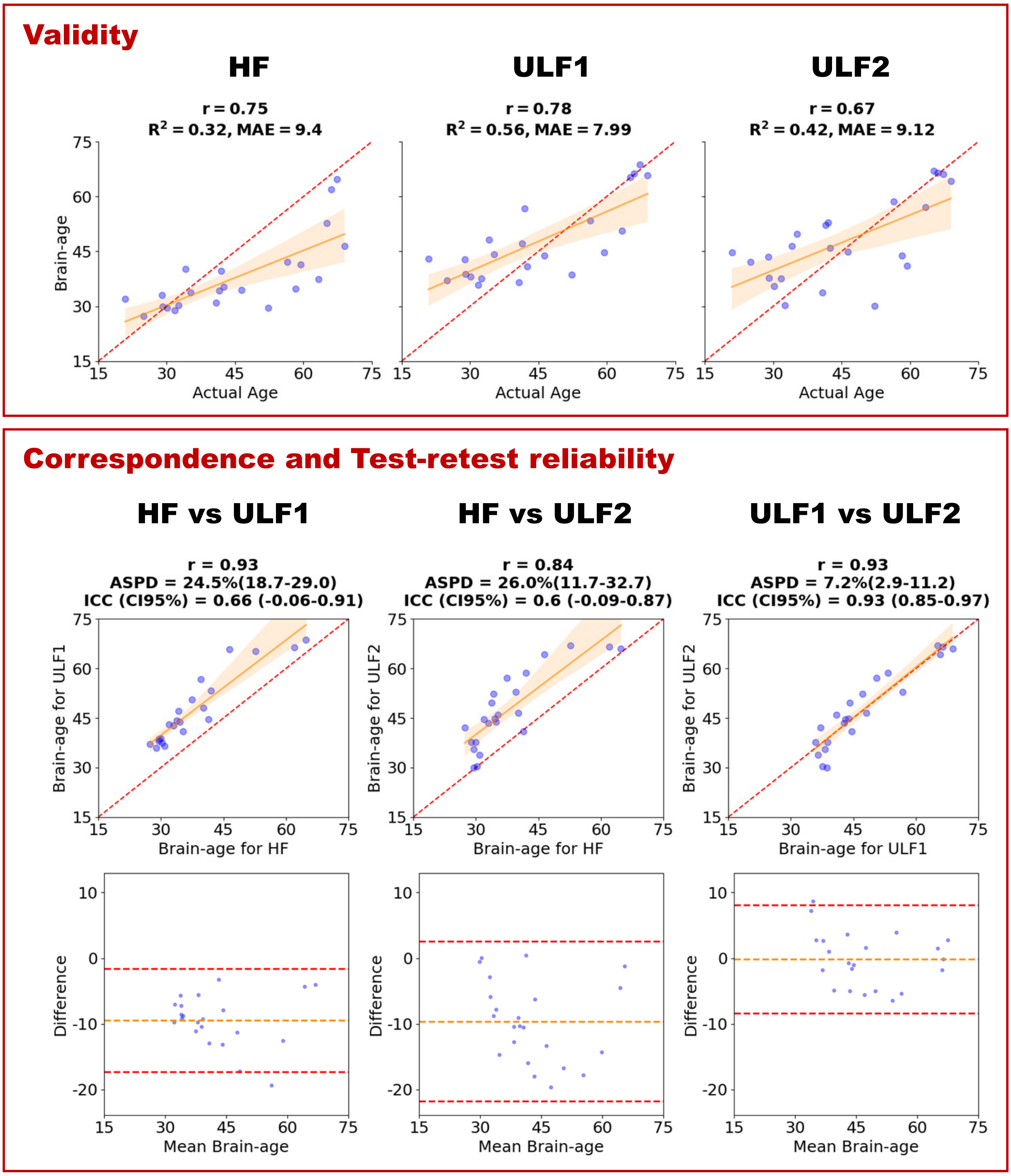

Figure S28: **PyBrainAge on T1-weighted scans using FreeSurfer v7.4 recon-all-clinical**. Top panel (validity): three scatter plots of brain-age vs. actual age for (1) HF T1, (2) ULF1 T1, and (3) ULF2 T1. Bottom panel (correspondence and test-retest reliability): the upper row shows, in this order from the left, HF-ULF correspondence scatter plots for (1) HF T1 vs. ULF1 T1, (2) HF T1 vs. ULF2 T1, and (3) test-retest reliability (ULF1 T1 vs. ULF2 T1). The lower row shows the corresponding Bland–Altman plots for the same pairings. Scatter plots include the identity line (red dashed) and a least-squares fit with a 95% confidence band. Abbreviations: Pearson correlation $r$; coefficient of determination $R^2$; mean absolute error (MAE); absolute symmetric percent difference (ASPD) with 95% confidence interval (CI); and intraclass correlation coefficient (ICC) with 95% CI; HF = high-field; ULF1/ULF2 = ultra-low-field sites 1/2.

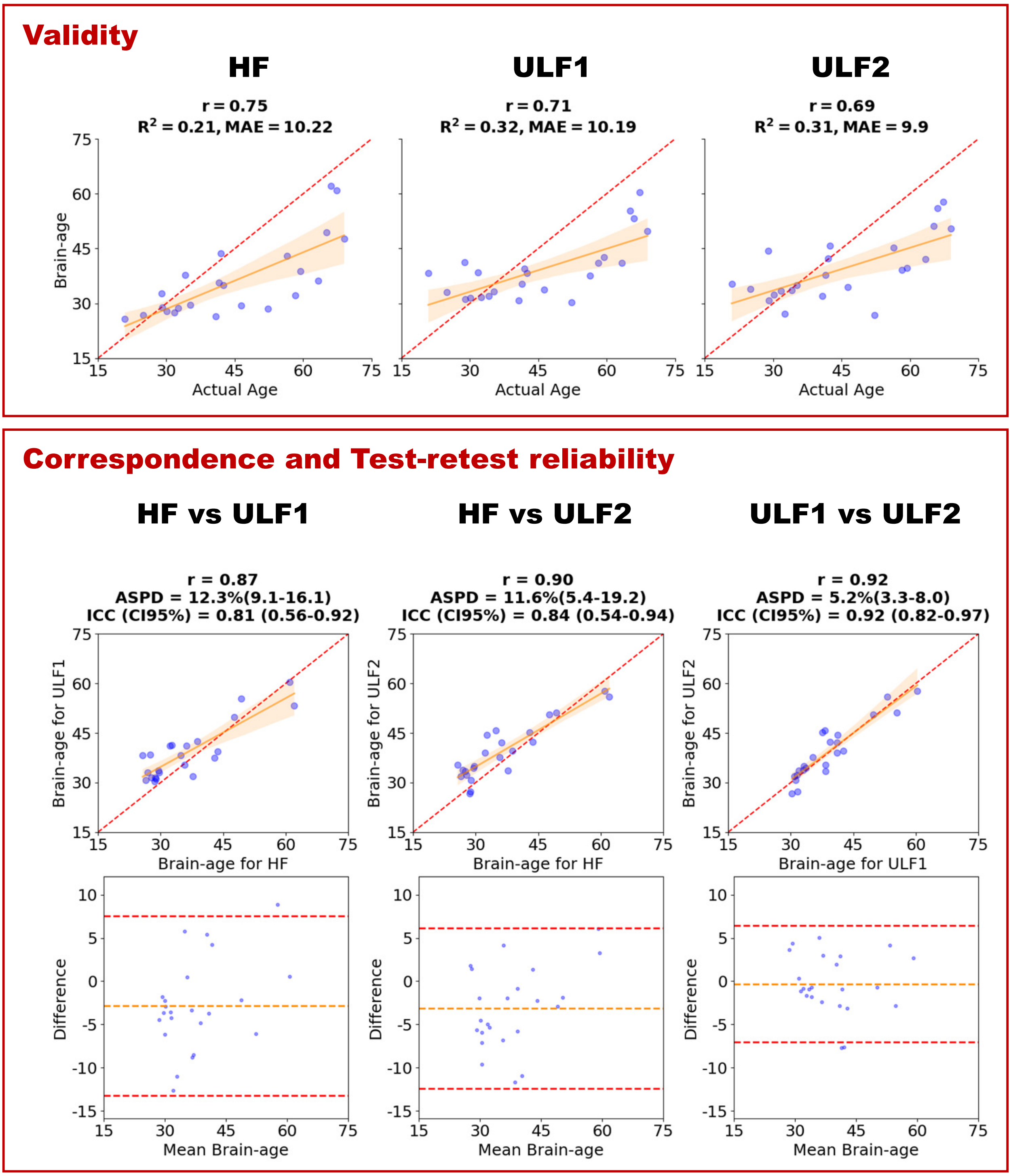

Figure S29: **PyBrainAge on T2-weighted scans using FreeSurfer v7.4 recon-all-clinical**. Top panel (validity): three scatter plots of brain-age vs. actual age for (1) HF T2, (2) ULF1 T2, and (3) ULF2 T2. Bottom panel (correspondence and test-retest reliability): the upper row shows, in this order from the left, HF-ULF correspondence scatter plots for (1) HF T2 vs. ULF1 T2, (2) HF T2 vs. ULF2 T2, and (3) test-retest reliability (ULF1 T2 vs. ULF2 T2). The lower row shows the corresponding Bland–Altman plots for the same pairings. Scatter plots include the identity line (red dashed) and a least-squares fit with a 95% confidence band. Abbreviations: Pearson correlation $r$; coefficient of determination $R^2$; mean absolute error (MAE); absolute symmetric percent difference (ASPD) with 95% confidence interval (CI); and intraclass correlation coefficient (ICC) with 95% CI; HF = high-field; ULF1/ULF2 = ultra-low-field sites 1/2.

#### 5.6 PyBrainAge v8.0.0

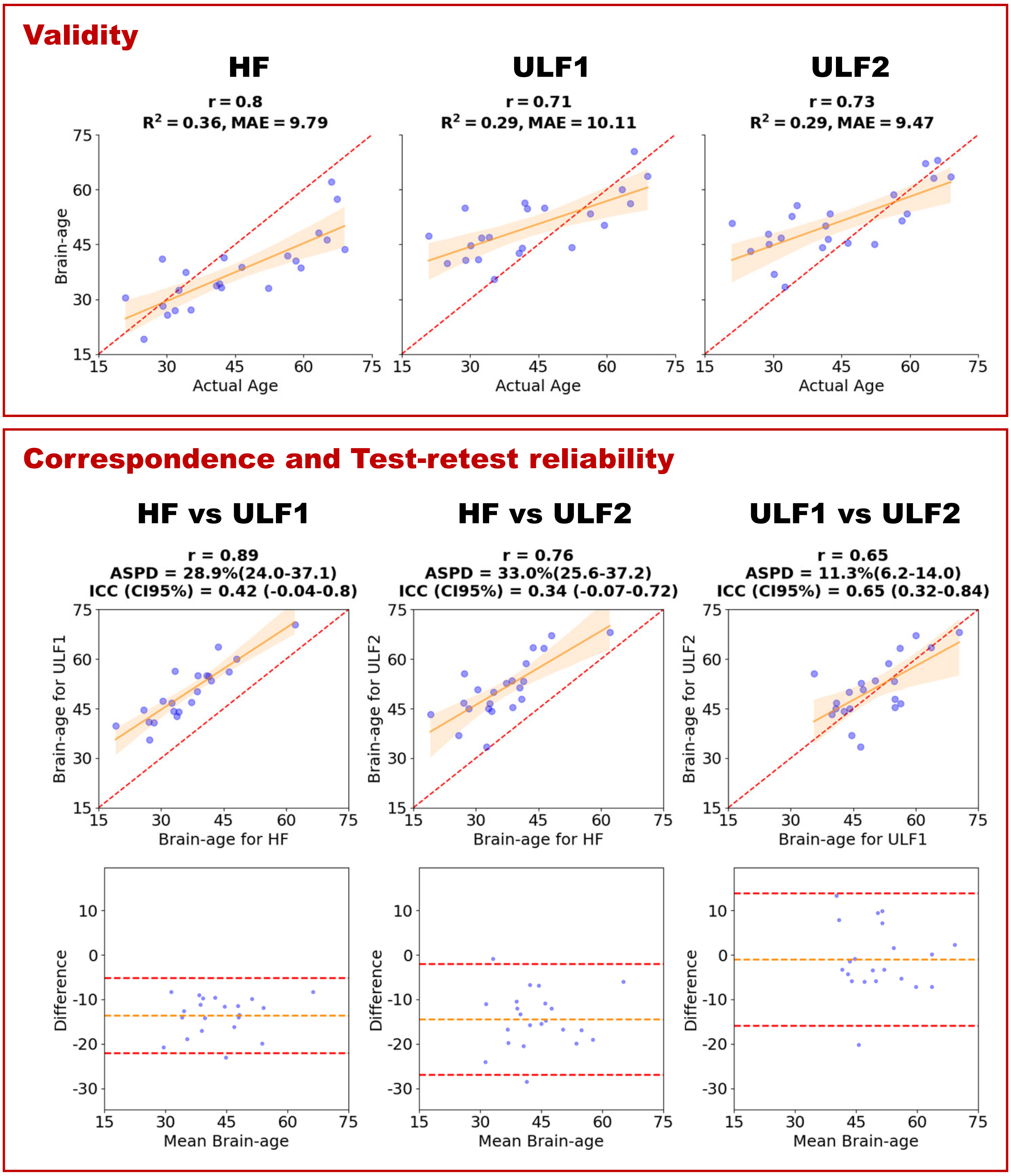

Figure S30: **PyBrainAge on T1-weighted scans using FreeSurfer v8.0 recon-all**. Top panel (validity): three scatter plots of brain-age vs. actual age for (1) HF T1, (2) ULF1 T1, and (3) ULF2 T1. Bottom panel (correspondence and test-retest reliability): the upper row shows, in this order from the left, HF-ULF correspondence scatter plots for (1) HF T1 vs. ULF1 T1, (2) HF T1 vs. ULF2 T1, and (3) test-retest reliability (ULF1 T1 vs. ULF2 T1). The lower row shows the corresponding Bland–Altman plots for the same pairings. Scatter plots include the identity line (red dashed) and a least-squares fit with a 95% confidence band. Abbreviations: Pearson correlation $r$; coefficient of determination $R^2$; mean absolute error (MAE); absolute symmetric percent difference (ASPD) with 95% confidence interval (CI); and intraclass correlation coefficient (ICC) with 95% CI; HF = high-field; ULF1/ULF2 = ultra-low-field sites 1/2.

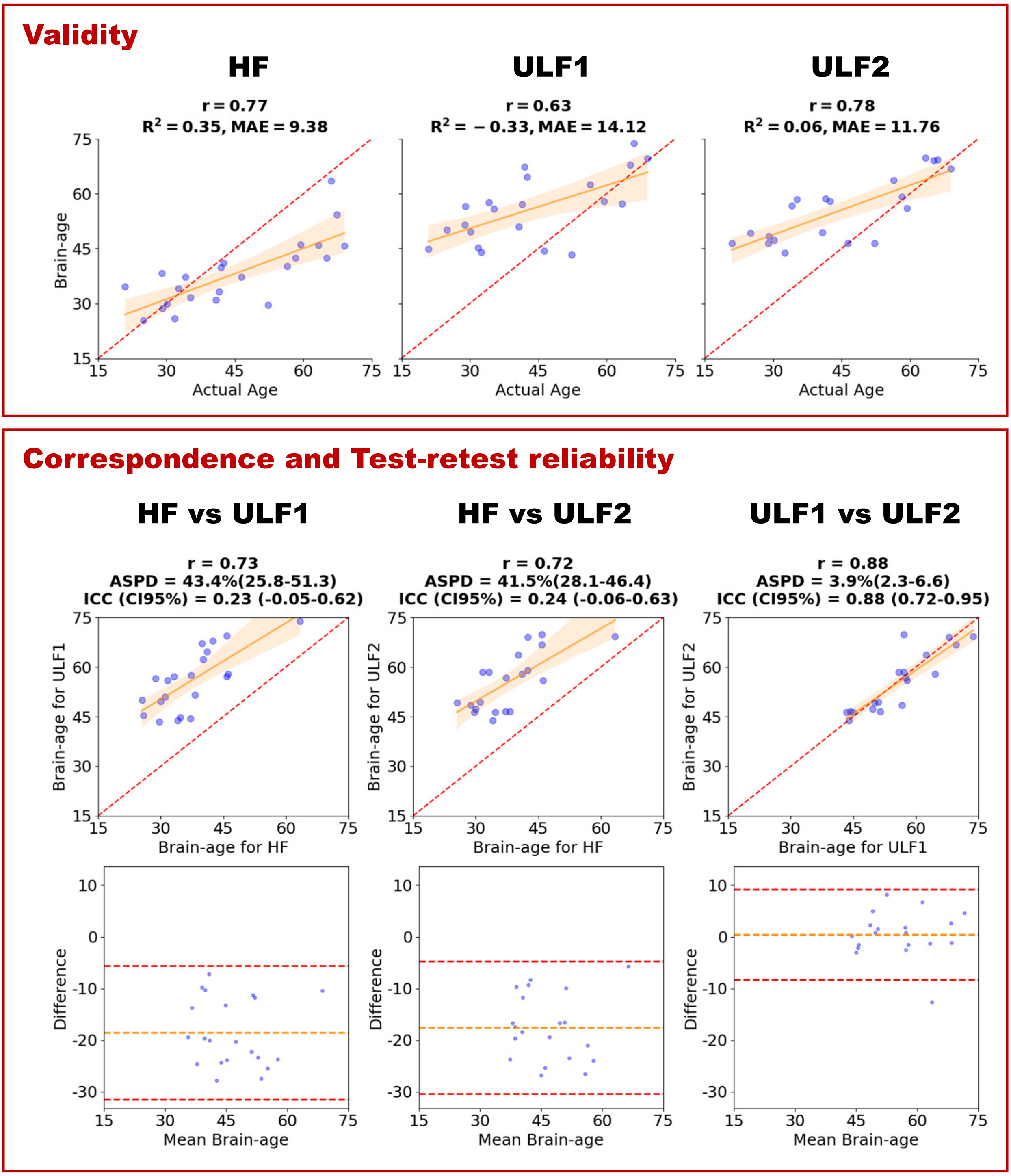

Figure S31: **PyBrainAge on T1+T2-weighted scans using FreeSurfer v8.0 recon-all (with –T2 and –T2pial flags)**. Top panel (validity): three scatter plots of brain-age vs. actual age for (1) HF T1, (2) ULF1 T1, and (3) ULF2 T1. Bottom panel (correspondence and test-retest reliability): the upper row shows, in this order from the left, HF-ULF correspondence scatter plots for (1) HF T1 vs. ULF1 T1, (2) HF T1 vs. ULF2 T1, and (3) test-retest reliability (ULF1 T1 vs. ULF2 T1). The lower row shows the corresponding Bland–Altman plots for the same pairings. Scatter plots include the identity line (red dashed) and a least-squares fit with a 95% confidence band. Abbreviations: Pearson correlation $r$; coefficient of determination $R^2$; mean absolute error (MAE); absolute symmetric percent difference (ASPD) with 95% confidence interval (CI); and intraclass correlation coefficient (ICC) with 95% CI; HF = high-field; ULF1/ULF2 = ultra-low-field sites 1/2.

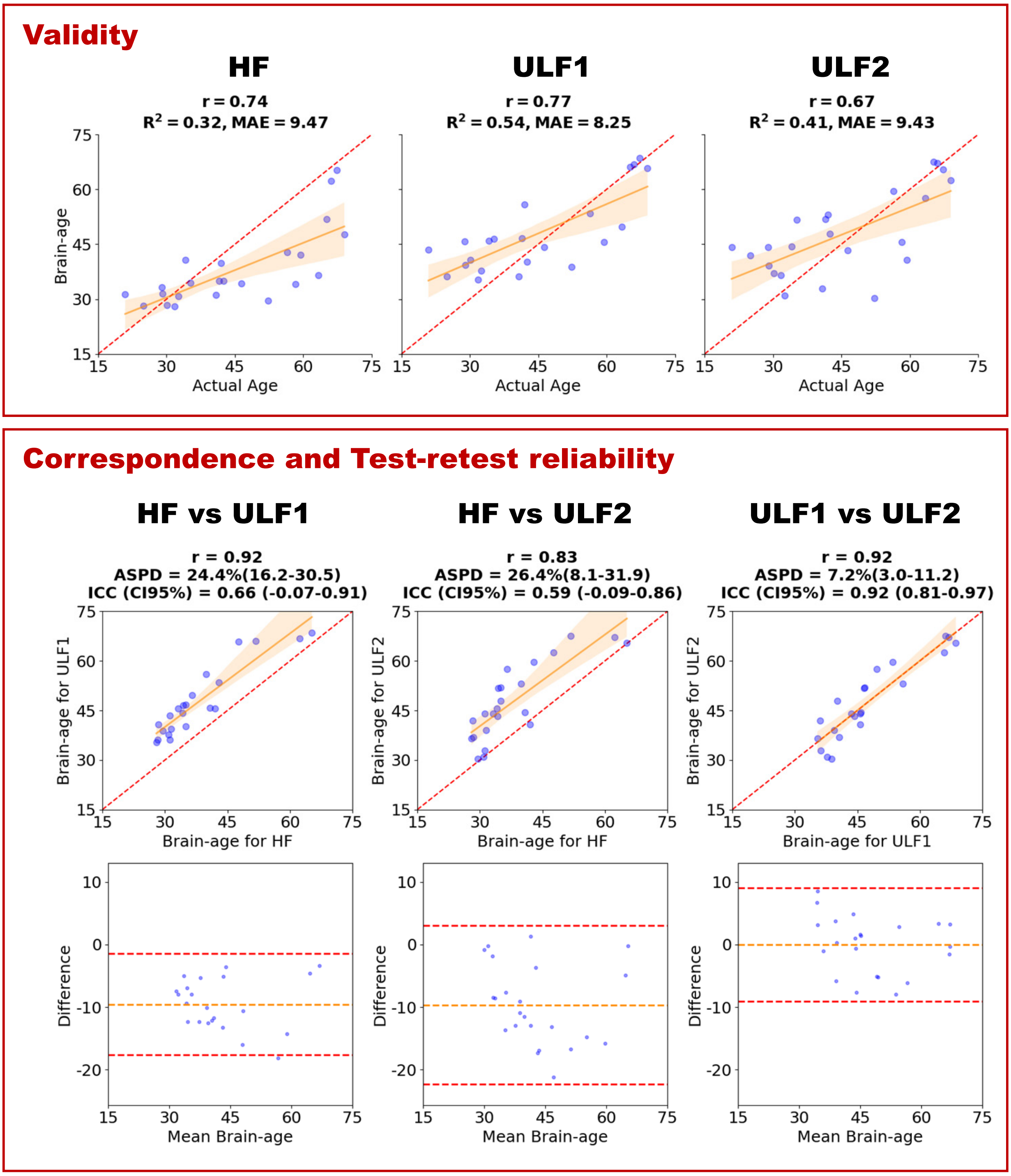

Figure S32: **PyBrainAge on T1-weighted scans using FreeSurfer v8.0 recon-all-clinical**. Top panel (validity): three scatter plots of brain-age vs. actual age for (1) HF T1, (2) ULF1 T1, and (3) ULF2 T1. Bottom panel (correspondence and test-retest reliability): the upper row shows, in this order from the left, HF-ULF correspondence scatter plots for (1) HF T1 vs. ULF1 T1, (2) HF T1 vs. ULF2 T1, and (3) test-retest reliability (ULF1 T1 vs. ULF2 T1). The lower row shows the corresponding Bland–Altman plots for the same pairings. Scatter plots include the identity line (red dashed) and a least-squares fit with a 95% confidence band. Abbreviations: Pearson correlation $r$; coefficient of determination $R^2$; mean absolute error (MAE); absolute symmetric percent difference (ASPD) with 95% confidence interval (CI); and intraclass correlation coefficient (ICC) with 95% CI; HF = high-field; ULF1/ULF2 = ultra-low-field sites 1/2.

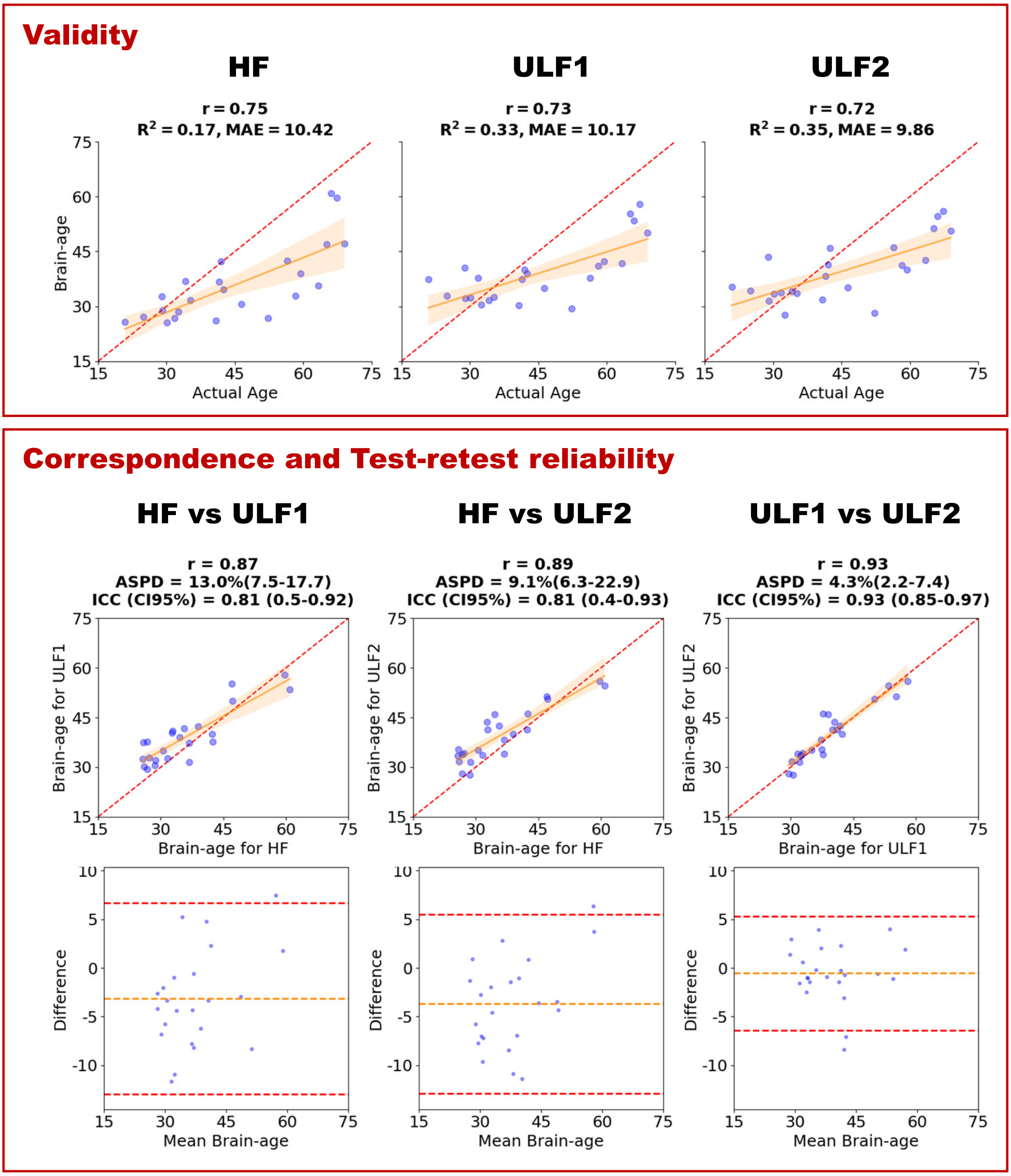

Figure S33: **PyBrainAge on T2-weighted scans using FreeSurfer v8.0 recon-all-clinical**. Top panel (validity): three scatter plots of brain-age vs. actual age for (1) HF T2, (2) ULF1 T2, and (3) ULF2 T2. Bottom panel (correspondence and test-retest reliability): the upper row shows, in this order from the left, HF-ULF correspondence scatter plots for (1) HF T2 vs. ULF1 T2, (2) HF T2 vs. ULF2 T2, and (3) test-retest reliability (ULF1 T2 vs. ULF2 T2). The lower row shows the corresponding Bland–Altman plots for the same pairings. Scatter plots include the identity line (red dashed) and a least-squares fit with a 95% confidence band. Abbreviations: Pearson correlation $r$; coefficient of determination $R^2$; mean absolute error (MAE); absolute symmetric percent difference (ASPD) with 95% confidence interval (CI); and intraclass correlation coefficient (ICC) with 95% CI; HF = high-field; ULF1/ULF2 = ultra-low-field sites 1/2.

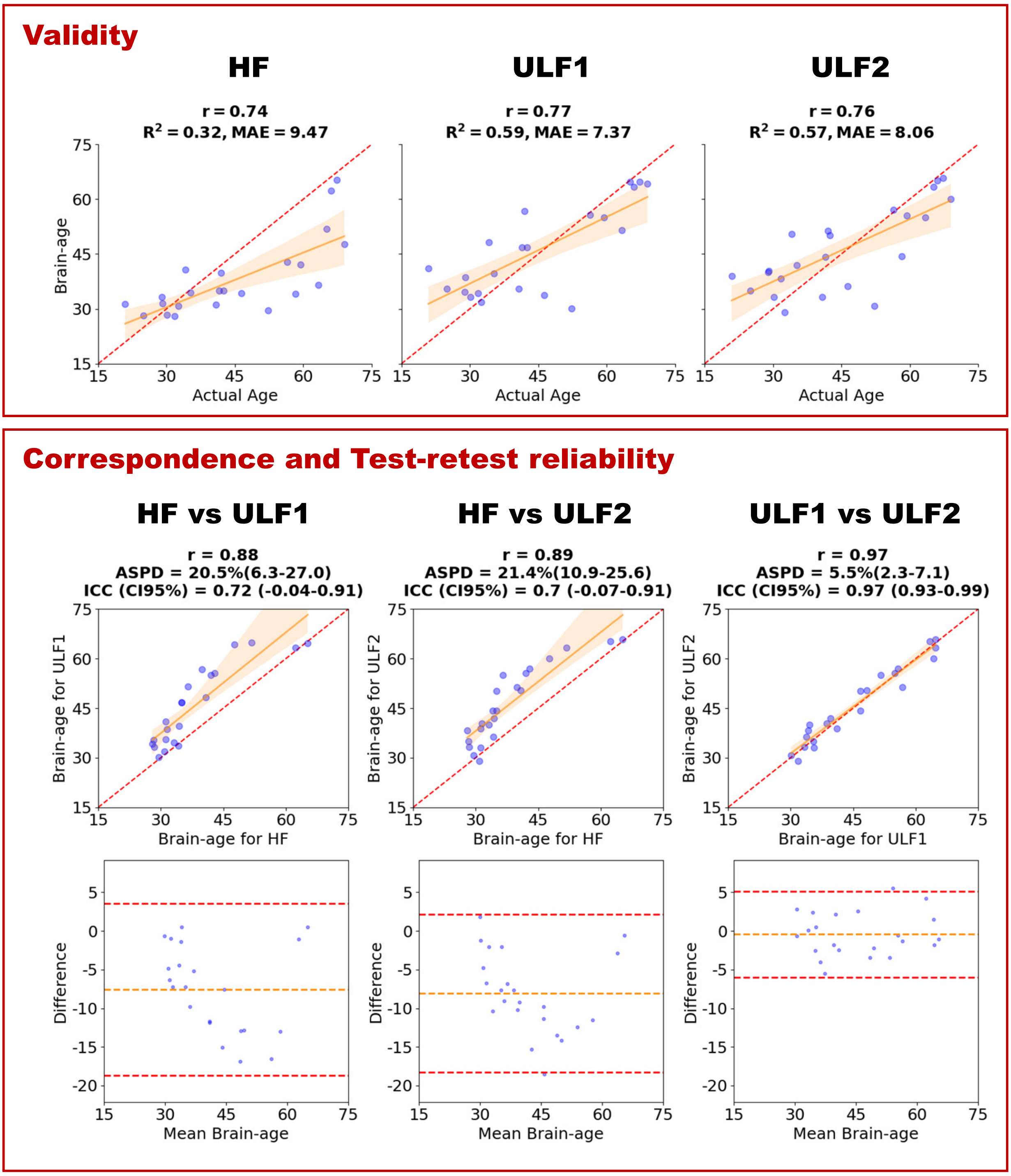

Figure S34: **PyBrainAge on T1-weighted scans with SynthSR (SSR) using FreeSurfer v8.0 recon-all-clinical**. Top panel (validity): three scatter plots of brain-age vs. actual age for (1) HF T1, (2) ULF1 T1_SSR, and (3) ULF2 T1_SSR. Bottom panel (correspondence and test-retest reliability): the upper row shows, in this order from the left, HF-ULF correspondence scatter plots for (1) HF T1 vs. ULF1 T1_SSR, (2) HF T1 vs. ULF2 T1_SSR, and (3) test-retest reliability (ULF1 T1_SSR vs. ULF2 T1_SSR). The lower row shows the corresponding Bland–Altman plots for the same pairings. Scatter plots include the identity line (red dashed) and a least-squares fit with a 95% confidence band. Abbreviations: Pearson correlation $r$; coefficient of determination $R^2$; mean absolute error (MAE); absolute symmetric percent difference (ASPD) with 95% confidence interval (CI); and intraclass correlation coefficient (ICC) with 95% CI; HF = high-field; ULF1/ULF2 = ultra-low-field sites 1/2.

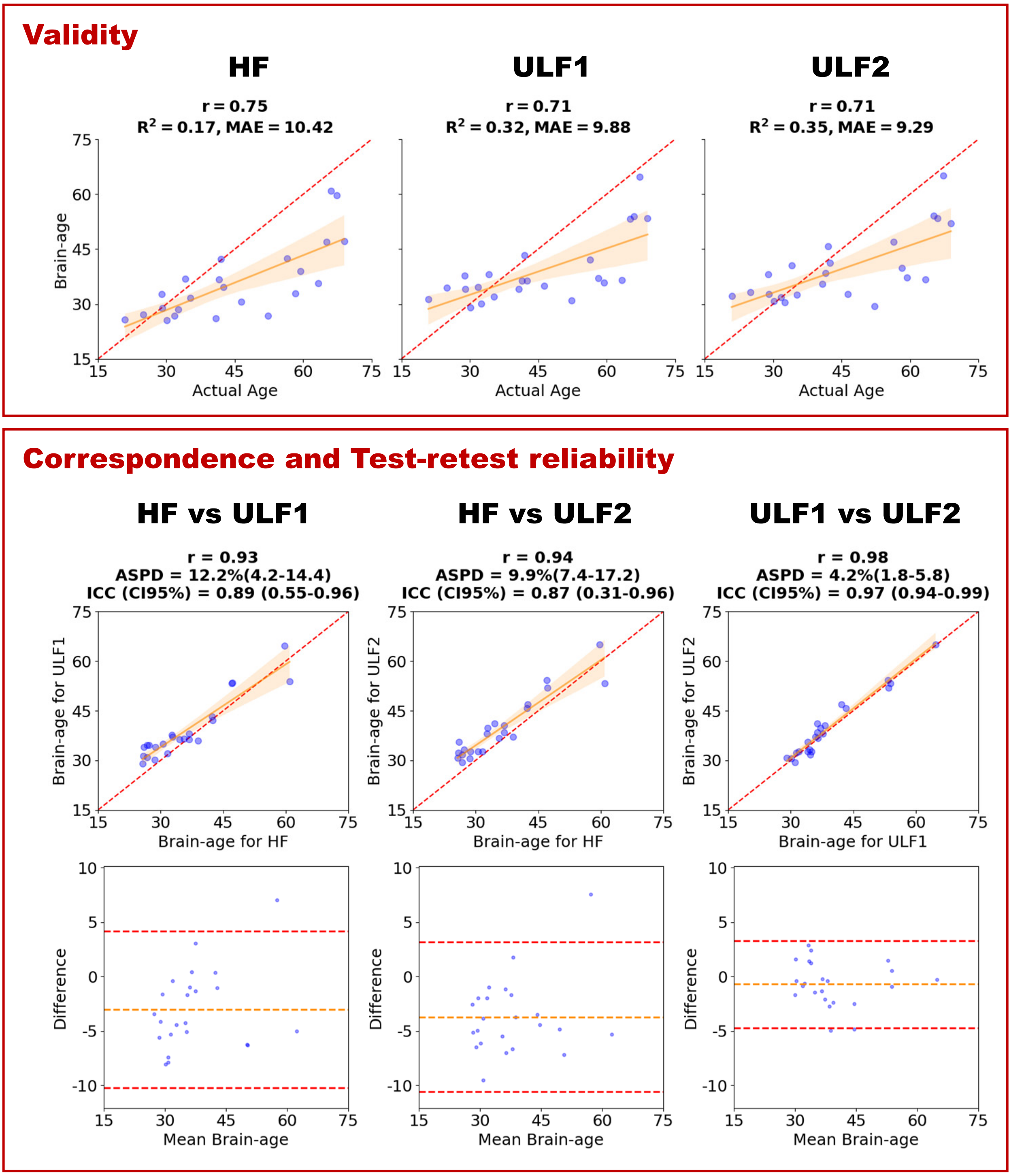

Figure S35: **PyBrainAge on T2-weighted scans with SynthSR (SSR) using FreeSurfer v8.0 recon-all-clinical**. Top panel (validity): three scatter plots of brain-age vs. actual age for (1) HF T2, (2) ULF1 T2_SSR, and (3) ULF2 T2_SSR. Bottom panel (correspondence and test-retest reliability): the upper row shows, in this order from the left, HF-ULF correspondence scatter plots for (1) HF T2 vs. ULF1 T2_SSR, (2) HF T2 vs. ULF2 T2_SSR, and (3) test-retest reliability (ULF1 T2_SSR vs. ULF2 T2_SSR). The lower row shows the corresponding Bland–Altman plots for the same pairings. Scatter plots include the identity line (red dashed) and a least-squares fit with a 95% confidence band. Abbreviations: Pearson correlation $r$; coefficient of determination $R^2$; mean absolute error (MAE); absolute symmetric percent difference (ASPD) with 95% confidence interval (CI); and intraclass correlation coefficient (ICC) with 95% CI; HF = high-field; ULF1/ULF2 = ultra-low-field sites 1/2.
